# Therapeutic efficacy study on shoulder impingement syndrome in swimmers: a network meta-analysis

**DOI:** 10.64898/2026.06.11.26355435

**Authors:** Yufeng Chuo, Jiaqi Li, Yutong Duan, Tianle Wang

## Abstract

Shoulder impingement syndrome (SIS), including subacromial impingement and rotator cuff tendinitis, is commonly caused by repetitive swimming movements and associated shoulder joint dysfunction. Despite numerous available treatment options, no consensus exists on the most effective treatment option. Therefore, this systematic review and network meta-analysis aimed to investigate treatment methods for SIS in swimmers. Using a frequentist framework and Cochrane PICOS principles, we compared SIS treatments, constructed network evidence diagrams, and assessed heterogeneity. A total of 45 studies were included in the qualitative synthesis, and 42 contributed to the network meta-analysis, comprising 1752 participants, 9 treatment categories, and outcome measures. For pain outcomes, some adjunctive interventions combined with exercise showed favorable ranking probabilities, although several estimates were accompanied by wide confidence intervals. For shoulder range-of-motion outcomes, taping, acupuncture, manual therapy, and sport-specific training showed favorable effects in selected comparisons, particularly for external and internal rotation. According to surface under the cumulative ranking curve (SUCRA) rankings, exercise combined with medium-frequency therapy ranked highly for pain reduction, whereas exercise combined with acupuncture or extracorporeal shock wave therapy ranked highly for shoulder flexion. Exercise combined with taping ranked highly for external rotation, and exercise combined with manual therapy ranked highly for internal rotation. However, the interpretation of ranking results should remain cautious because uncertainty and inconsistency were present in some comparisons. Exercise-based rehabilitation appears to remain central to the management of SIS in swimmers. Several adjunctive interventions showed favorable findings for selected outcomes, especially pain relief and shoulder rotational function. However, the available evidence was affected by heterogeneity, inconsistency, and imprecision across some treatment comparisons. More rigorously designed swimmer-specific randomized controlled trials are needed before firm treatment hierarchies can be established.

**Trial registration:** The protocol for this systematic review is registered with PROSPERO (www.crd.york.ac.uk/PROSPERO; registration number: CRD42024498851). The first submission of PROSPERO was on January 15, 2024, and it was revised and updated on March 25, 2026.

## Introduction

In swimming, shoulder impingement syndrome (SIS), or otherwise, “swimmer’s shoulder,” is the subacromial impingement syndrome and rotator cuff tendinitis caused by repetitive swimming movements and associated shoulder joint dysfunction. Kennedy and Hawkins [1] conducted the earliest epidemiological research on shoulder joint injuries in 2,496 Canadian swimmers as their orthopedic consultant in the 1972 Munich Olympics. They found a relatively high incidence of orthopedic diseases in swimmers; of 43 visits, 16 (37.2%) were owing to shoulder joint injuries. Additionally, their large-scale survey of the entire Canadian swimming team revealed that 90% of sports injuries involved the shoulder, knee, calf, and ankle joints but most commonly the shoulder [1]; thus, they introduced the concept of “swimmer’s shoulder.”

Other studies later emerged, focusing on college swimmers, reporting “athlete exposure” rate, defined as the ratio of “injury occurrences” to “training exposure instances.” “Exposure instances” refer to the number of times an athlete participates in a training session or competition (such as races, water training, strength training, or cross-training). Numerous studies based on the National Collegiate Athletic Association report that athlete exposure values for college swimmers range from 1.43/1000 to 5.5/1000, with shoulder joint injuries and pain being the most common. For example, a 7-year longitudinal study by McFarland and Wasik [2] reported an athlete exposure value of 2.12/1000 for National Collegiate Athletic Association female swimmers, with 55% of injuries involving the shoulder joint.

Wolf et al. [3] reported values of 4/1000 and 3.78/1000 for males and females, respectively, with 41.8% shoulder joint involvement. According to Chase et al. [4], 5.5/1000 was reported among swimmers and 38.1% for the shoulder joint. For male and female National Collegiate Athletic Association swimmers in training and competition, Kerr et al. [5] reported 1.43/1000 vs. 1.96/1000 and 1.62/1000 vs. 1.70/1000, respectively, with shoulder pain and injury incidence of 34.7% and 31.3% for males and females, respectively. Furthermore, a study on the 2009, 2013, and 2015 World Swimming Championships reported that 15.0%, 22.1%, and 19.2% of athletes reported shoulder pain and injury, respectively [6]. These findings suggest that the shoulder joint is the most vulnerable area for injury during high-intensity competitions.

The prevalence of illness among college swimmers reported by Stocker et al. [7] was 47%, whereas McMaster [8] reported 10%, 13%, and 26% prevalence among swimmers of different age groups and national teams. Under high-intensity training, swimmers are prone to supraspinatus tendonitis, accompanied by tendon thickening. As the rotator cuff tendons become damaged or thickened, the gap between the acromion and the humerus narrows, increasing the risk of subacromial impingement, which can lead to chronic pain [9]. Additionally, other studies report that prolonged swimming training may lead to an imbalance in the range of motion and muscle strength of the shoulder joint, and this imbalance is closely related to the reduction in the subacromial space [10].

Richardson [11] indicated that the shoulder movement trajectory of athletes in competitive swimming is highly similar to that of athletes in throwing events (such as baseball and tennis). However, owing to the water resistance, the required range of shoulder external rotation (ER) and internal rotation (IR) speed for swimmers is relatively lower and differs significantly from those in throwing events. Moreover, the shoulder movement frequency of competitive swimmers is much higher than that of athletes in other sports.

Guoping et al. [12] and Minghui et al. [13] found a 6.6% and 47.44% shoulder injury incidence rate among professional swimming team athletes in China, respectively. Studies report that elite swimmers can train at distances ranging from 9,000 to 18,000 m daily and perform over 1 million strokes per week, primarily involving overhead shoulder rotations. Among them, freestyle swimmers have the highest shoulder injury rate. During freestyle swimming, the arm must perform various motions, including entering the water, extending, pulling, exiting the water, and moving the arm in the air. Early in the stroke phase and during the early arm recovery phase, the long head of the biceps and supraspinatus may collide with the front of the shoulder joint. This occurs particularly with the technique of entering the water with the thumb first, which increases the risk of impact between the long head of the biceps and the shoulder joint, thus raising the risk of shoulder injury. Additionally, asymmetrical body rolling or a unilateral breathing pattern may also lead to injuries. When one side of the body’s role is restricted, or the non-breathing side is limited, compensatory movements occur in the contralateral shoulder during propulsion, further increasing the risk of injury.

Zhiyong et al. [14] investigated the characteristics of injury-prone areas among 124 elite swimmers from the Chinese national training team. They showed that 86 athletes reported shoulder injuries, making it the most commonly affected area. Swimming relies on the shoulder joint as the primary axis for stroke execution, subjecting the shoulders to prolonged high-load conditions, which increases the risk of chronic shoulder pain. From a physiological perspective, external forces may cause damage to the surrounding muscles of the shoulder joint, leading to inflammatory responses in the rotator cuff and biceps brachii. Being a joint with a wide range of motion, the shoulder joint has relatively poor stability, making it more susceptible to injury. Additionally, the biceps brachii is attached to the shoulder joint, and during IR movements, the humeral head may compress the surrounding blood vessels and tendons, leading to localized ischemia and subsequent inflammation.

Similarly, repetitive abduction and IR movements of the biceps brachii may result in inflammation due to insufficient oxygen supply. During long-term swim training, repetitive stroking motions may lead to laxity of the anterior shoulder joint capsule, a primary cause of the swimmer’s shoulder. Moreover, muscle imbalances around the shoulder joint are also a key contributing factor to the swimmer’s shoulder.

However, the abundant treatment options do not necessarily contribute to doctors’ decision-making, and no consensus exists on the appropriate treatment option. Currently, many randomized controlled trials have compared the effectiveness of different treatment methods, supporting certain conclusions. Some systematic reviews are also only focused on pairwise comparisons of different treatment methods, but reviews of all available treatment methods are excluded. Owing to these limitations of existing reviews and since many relatively new studies have been published, an accurate and comprehensive review of this topic is urgently needed to account for all treatment methods.

In this study, we aimed to systematically summarize the treatment methods for subacromial impingement syndrome, including exercise therapy, pharmacological treatment, physical therapy, traditional Chinese medicine, and surgical interventions, by identifying existing randomized controlled trials on this condition (Table 1). Furthermore, statistical methods on network meta-analysis can calculate the rank probability of each treatment. In this type of analysis, researchers may consider all possible related treatments. Thus, we believe that these results can support doctors, coaches, and athletes in decision-making.

**Table 1.**
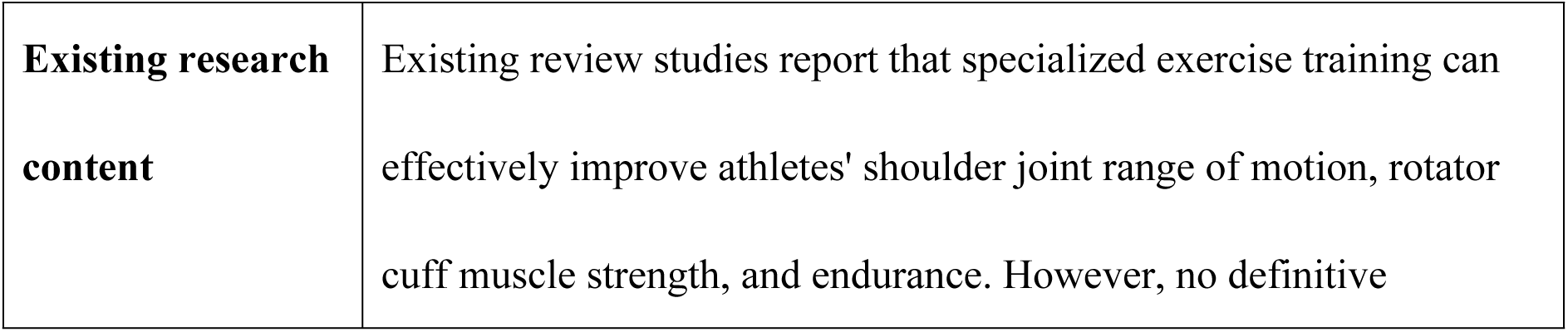
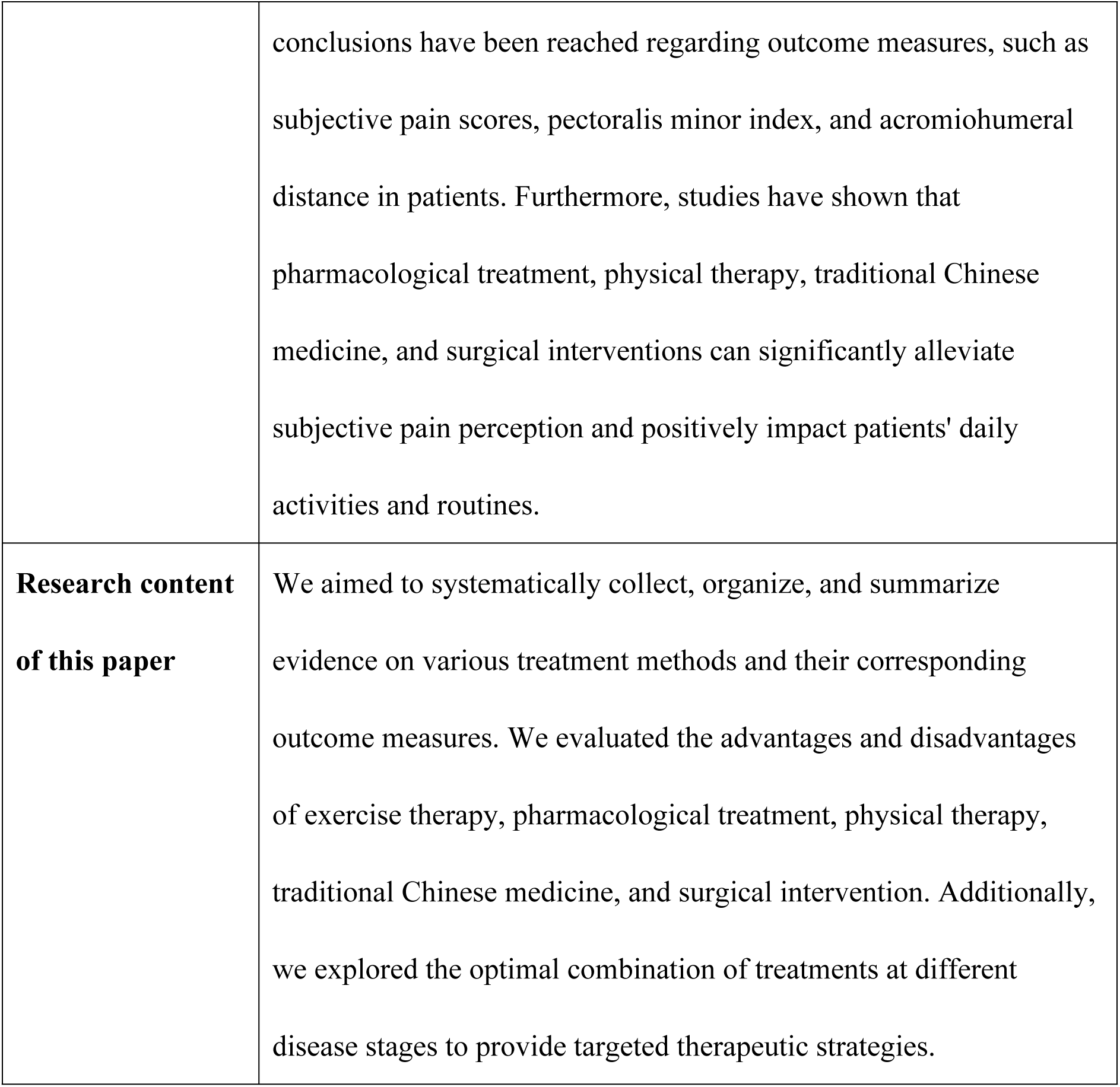
Research innovation.

|  |  |
| --- | --- |
| **Existing research content** | Existing review studies report that specialized exercise training can effectively improve athletes' shoulder joint range of motion, rotator cuff muscle strength, and endurance. However, no definitive |
|  | conclusions have been reached regarding outcome measures, such as subjective pain scores, pectoralis minor index, and acromiohumeral distance in patients. Furthermore, studies have shown that pharmacological treatment, physical therapy, traditional Chinese medicine, and surgical interventions can significantly alleviate subjective pain perception and positively impact patients' daily activities and routines. |
| **Research content of this paper** | We aimed to systematically collect, organize, and summarize evidence on various treatment methods and their corresponding outcome measures. We evaluated the advantages and disadvantages of exercise therapy, pharmacological treatment, physical therapy, traditional Chinese medicine, and surgical intervention. Additionally, we explored the optimal combination of treatments at different disease stages to provide targeted therapeutic strategies. |

## Materials and Methods

### Aim, design and setting

We aimed to compare the relative efficacy of available interventions for shoulder impingement syndrome (SIS) in swimmers. It was conducted as a systematic review and frequentist network meta-analysis of randomized controlled trials, based on a structured process of literature searching, study selection, data extraction, and quantitative synthesis across major international and Chinese electronic databases.

### Protocol and reporting

The protocol for this systematic review is registered with PROSPERO (www.crd.york.ac.uk/PROSPERO; registration number: CRD42024498851). The first submission of PROSPERO was on January 15, 2024, and it was revised and updated on March 25, 2026. The review was conducted and reported in accordance with the Preferred Reporting Items for Systematic Reviews and Meta-Analyses (PRISMA) statement and relevant methodological guidance for network meta-analysis.

### Eligibility criteria and study framework

Eligibility was defined a priori according to the participants, interventions, comparisons, outcomes, and study design framework. To avoid redundancy, the detailed inclusion and exclusion criteria used during full-text screening are presented in Table 2, whereas the broader PICOS framework is summarized in Table 3.

**Table 2.** Basic criteria for literature search.

| Item | Content |
| --- | --- |
| Databases Searched | PubMed, Medline, EMbase, Web of Science Core Collection, Scopus, Cochrane Library, EBSCO, CNKI |
| Search scope | Title, Keywords, Abstract, Subject |
| Search time period | No limit |
| Database search timeline | PubMed: 1983–2026<br>Medline: 1966–2026<br>EMbase: 1966–2026<br>Web of Science Core Collection: 1990–2026<br>Scopus: 1968–2026<br>Cochrane Library: 2006–2026<br>CNKI: 1999–2026 |
| Study design | Randomized controlled trials |
| Languages | Chinese, English |
| Initial screening date | January 1, 2026 |
| Screening date | January 1, 2026 |

**Table 3.** Participants, interventions, comparisons, outcomes, and study design principles.

| Items | Detailed inclusion criteria |
| --- | --- |
| Participants | Athletes with subacromial impingement syndrome |
| Interventions | Physical therapy modalities, traditional Chinese medicine treatments, pharmacological treatments, surgical interventions, and specialized exercise training therapy |
| Control/comparison | Non-placebo, exercise therapy group, or other intervention methods |
| Outcome | Subjective assessments, physical performance indicators, and movement tests |
| Study design | Randomized controlled trials |

Studies were eligible if they met the following criteria: participants were competitive, trained, or professional swimmers diagnosed with subacromial or other closely related non-traumatic shoulder impingement conditions; interventions involved at least one eligible treatment approach for SIS; comparators included exercise therapy, placebo or sham treatment, or another active intervention; outcomes reported at least one clinically relevant measure of pain, function, range of motion, muscle performance, or structural status; and the study design was a randomized controlled trial. Studies were excluded if they involved non-swimmer populations, traumatic shoulder injuries, non-randomized designs, insufficient outcome reporting, or unavailable full texts.

### Information sources and search strategy

A comprehensive literature search was performed in PubMed, MEDLINE, Embase, Web of Science Core Collection, Scopus, the Cochrane Library, EBSCO, and CNKI from database inception to January 1, 2026. The search covered titles, abstracts, keywords, and subject headings without restriction on publication year. Only randomized controlled trials published in English or Chinese were considered.

The search strategy combined controlled vocabulary and free-text terms related to swimmers, shoulder impingement or rotator cuff-related disorders, and therapeutic interventions. Boolean operators (AND/OR) were used to combine search terms. The detailed PubMed search strategy is provided in Table 4, and equivalent logic was adapted for the other databases.

**Table 4.** Keywords searched in PubMed database.

| Step number | Search strategy |
| --- | --- |
| #1 | (“Rotator Cuff”[MeSH Terms] OR “Rotator Cuff Injuries”[MeSH Terms] OR (“Shoulder”[MeSH Terms] OR “Shoulder Pain”[MeSH Terms]) OR “subacrom*”[Title/Abstract] OR “supraspinat*”[Title/Abstract] OR “swimming athlete*”[Title/Abstract] OR “swimmer”[MeSH Terms] ) |
| #2 | AND(“Shoulder Impingement Syndrome”[MeSH Terms] OR “impingement”[Title/Abstract]) |
| #3 | AND (“Factorial”[Title/Abstract] OR “Random”[Title/Abstract] OR “single blind procedure”[Title/Abstract] OR “double blind procedure”[Title/Abstract] OR “visual analogue scale”[Title/Abstract] OR “Numerical Rating Scale”[Title/Abstract] OR “Pectoralis Minor Index”[Title/Abstract] OR “Constant Murley Score”[Title/Abstract] OR “Disabilities of the Arm, Shoulder and Hand”[Title/Abstract] OR “Rang of motion”[Title/Abstract]) |
| #4 | (“expectant treatment”[Title/Abstract] OR “conservative treatment”[Title/Abstract] OR “non invasive treatment”[Title/Abstract] OR |
| #5 | “clinic treatment”[Title/Abstract] OR “operative treatment”[Title/Abstract]<br>OR “traumatic therapy”[Title/Abstract]<br>OR(“Combined”[Title/Abstract] “OR” musculoskeletal<br>manipulations”[Title/Abstract] OR “Manual”[Title/Abstract] OR “adrenal<br>cortex hormones”[Title/Abstract] OR “Corticosteroid”[Title/Abstract] OR<br>“Injections”[Title/Abstract] OR “Lasers”[Title/Abstract] OR<br>“Ultrasonography”[Title/Abstract] OR “Ultrasonics”[Title/Abstract] OR<br>“Extracorporeal”[Title/Abstract] OR “Shock”[Title/Abstract] OR<br>“Wave”[Title/Abstract] OR “Pulsed”[Title/Abstract] OR “electromagnetic<br>radiation”[Title/Abstract] OR “electrical stimulation<br>therapy”[Title/Abstract] OR<br>“Taping therapy”[Title/Abstract] OR “Extracorporeal shock wave<br>therapy”[Title/Abstract] OR “Medium frequency treatment |
| #6 | instrument”[Title/Abstract] OR “Extracorporeal shock wave<br>therapy”[Title/Abstract] OR “acupuncture and moxibustion |
| #7 | therapy”[Title/Abstract] OR “Special sports training<br>therapy”[Title/Abstract]) |
| #8 | OR “Placebos”[MeSH Terms] OR “Randomized Controlled |
| #9 | Trial”[Publication Type] OR “Clinical Trial”[Publication Type]<br>OR (“Therapeutics”[MeSH Terms] OR “therapy”[MeSH Subheading]) OR<br>“Exercise”[MeSH Terms])<br>#4 OR #5 OR #6 OR #7 |
|  | #1 AND #2 AND #3 AND #8 |

### Study selection

All retrieved records were imported into EndNote 20 for duplicate removal. Two reviewers independently screened titles and abstracts and then assessed the full texts of potentially eligible studies. Any disagreements were resolved through discussion, with a third reviewer consulted when necessary.

### Data extraction

Two reviewers independently extracted data using a standardized form. Extracted items included first author, publication year, sample size, participant characteristics, athletic level, diagnostic information, intervention details, comparator details, treatment duration, follow-up duration, outcome measures, and numerical data required for quantitative synthesis. Any discrepancies were resolved by consensus.

### Interventions, comparisons and outcomes

The interventions of interest included exercise therapy, manual therapy, acupuncture, Kinesio taping, extracorporeal shock wave therapy, medium-frequency therapy, microwave diathermy, electrical stimulation therapy, special sports training, and surgical treatment where applicable. Generic intervention terms were used throughout the manuscript. If a proprietary device or brand name was explicitly reported in an included study and was methodologically relevant, it was retained in parentheses at first mention. The intervention domains and comparison framework are summarized in Table 3.

### Risk of bias assessment

The methodological quality of the included randomized controlled trials was independently assessed by two reviewers using the Cochrane risk-of-bias tool. The following domains were evaluated: random sequence generation, allocation concealment, blinding of participants and personnel, blinding of outcome assessment, incomplete outcome data, selective reporting, and other potential sources of bias. Disagreements were resolved through discussion.

### Statistical analysis

A frequentist network meta-analysis was performed using Stata 16.0. Continuous outcomes were summarized as mean differences (MDs) with 95% confidence intervals (CIs). Depending on data availability, either post-intervention values or change scores were extracted, and corresponding standard deviations were used or calculated where possible.

Network plots were generated to illustrate the geometry of the available evidence. Treatment ranking probabilities were estimated using the surface under the cumulative ranking curve (SUCRA). Statistical heterogeneity was assessed within each outcome network, and consistency between direct and indirect evidence was examined using loop-based inconsistency testing where applicable. Sensitivity analyses were conducted when sufficient data were available, and comparison-adjusted funnel plots were used to explore possible small-study effects. The overall analytical workflow is summarized in Table 5.

**Table 5.** Network group command.

| Command | Data format | Graphing |
| --- | --- | --- |
| Network setup and convert |  |  |
| Network map | Study-level data | Network evidence plot |
| Net weight | Study-level data | Contribution plot |
| If plot | Study-level data | Loop consistency plot |
| Net funnel | Study-level data | Comparison-adjusted plot |
| Network convert augment<br>Network meta i | Arm-based data |  |
| Network forest | After mvmeta | Network forest plot |
| Net league | After mvmeta | Consistency model inverted triangle plot<br>(pairwise comparison of core results) |
| Interval plot | After mvmeta | Prediction interval graph |
| Surface under the<br>cumulative ranking curve | After network<br>rank | Surface under the cumulative ranking<br>plot |

### Power calculation

A formal a priori power calculation was not performed because this study was a secondary analysis of previously published randomized controlled trials rather than a primary data-collection study.

### Ethics statement

Ethical approval was not required because this study was a systematic review and network meta-analysis based exclusively on previously published data.

## Results

### Eligible studies

A total of 1,223 records were identified through the database search, including 1,168 records in English and 55 records in Chinese. After removal of 329 duplicates, 894 records remained for title and abstract screening. Following exclusion of 598 records, 296 full-text articles were assessed for eligibility. Finally, 45 studies were included in the qualitative synthesis, and 42 studies contributed to the quantitative network meta-analysis. The study-selection process is shown in Fig 1, whereas the characteristics of the included studies and the quality assessment results are summarized in Tables 6 and 7.

**Fig 1.**
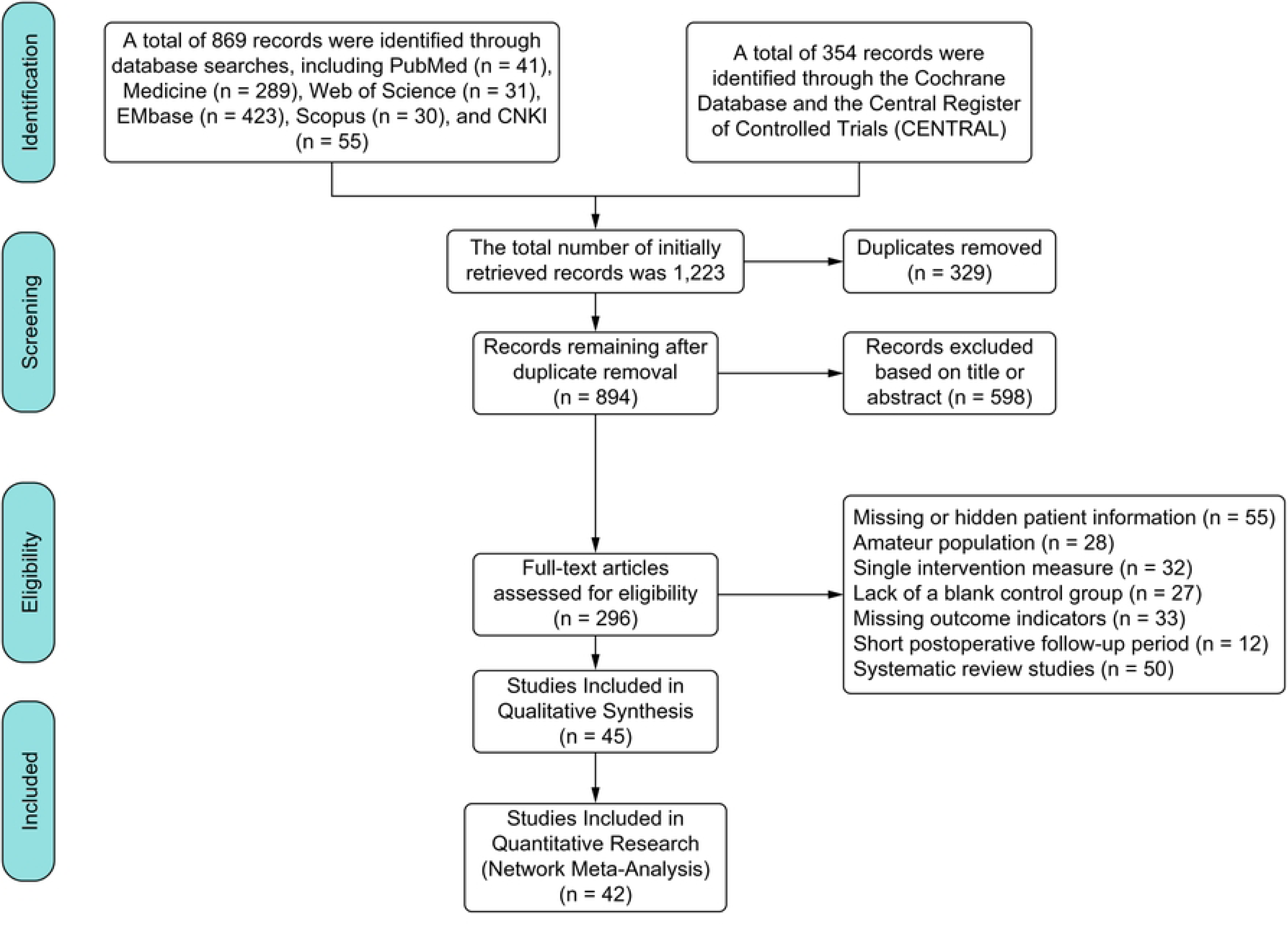
PRISMA flow chart of the study selection process.

**Table 6.**
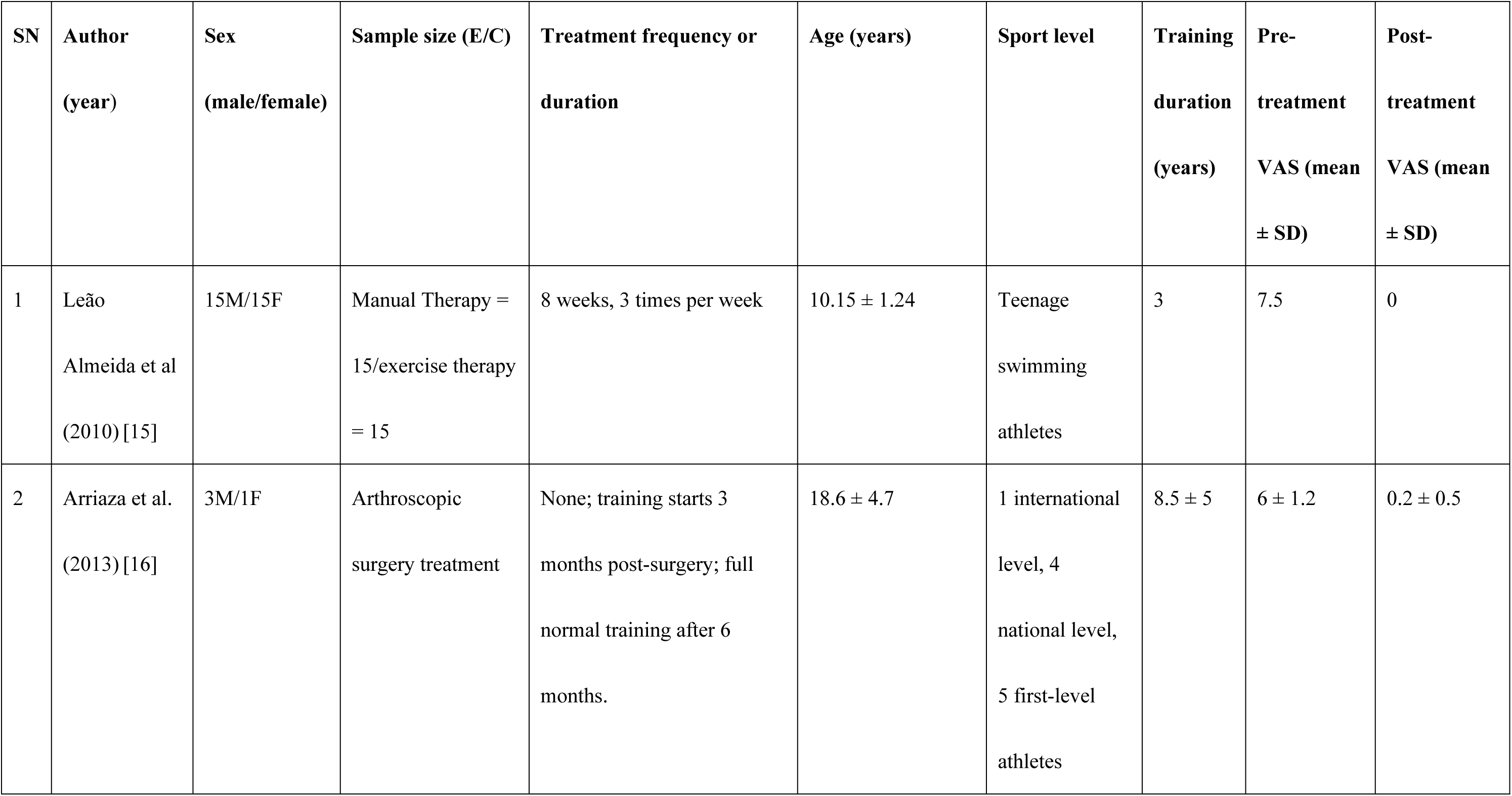
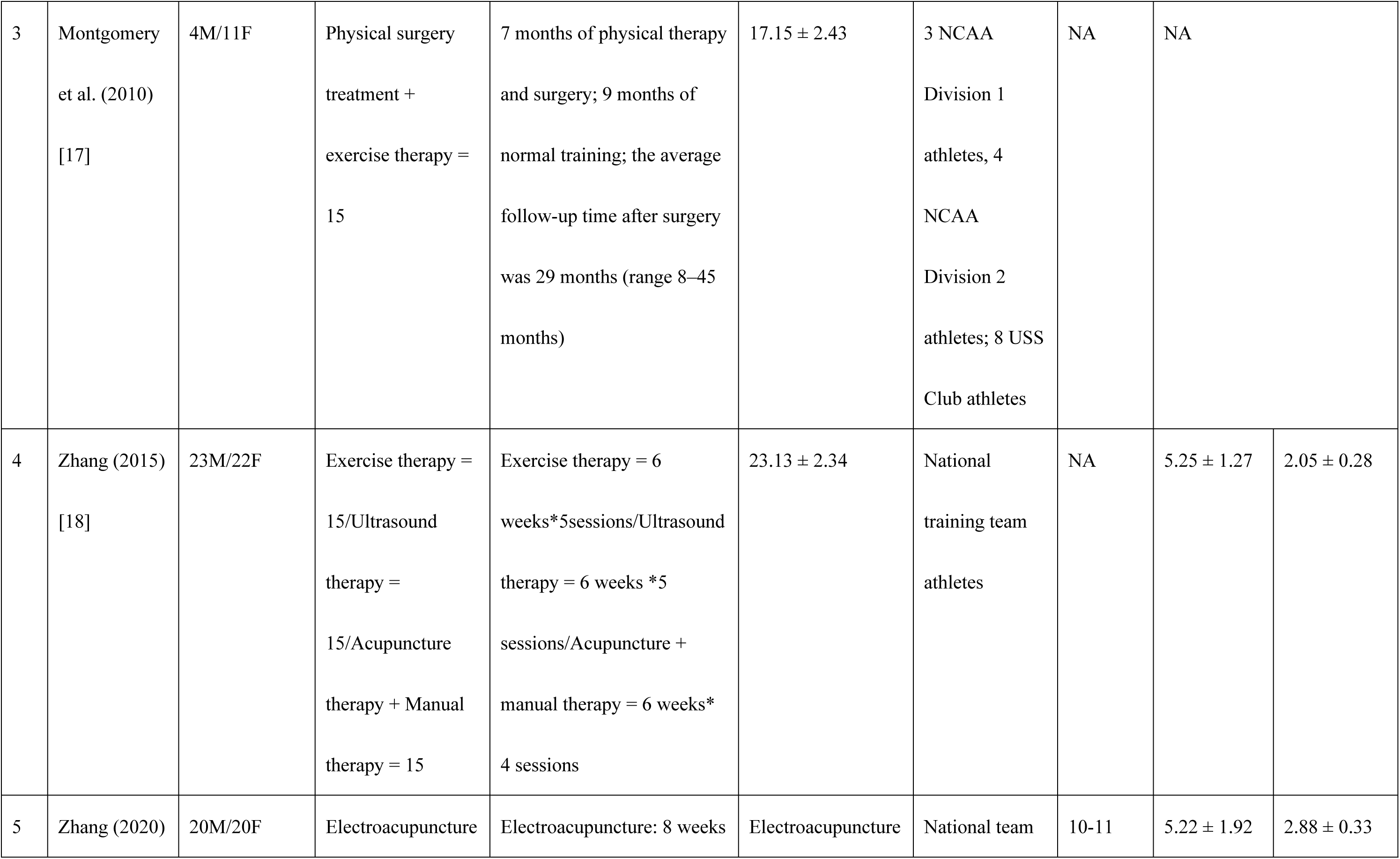
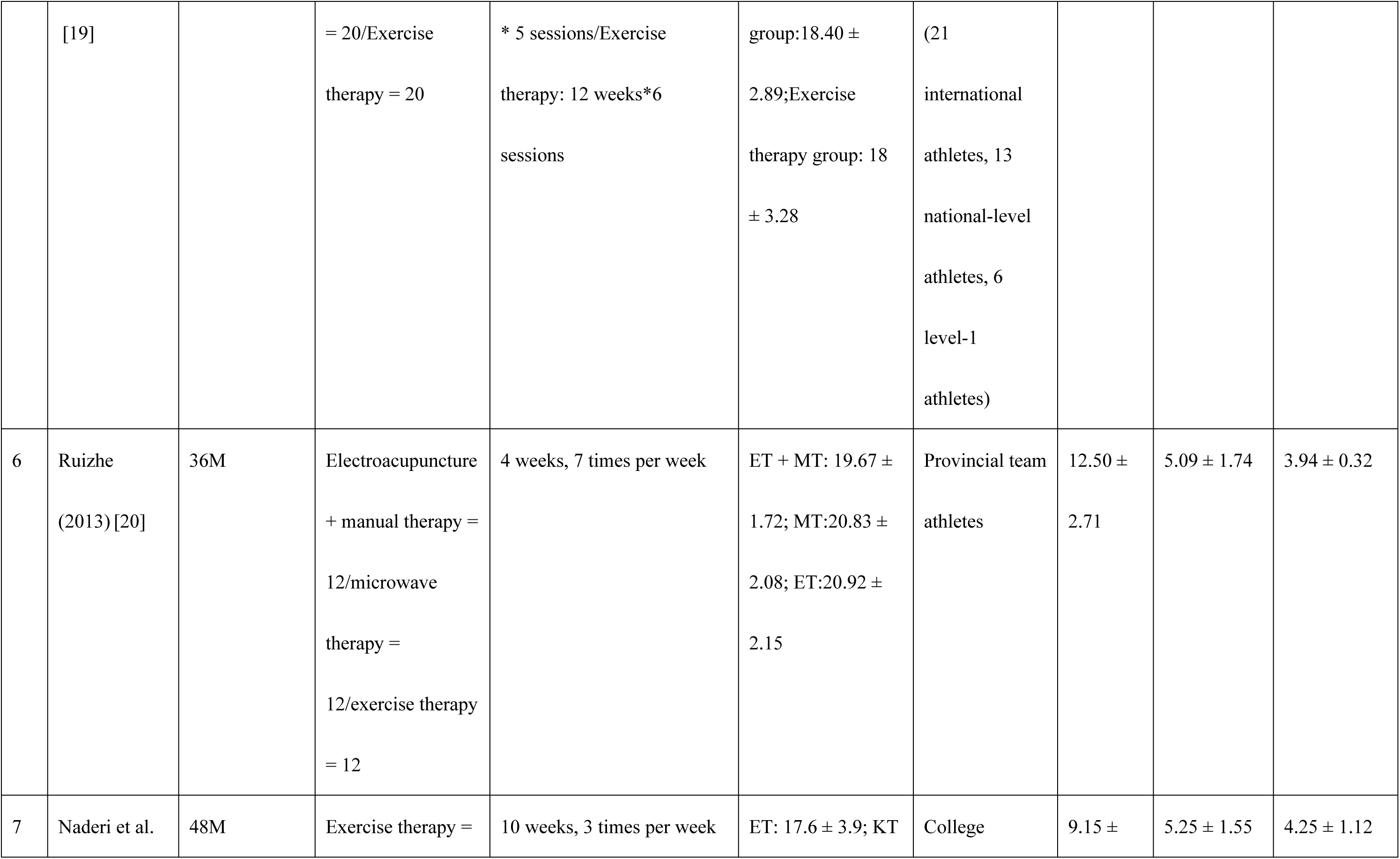
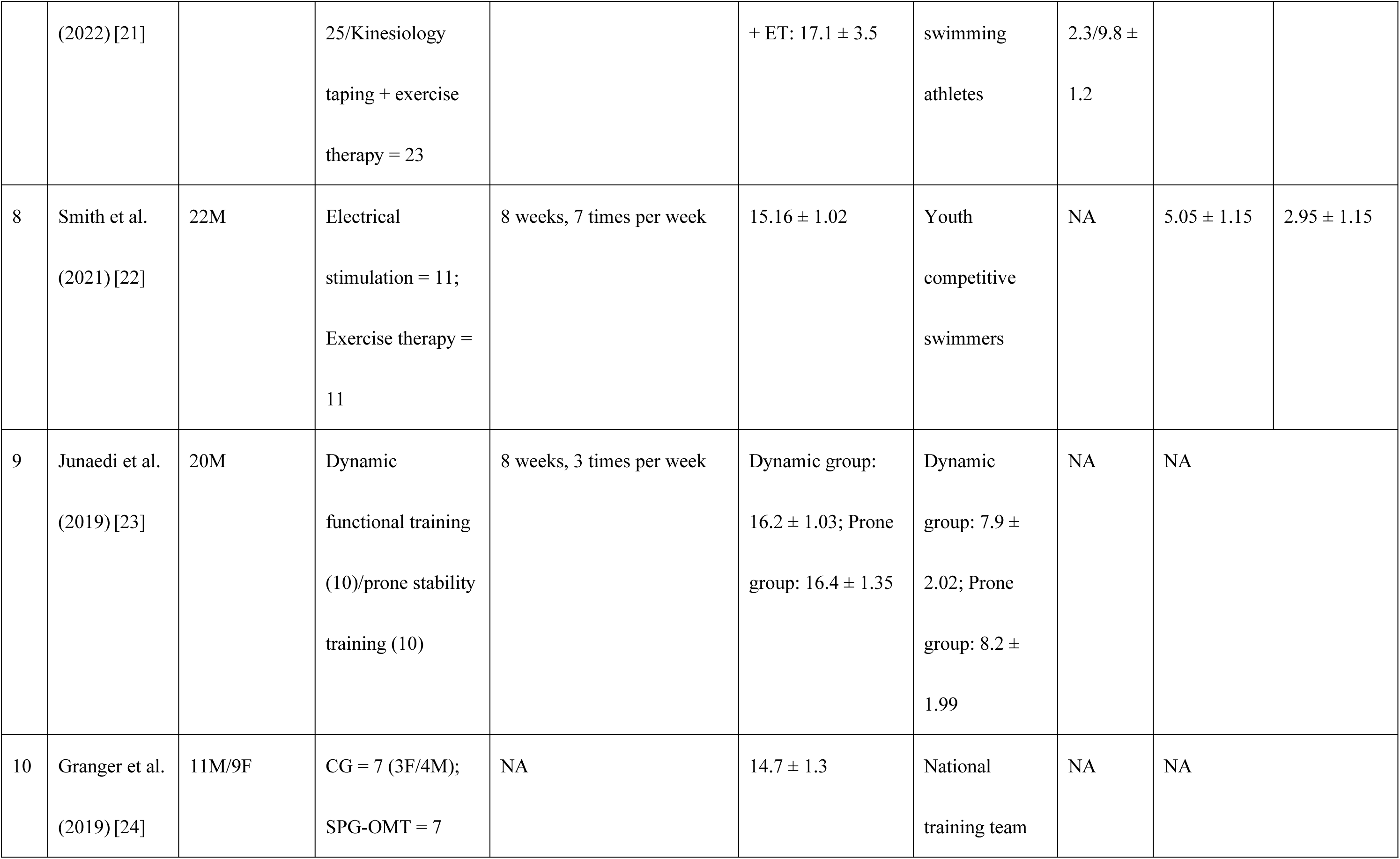
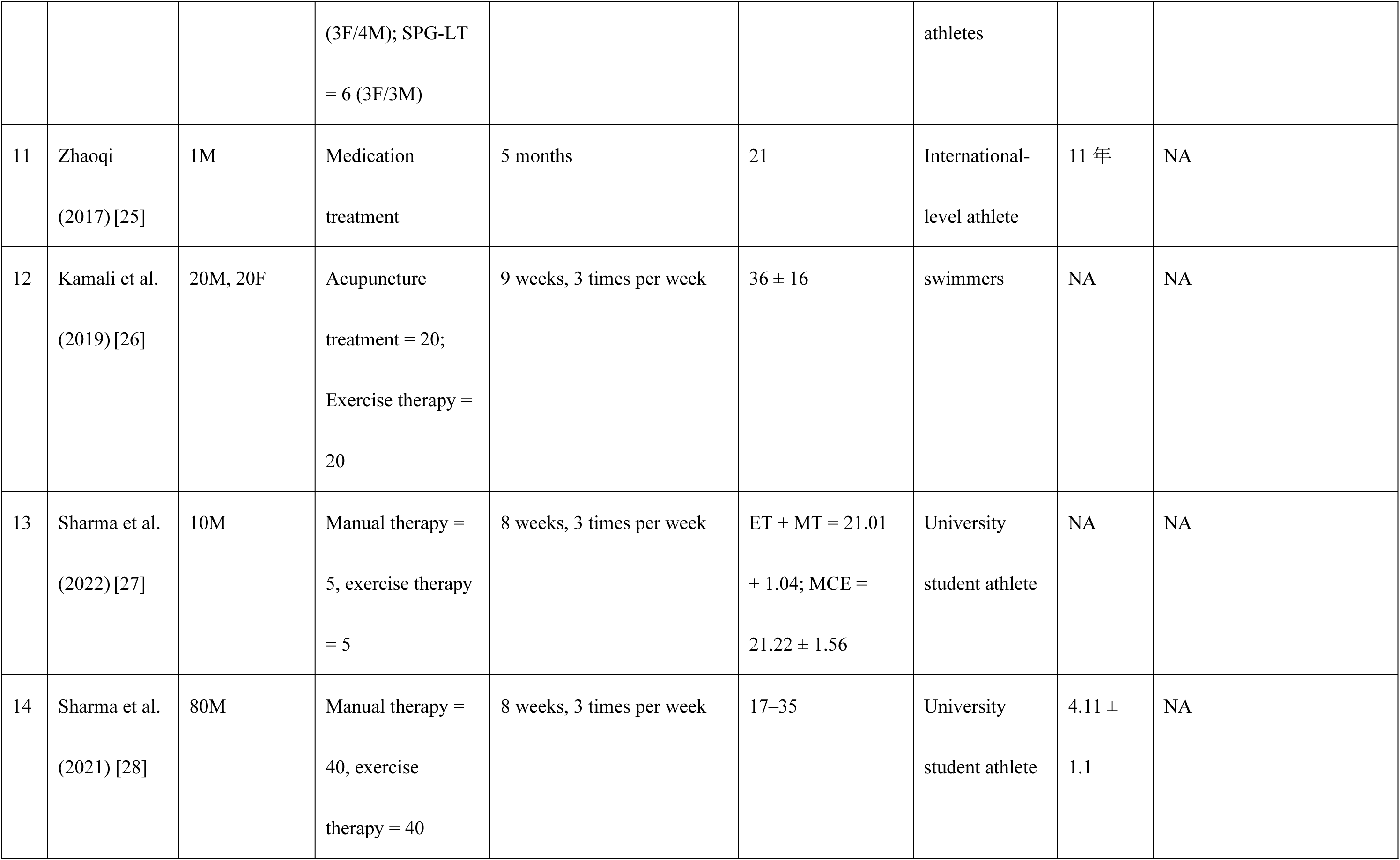
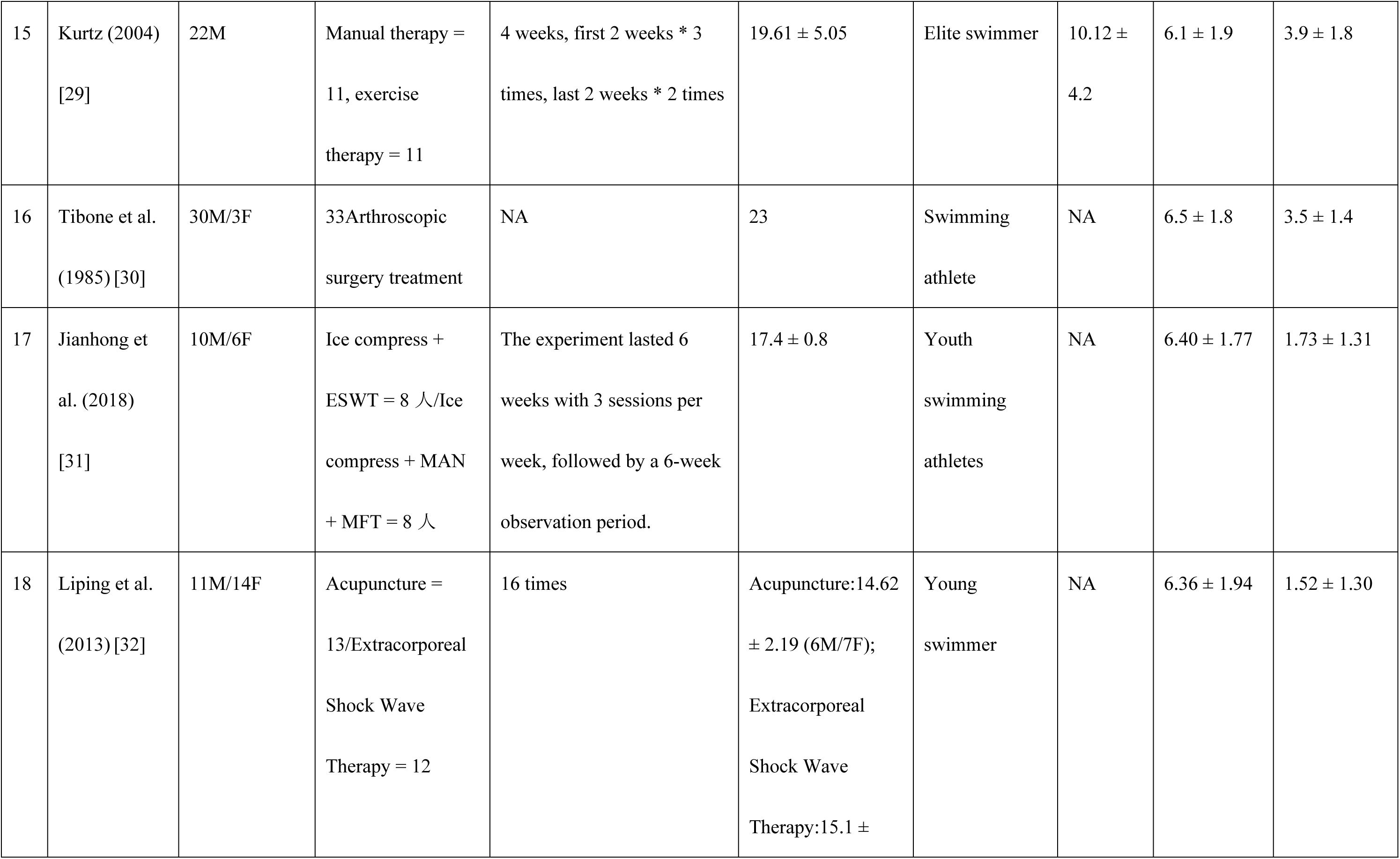
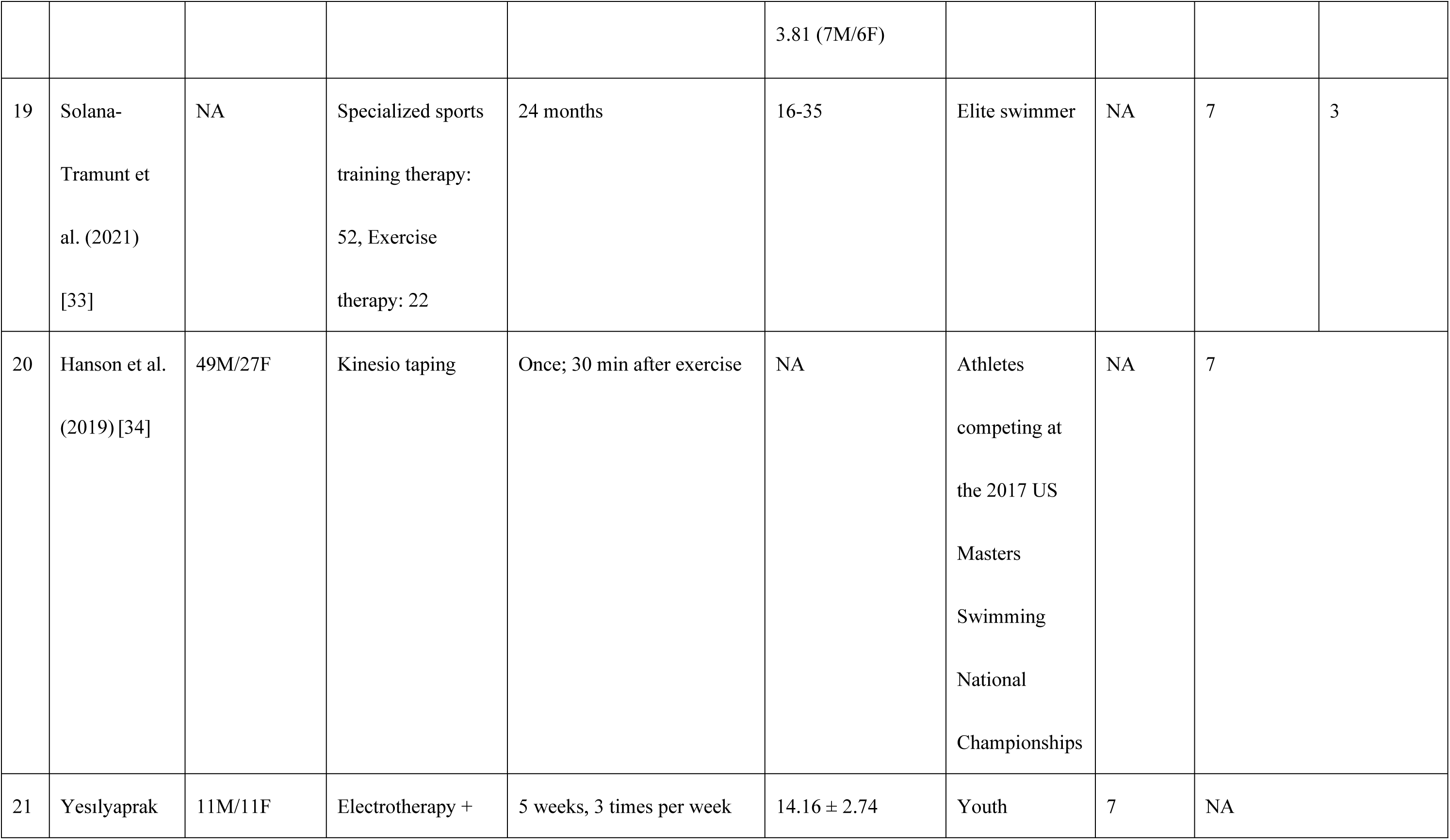
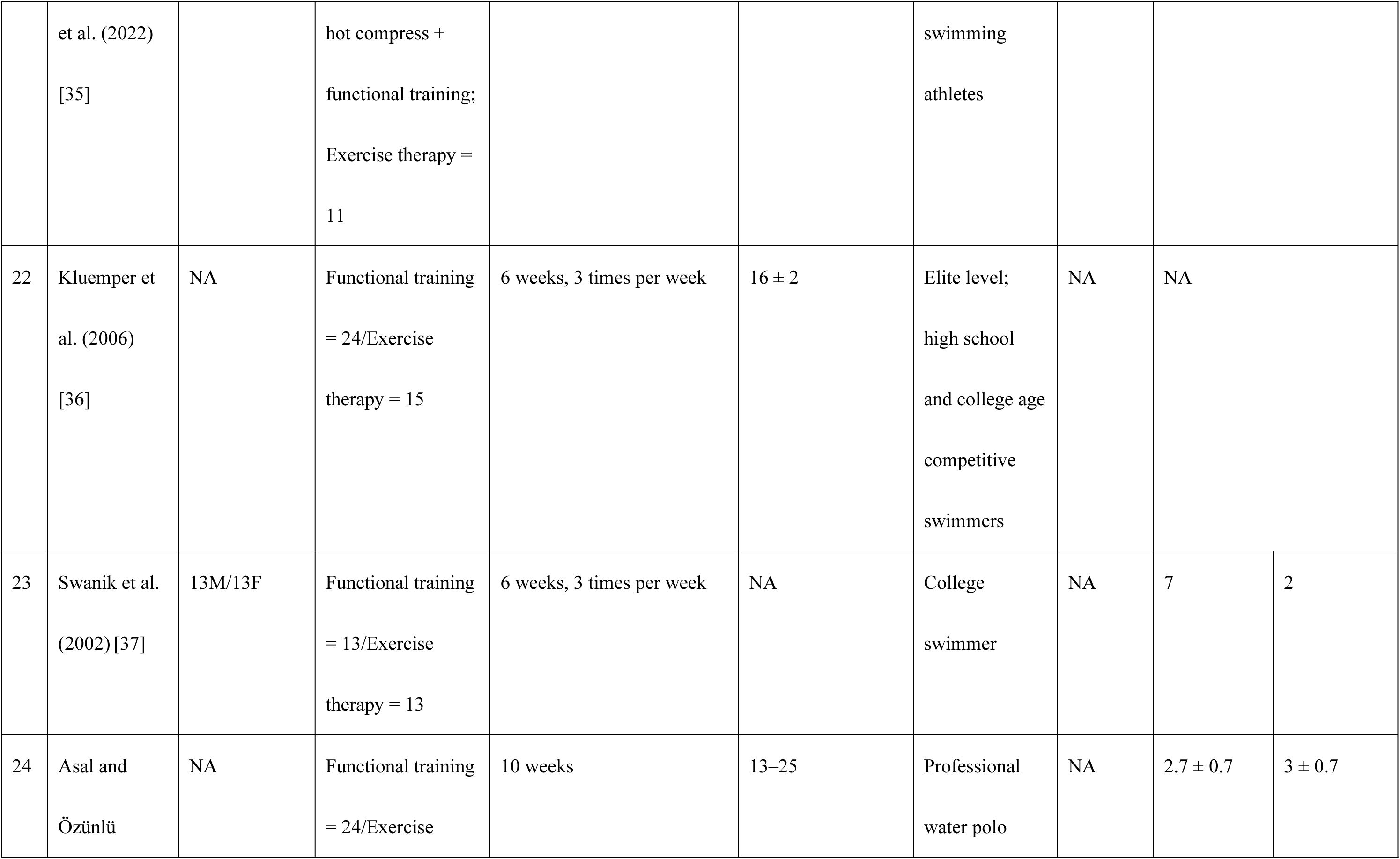
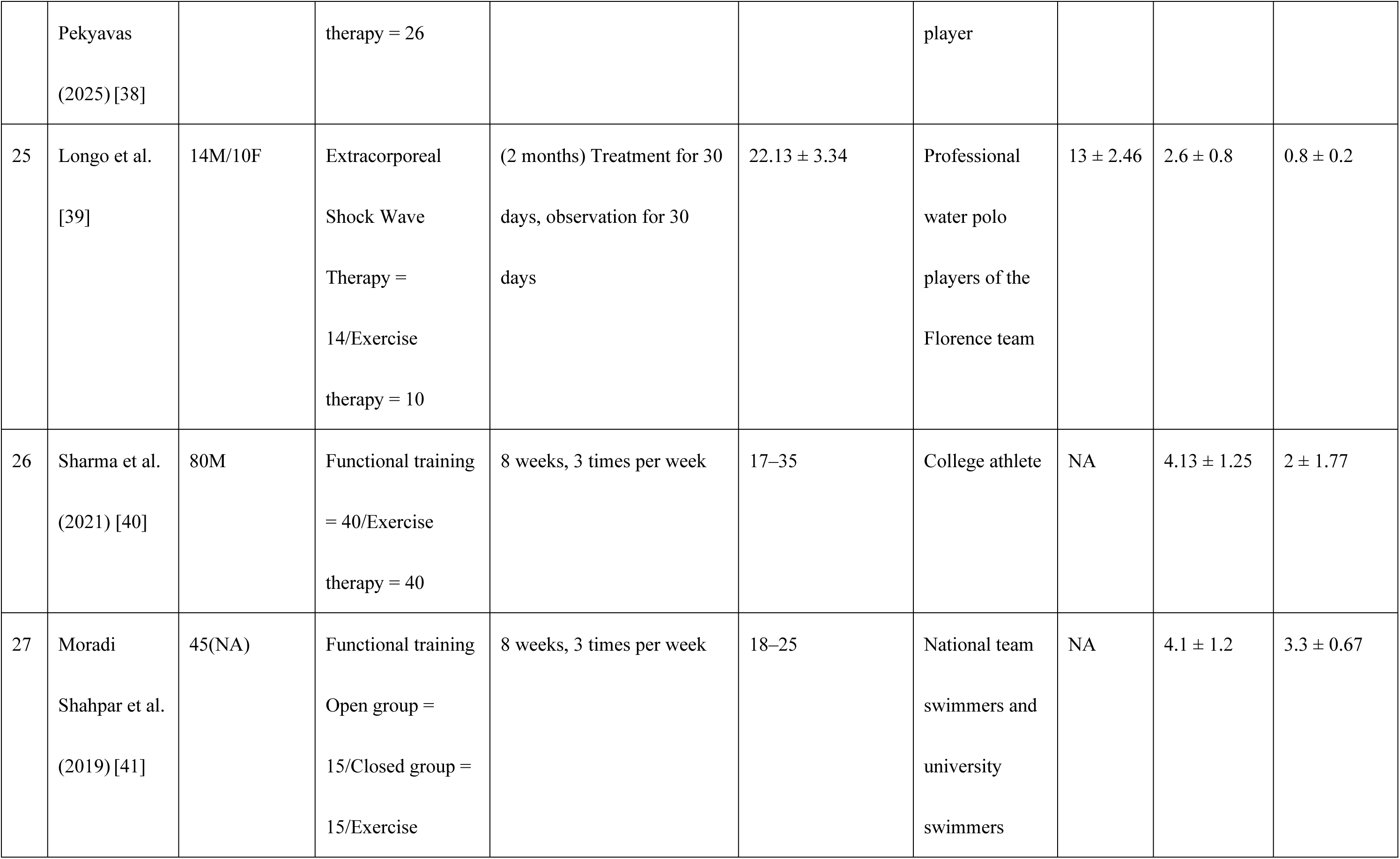
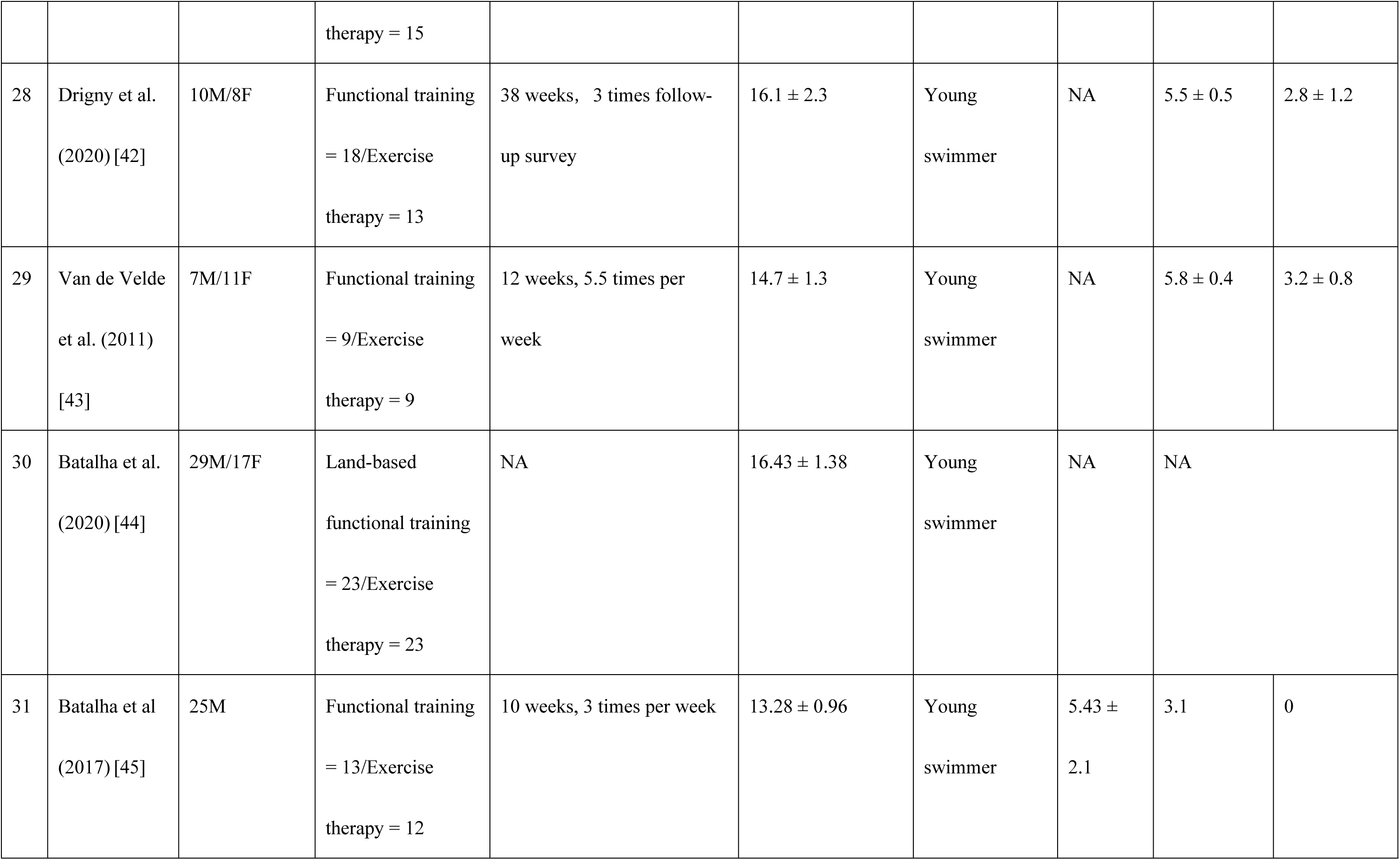
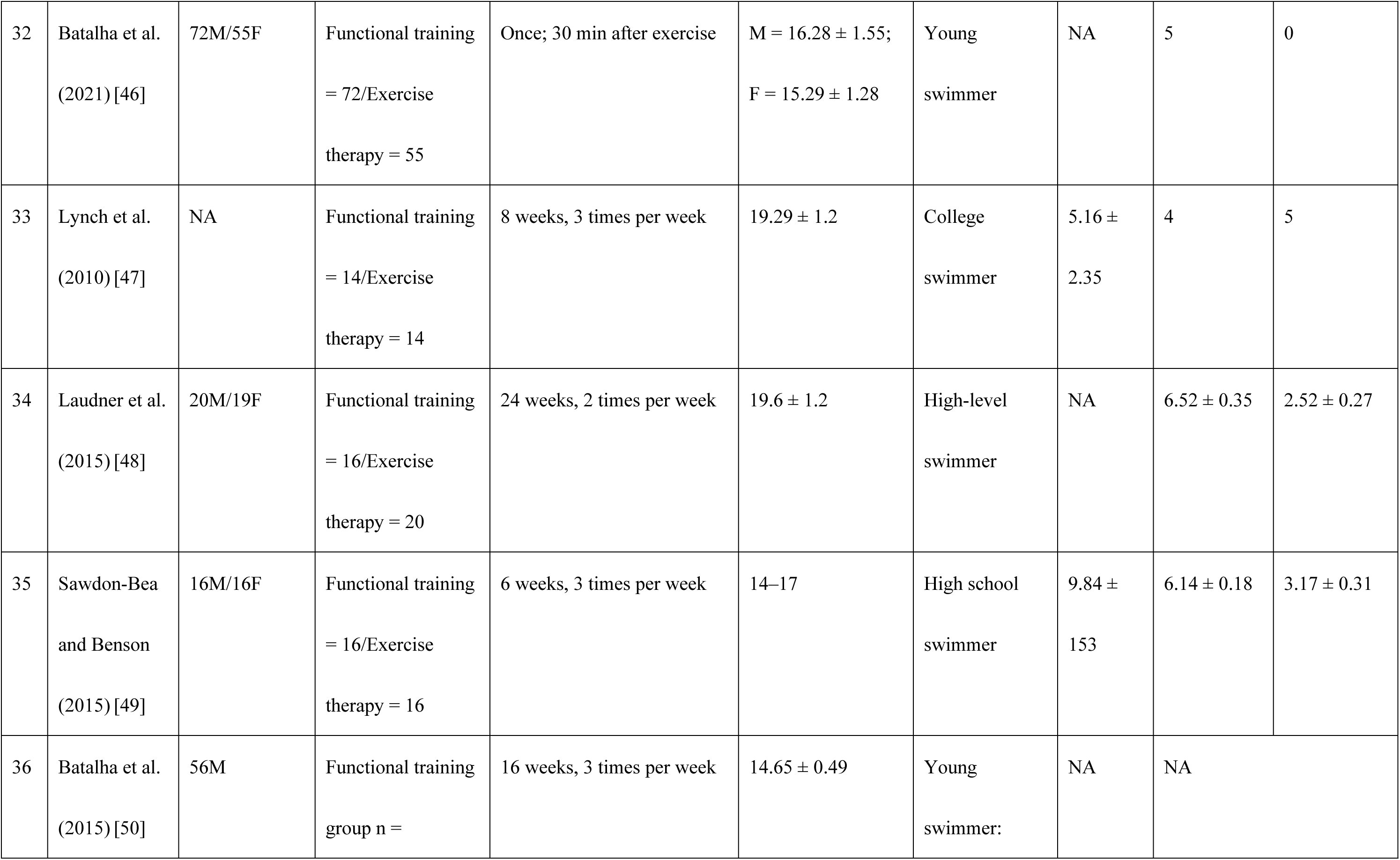
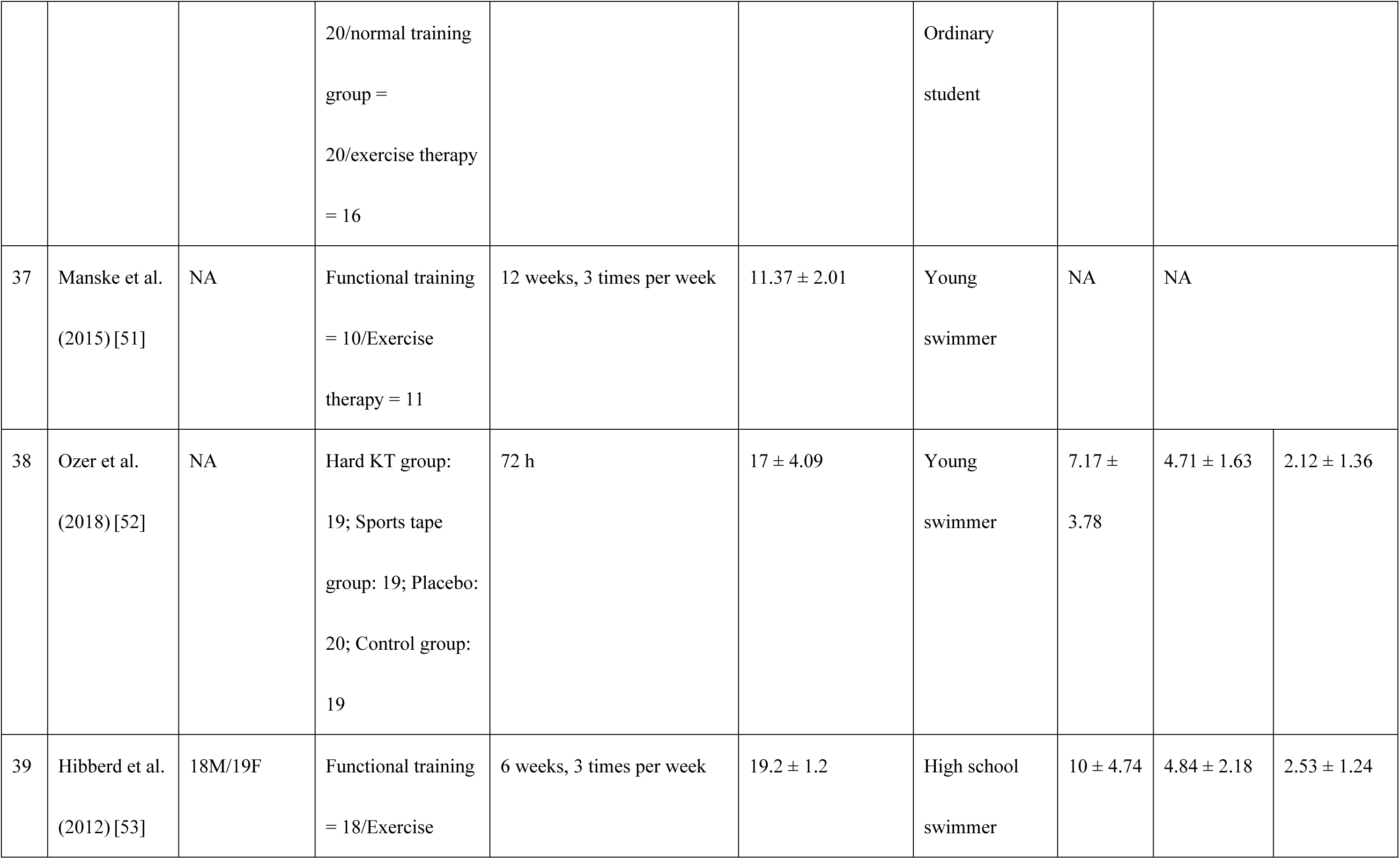
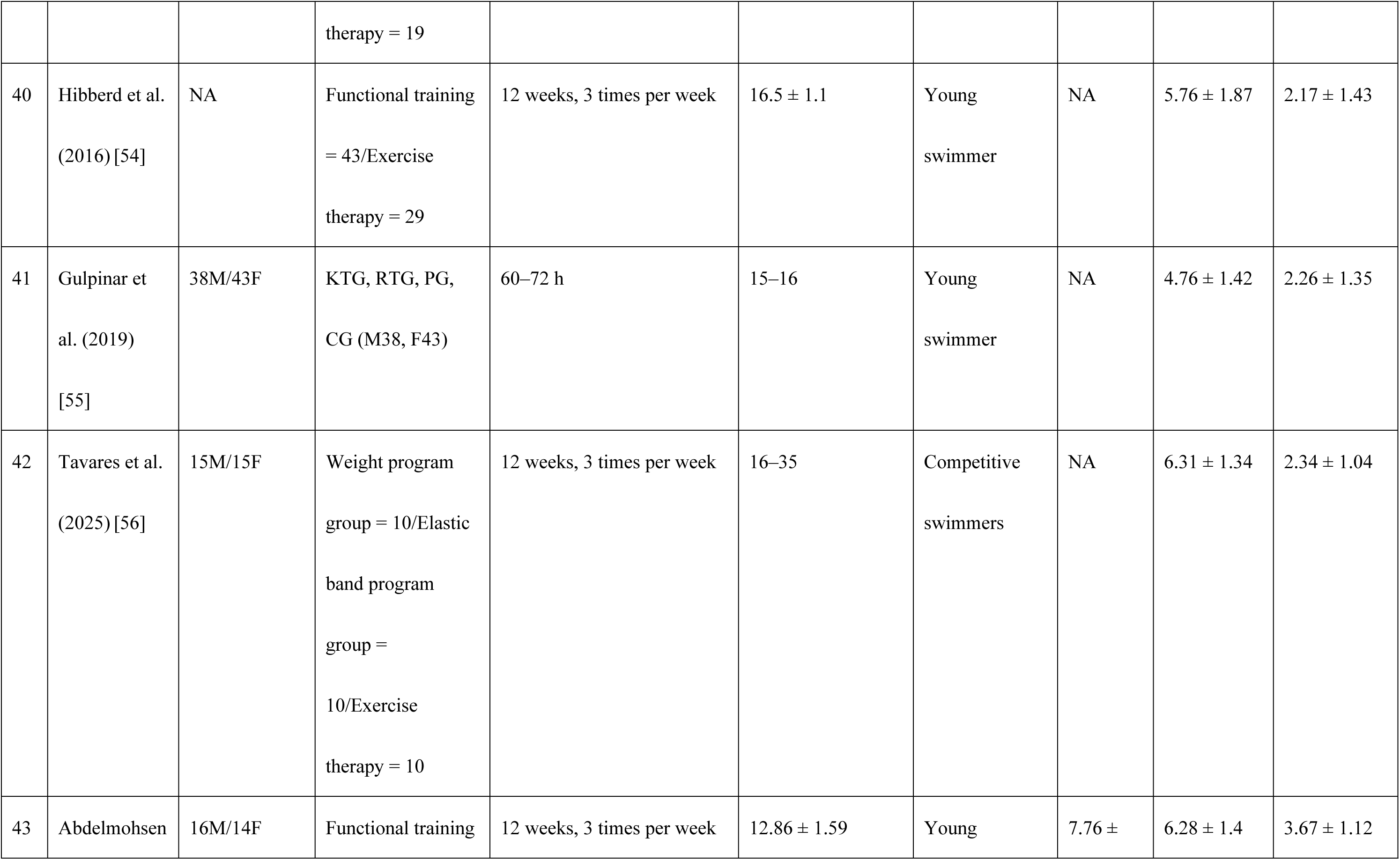
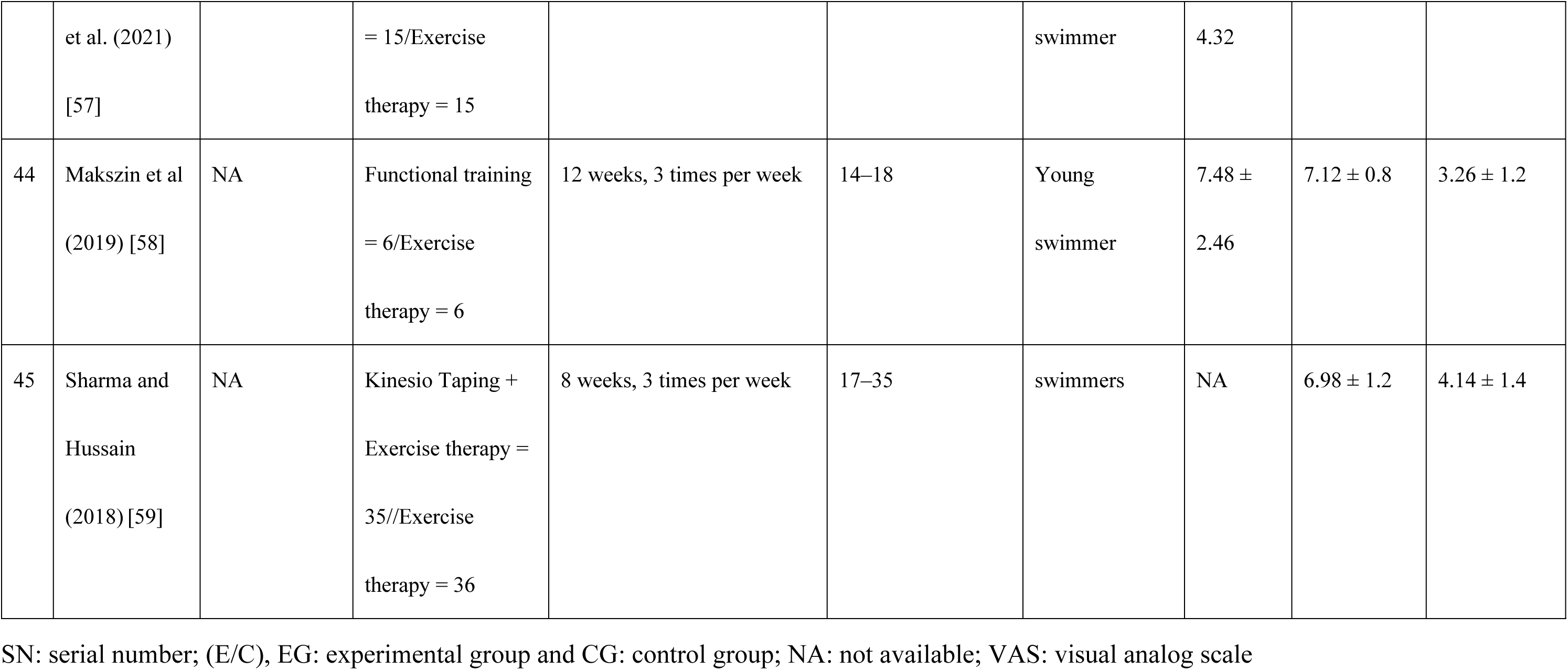
Characteristics of selected articles and included participants.

**Table 7.** Quality evaluation of the included articles.

| SN | Author<br>(Year) | Random<br>sequence | Allocation<br>concealment | Blinding of<br>participants | Blinding of<br>outcome | Incomplete<br>outcome | Selective<br>outcome | Other<br>risks of |
| --- | --- | --- | --- | --- | --- | --- | --- | --- |

|  |  | **generation** |  | **and**<br>**personnel** | **assessors** |  | **reporting** | **bias** |
| --- | --- | --- | --- | --- | --- | --- | --- | --- |
| 1 | Leão Almeida et al<br>(2010) [15] | Unclear | Unclear | Unclear | Low | Low | Low | Low |
| 2 | Arriaza et al. (2013) [16] | High | Unclear | Unclear | Low | Low | Low | Low |
| 3 | Montgomery et al.<br>(2010) [17] | Unclear | Unclear | Unclear | Low | Low | Low | Low |
| 4 | Zhang (2015) [18] | Low | Low | Low | Low | Low | Low | Low |
| 5 | Zhang et al. (2020) [19] | High | High | Low | Low | Low | Low | Low |
| 6 | Ruizhe (2013) [20] | Low | High | Unclear | Low | Low | Low | Low |
| 7 | Naderi et al. (2022) [21] | Low | Low | Low | Low | Low | Low | Low |
| 8 | Smith et al. (2021) [22] | Unclear | Unclear | Unclear | Low | Low | Low | Low |
| 9 | Junaedi et al. (2019) [23] | Low | Low | Low | Low | Low | Low | Low |
| 10 | Granger et al. (2019)<br>[24] | Unclear | Unclear | Low | Low | Low | Low | Low |
| 11 | Zhaoqi (2017) [25] | Unclear | Unclear | Low | Low | Low | Low | Low |
| 12 | Kamali et al. (2019) [26] | Low | Low | Low | Low | Low | Low | Low |
| 13 | Sharma et al. (2022) [27] | Low | Low | Low | Low | Low | High | Low |
| 14 | Sharma et al. (2021)<br>[28,40] | Low | Low | Low | Low | Low | Low | Low |
| 15 | Kurtz (2004) [29] | Unclear | Unclear | Low | Low | Low | High | Low |
| 16 | Tibone et al. (1985) [30] | Unclear | Low | Low | Low | Low | Low | Low |
| 17 | Jianhong et al. (2018)<br>[31] | Low | Unclear | Unclear | Low | Low | Low | Low |
| 18 | Liping et al. (2013) [32] | Unclear | Low | Unclear | Low | Low | Low | Low |
| 19 | Solana-Tramunt et al.<br>(2021) [33] | Unclear | Unclear | Low | Low | Low | Low | Low |
| 20 | Hanson et al (2018) [34] | High | Low | Low | Low | Low | Low | High |
| 21 | Yesilyaprak et al. (2022) [35] | Unclear | Unclear | Low | Low | Low | Low | Low |
| 22 | Kluemper et al. (2006) [36] | Low | Low | Low | Low | Low | Low | Low |
| 23 | Swanik et al. (2002) [37] | Low | Low | Low | Low | Low | Low | Low |
| 24 | Longo et al. (2022) [39] | Low | Low | Low | Low | Low | Low | Low |
| 25 | Sharma et al. (2021) [28,40] | Low | Low | Low | Low | Low | Low | Low |
| 26 | Moradi Shahpar et al. (2019) [41] | Low | Low | Low | Low | Low | Low | Low |
| 27 | Drigny et al. (2020) [42] | Unclear | Low | Low | Low | Low | Low | Low |
| 28 | Van de Velde et al. (2011) [43] | Low | Low | Low | Low | Low | Low | Low |
| 29 | Batalha et al. (2020) [44] | High | High | Low | Low | Low | Low | Low |
| 30 | Batalha et al (2017) [45] | Low | Low | Low | Low | Low | Low | Low |
| 31 | Batalha et al. (2021) [46] | High | Unclear | Low | Low | Low | Low | Low |
| 32 | Lynch et al. (2010) [47] | Low | Low | Low | Low | Low | Low | Low |
| 33 | Laudner et al. (2015)<br>[48] | Unclear | Unclear | Low | Low | Low | Low | Low |
| 34 | Sawdon-Bea and Benson<br>(2015) [49] | High | Unclear | Low | Low | Low | Low | Low |
| 35 | Batalha et al. (2015) [50] | Unclear | Unclear | Low | Low | Low | Low | Low |
| 36 | Manske et al. (2015) [51] | Unclear | Unclear | Low | Low | Low | Low | Low |
| 37 | Ozer et al. (2018) [52] | Low | Low | Low | Low | Low | Low | Low |
| 38 | Hibberd et al. (2012)<br>[53] | Low | Unclear | Low | Low | Low | Low | Low |
| 39 | Hibberd et al. (2016)<br>[54] | High | Unclear | Low | Low | Low | Low | High |
| 40 | Gulpinar et al. (2017)<br>[55] | Low | Low | Low | Low | Low | Low | Low |
| 41 | Asal and Özünlü<br>Pekyavas (2025) [39] | Low | Low | Low | Low | Low | Low | Low |
| 42 | Tavares et al. (2025) [56] | Low | Low | Low | Low | Low | Low | Low |
| 43 | Abdelmohsen et al<br>(2022) [57] | High | Low | Low | Low | Low | Low | Low |
| 44 | Makszin et al (2019) [58] | High | Low | Low | Low | Low | Low | Low |
| 45 | Sharma and Hussain<br>(2018) [59] | Low | Low | Low | Low | Low | Low | Low |

S/N: serial number

### Risk of bias

Risk of bias was assessed across all 45 included studies, and the results are summarized in Figs 2 and 3 and detailed in Table 7. For random sequence generation, 23 studies were judged as low risk, 16 as unclear risk, and 6 as high risk. For allocation concealment, 25 studies were rated as low risk, 17 as unclear risk, and 3 as high risk. Blinding of participants and personnel represented the domain with the highest overall risk, whereas outcome assessment, incomplete outcome data, and selective reporting were generally rated more favorably. Overall, the evidence base was limited mainly by the relatively high risk of performance bias in a substantial proportion of studies.

**Fig 2.**
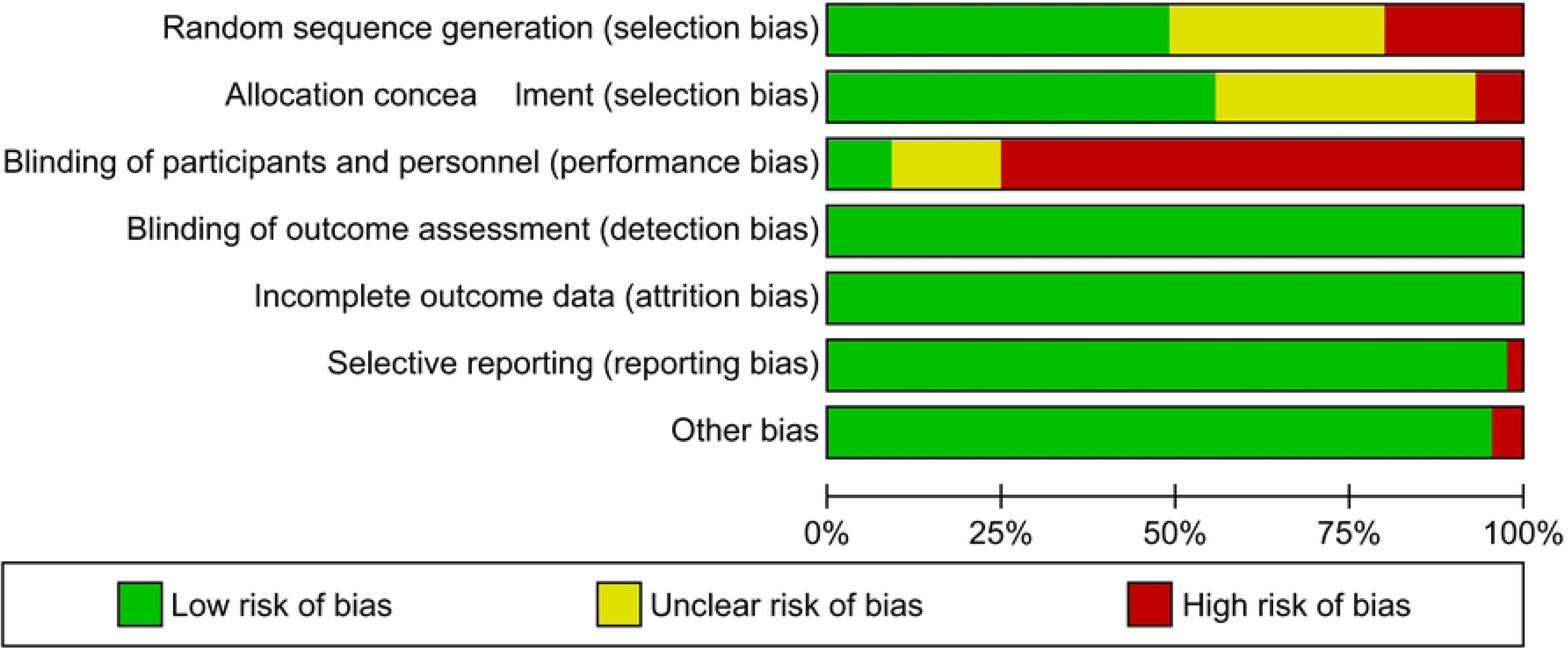
Risk of bias graph.

**Fig 3.**
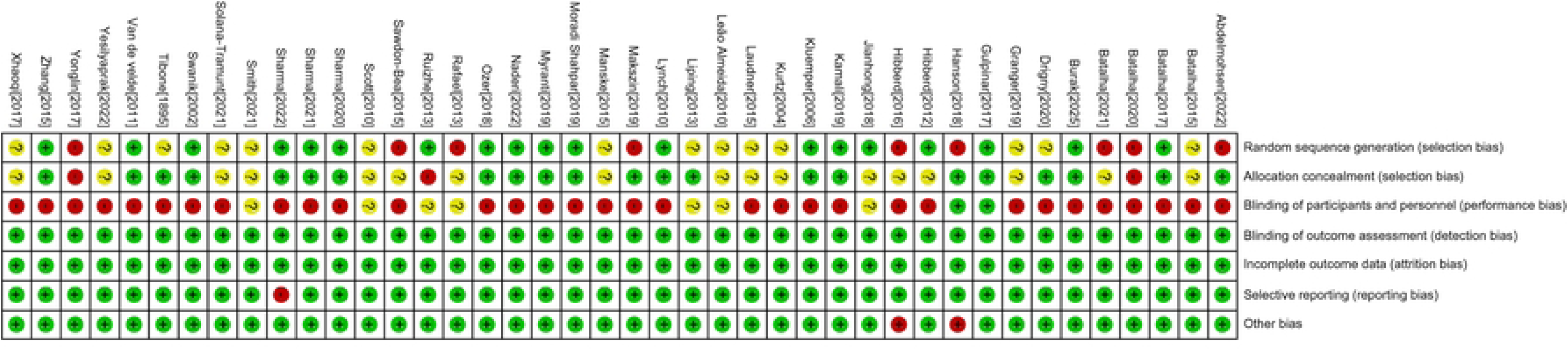
Risk of bias summary.

### Network geometry and contribution

The treatment network comprised 9 intervention modalities and 13 outcome measures, yielding 81 direct and 99 indirect comparisons. Exercise therapy occupied a central position in the network and served as the most frequent comparator across studies. Among the reported outcomes, the visual analog scale (VAS) was included in 17 studies, whereas internal rotation (IR) and external rotation (ER) were assessed in 25 and 23 studies, respectively. The greatest contribution of direct evidence was observed for comparisons between exercise therapy and special sports training, followed by comparisons of exercise therapy with acupuncture and manual therapy. The strongest indirect contributions were observed between acupuncture and manual therapy and between medium-frequency therapy and microwave diathermy. The network geometry and contribution of available evidence are shown in Figs 4 and 5.

**Fig 4.**
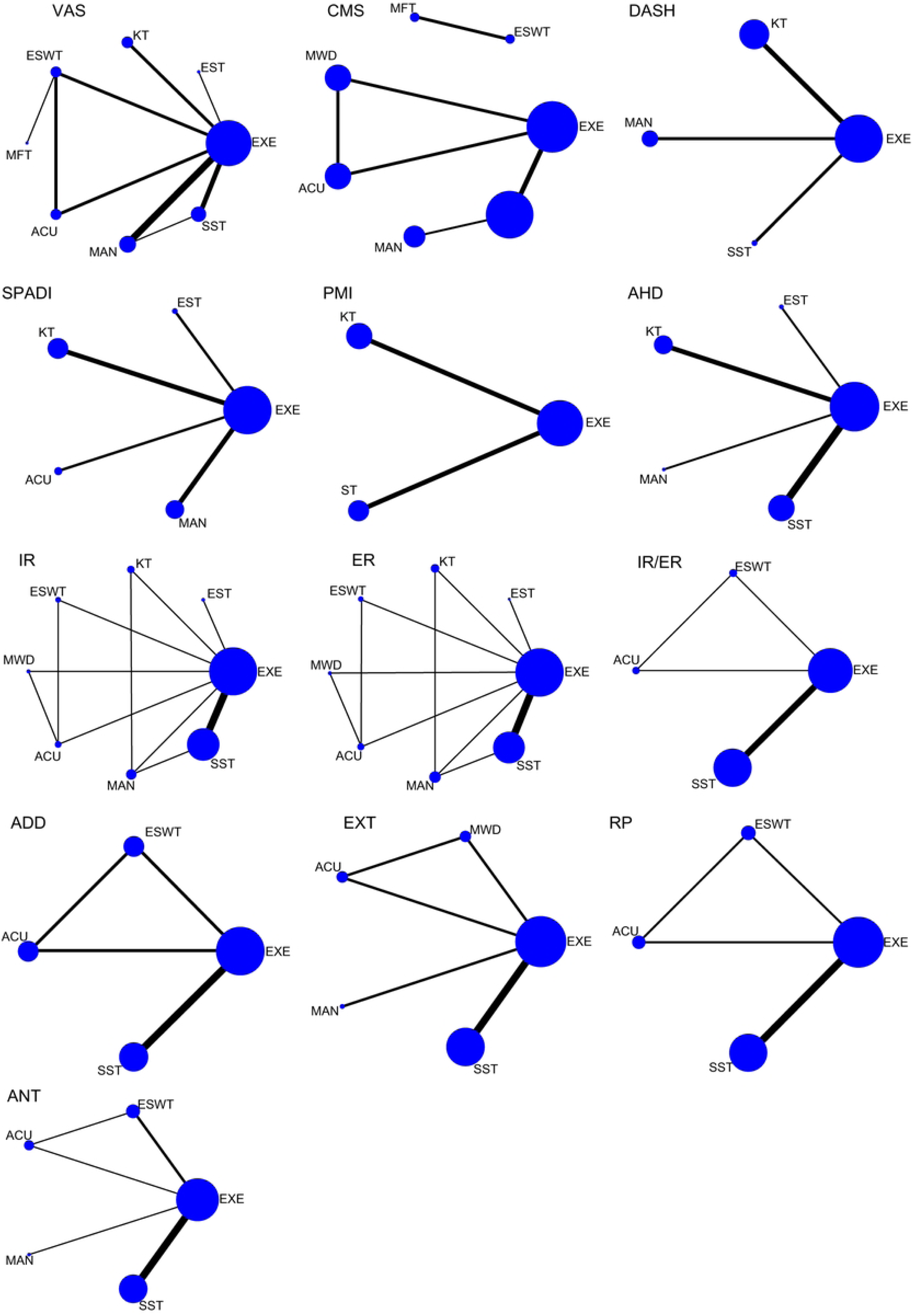
Network evidence map of the therapeutic effects of different treatment methods on swimmers with SIS. ACU: acupuncture; ANT: antexion; AHD: acromiohumeral distance; ADD: adduction; CMS: Constant–Murley score; DASH: Disabilities of the Arm, Shoulder, and Hand questionnaire; EST: electrical stimulation therapy; EXT: extended; EXE: exercise therapy; ER: external rotation; IR: internal rotation; KT: Kinesio taping; MAN: manual therapy; MFT: medium-frequency therapy apparatus; MWD: microwave diathermy; PMI: pectoralis minor index; ESWT: extracorporeal shock wave therapy; SPADI: Shoulder Pain and Disability Index; SST: special sports training therapy; SIS: shoulder impingement syndrome; VAS: visual analog scale.

**Fig 5.**
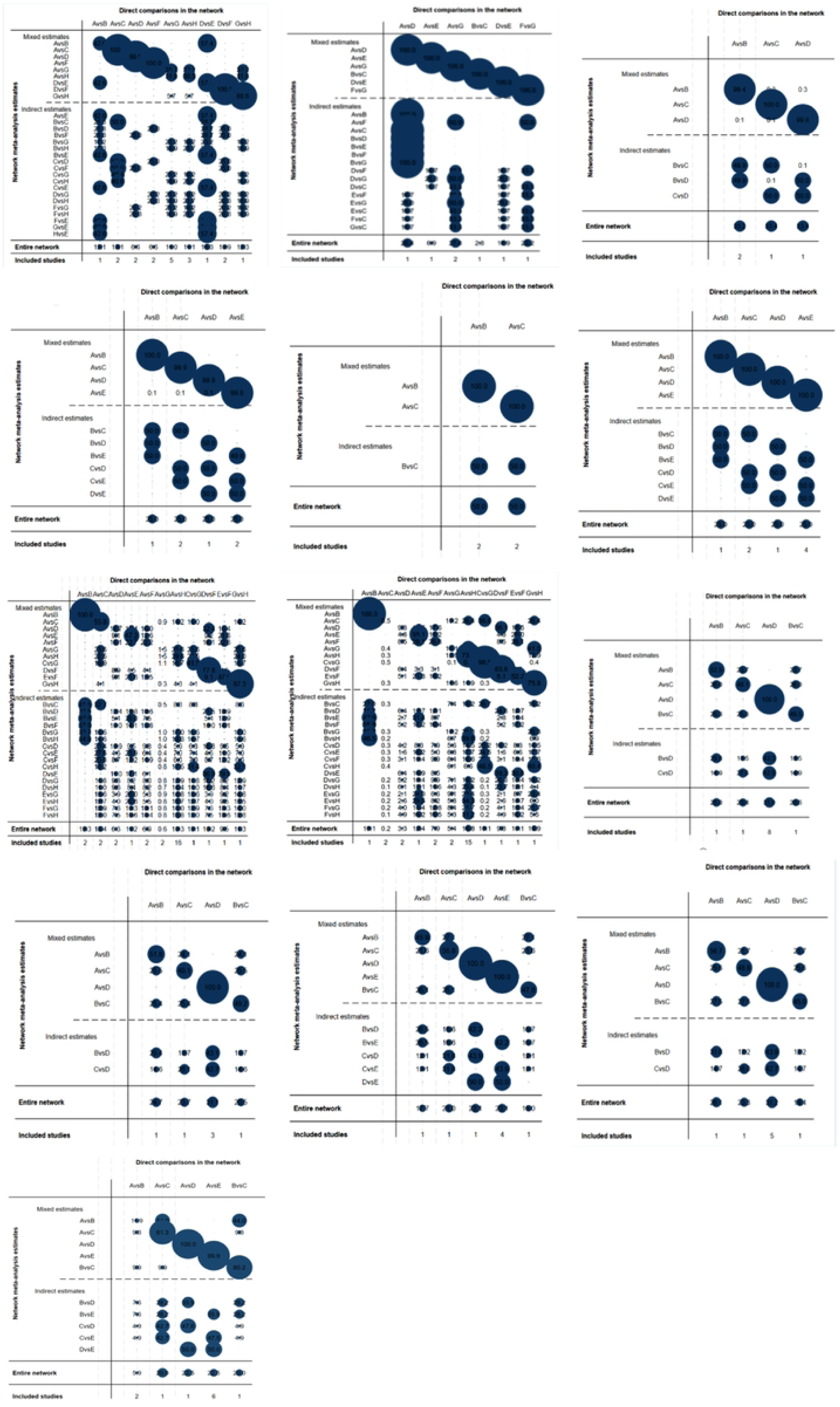
Network contribution map of the therapeutic effects of different treatment methods on swimmers with SIS. SIS: shoulder impingement syndrome.

### Inconsistency and heterogeneity

Loop-specific inconsistency analyses were performed for the VAS, shoulder flexion, ER, and IR networks (Figs 6–9). In the VAS network (Fig 6), two closed loops were identified. The EXE–ESWT–MFT loop showed evidence of inconsistency (inconsistency factor [IF] = 3.16, 95% CI 2.12 to 4.19), whereas the EXE–ACU–MAN loop had a CI that included 0 (IF = 2.65, 95% CI 0.00 to 9.82). In the shoulder flexion network (Fig 7), the EXE–ESWT–ACU loop also suggested inconsistency (IF = 4.62, 95% CI 1.59 to 7.65). In the ER network (Fig 8), inconsistency was present in the EXE–ESWT–ACU loop (IF = 6.27, 95% CI 2.36 to 10.18) and the EXE–KT–MAN loop (IF = 4.89, 95% CI 3.13 to 6.66), whereas the EXE–MWD–ACU and EXE–MAN–SST loops had CIs that included 0. A similar pattern was observed in the IR network (Fig 9), with inconsistency detected in the EXE–ESWT–ACU loop (IF = 4.74, 95% CI 2.45 to 7.03) and the EXE–KT–MAN loop (IF = 4.52, 95% CI 2.30 to 6.74), whereas the EXE–MWD–ACU and EXE–MAN–SST loops had CIs that included 0. Overall, inconsistency appeared to be localized to selected loops rather than affecting the entire network.

**Fig 6.**
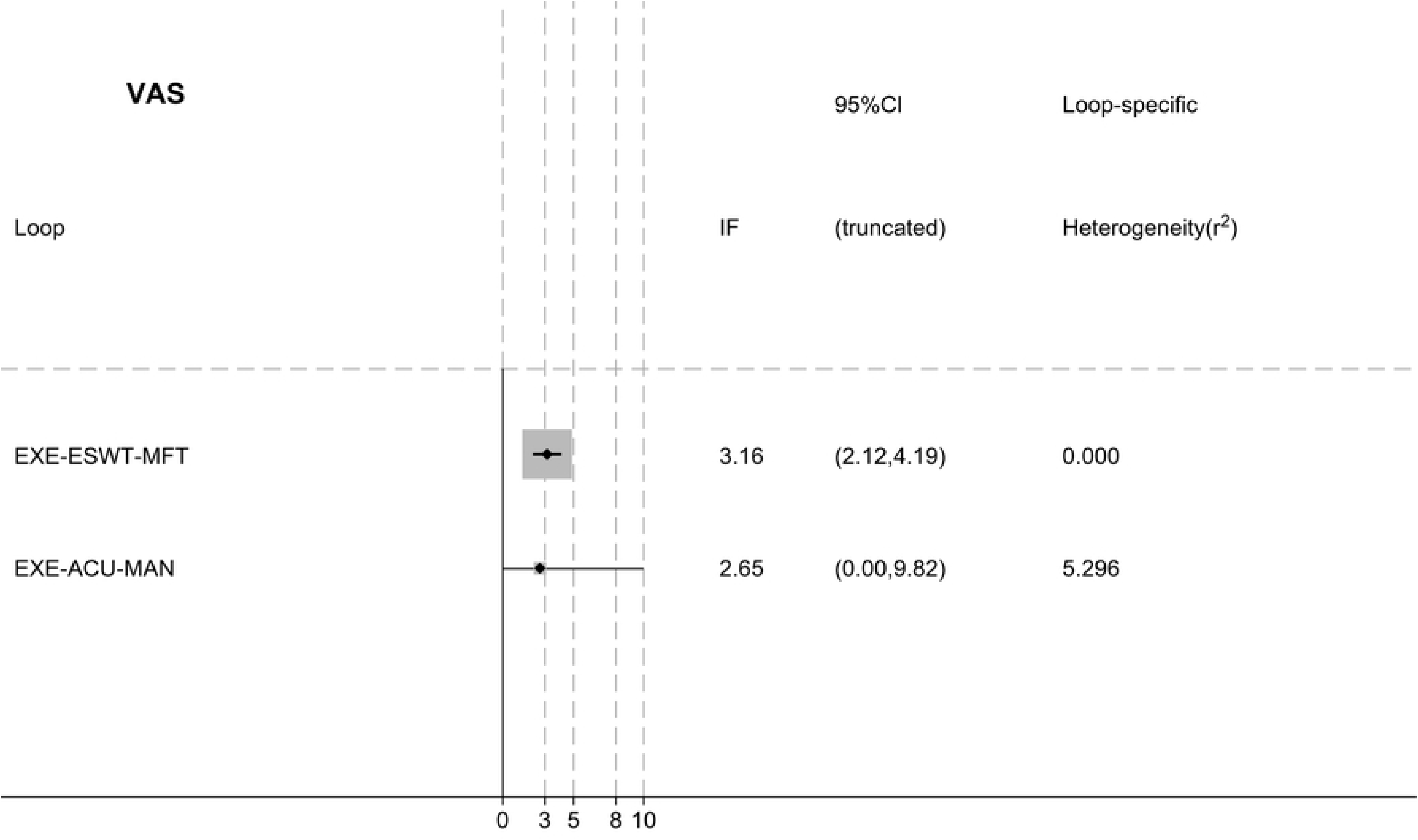
Visual analog scale inconsistency test. ACU: acupuncture; CI: confidence interval; EXE: exercise therapy; MAN: manual therapy; MFT: medium-frequency therapy apparatus; MWD: microwave diathermy; PMI: pectoralis minor index; ESWT: extracorporeal shock wave therapy; SPADI: Shoulder Pain and Disability Index; SST: special sports training therapy; SIS: shoulder impingement syndrome; VAS: visual analog scale.

**Fig 7.**
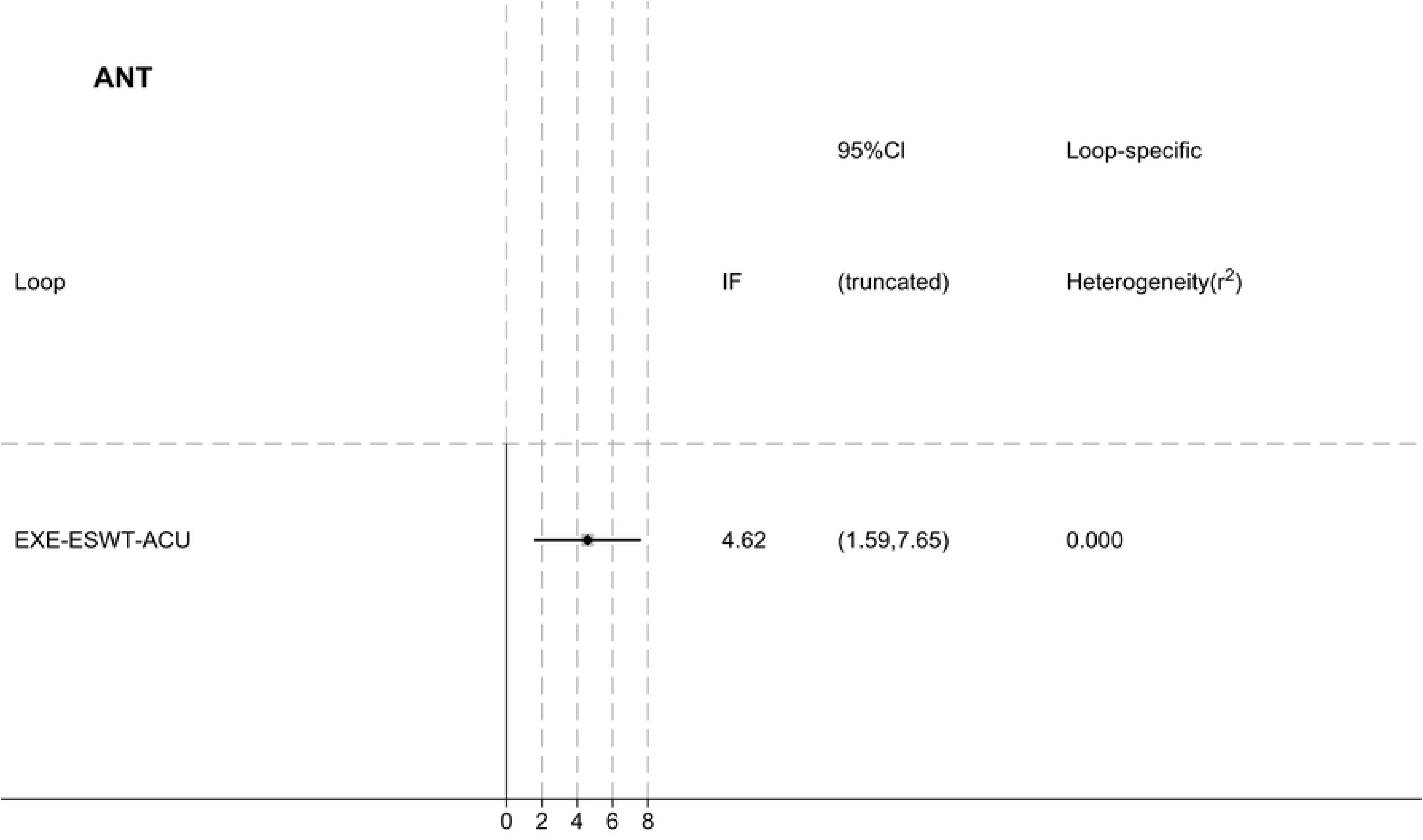
Shoulder flexion range of motion inconsistency test. ACU: acupuncture; CI: confidence interval; EXE: exercise therapy; ESWT: extracorporeal shock wave therapy.

**Fig 8.**
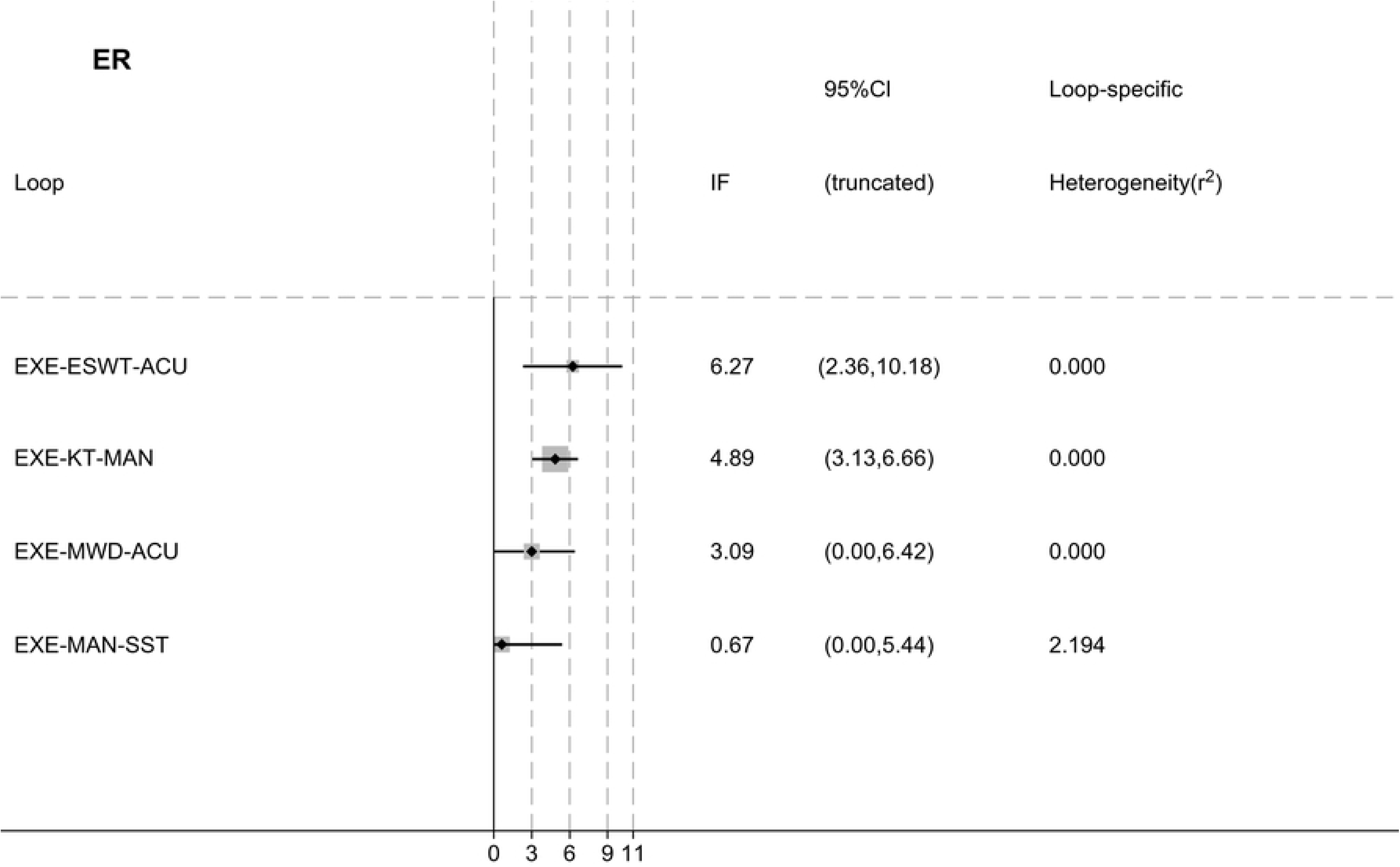
Shoulder external rotation range of motion inconsistency test.

**Fig 9.**
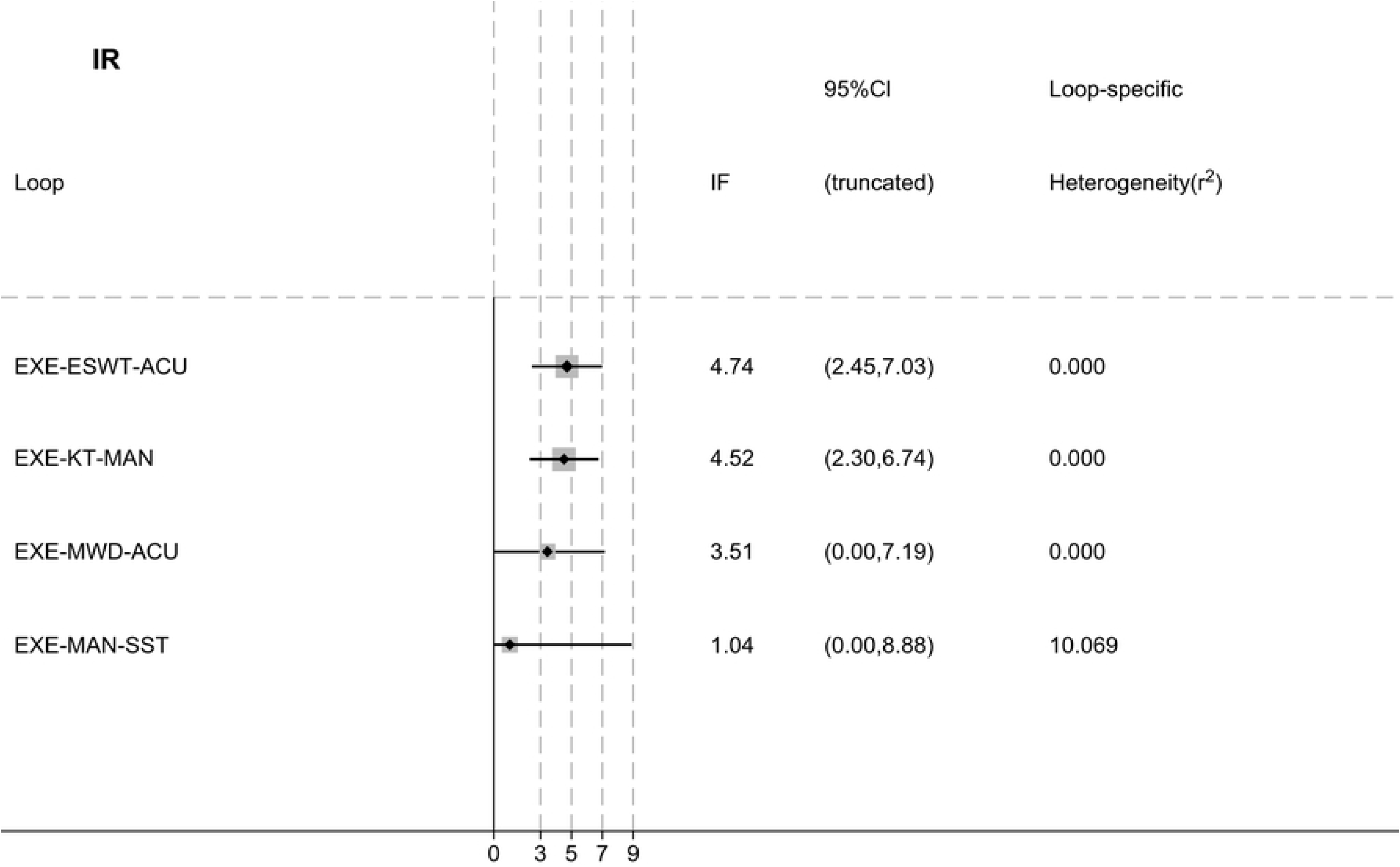
Shoulder internal rotation range of motion inconsistency test. ACU: acupuncture; EXE: exercise therapy; IR: internal rotation; KT: Kinesio taping; MAN: manual therapy; MWD: microwave diathermy; ESWT: extracorporeal shock wave therapy; SST: special sports training therapy.

Preliminary heterogeneity analyses were conducted after grouping the interventions into four broad categories: exercise therapy, physical therapy, traditional Chinese medicine therapy, and special sports training (Table 8). No closed loops were formed for shoulder pain and disability index (SPADI), Disabilities of the Arm, Shoulder, and Hand questionnaire (DASH), or acromiohumeral distance, and these outcomes therefore did not meet the assumptions required for consistency testing. Significant heterogeneity was observed for ER (P = 0.0007), IR (P = 0.0497), the IR/ER strength ratio (P = 0.0009), and adduction (P < 0.0001), whereas VAS, Constant–Murley Score, shoulder flexion, extension, and rear protraction showed no statistically significant heterogeneity (all P > 0.05). Additionally, the pectoralis minor index (PMI) showed borderline heterogeneity (P = 0.0598). After treatment regrouping, repeated pooled effect-size analysis of the remaining outcomes showed that ER remained heterogeneous (P = 0.0013), whereas VAS, IR, and shoulder flexion remained comparatively stable (Table 9).

**Table 8.** Preliminary pooled effect size of different treatment methods on various outcome measures.

| Outcome indicator | Therapeutic method | Prob>chi2 | Chi2 | p |
| --- | --- | --- | --- | --- |
| VAS | 1\2\3\4 | 0.8847 | 1.16 | 0.885 |
| CMS | 1\2\3\4 | 0.3635 | 2.02 | 0.363 |
| SPADI | 1\2\3\4 | **Does not meet consistency test** |  |  |
| DASH | 1\2\3\4 | **Does not meet consistency test** |  |  |
| PMI | 1\2\3\4 | 0.0598 | 3.54 | 0.06 |
| AHD | 1\2\3\4 | **Does not meet consistency test** |  |  |
| ER | 1\2\3\4 | 0.0007* | 21.37 | 0.000 |
| IR | 1\2\3\4 | 0.0497* | 11.09 | 0.05 |
| RP | 1\3\4 | 0.3863 | 1.9 | 0.386 |
| IR/ER | 1\3\4 | 0.0009* | 13.92 | 0.000 |
| ANT | 1\3\4 | 0.7907 | 0.47 | 0.791 |
| EXT | 1\2\3\4 | 0.3625 | 0.83 | 0.363 |
| ADD | 1\3\4 | 0.0000* | 46.35 | 0.000 |

1. Exercise therapy; 2. physical therapy; 3. traditional Chinese medicine treatment; 4. specialized sports training therapy
ANT: antexion; AHD: acromiohumeral distance; ADD: adduction; CMS: Constant–Murley score; DASH: Disabilities of the Arm, Shoulder, and Hand questionnaire; EST: electrical stimulation therapy; EXT: extended; EXE: exercise therapy; ER: external rotation; IR: internal rotation; PMI: pectoralis minor index; RP: Rear Protraction; SPADI: Shoulder Pain and Disability Index; VAS: visual analog scale.

**Table 9.** Pooled effect size of different treatment methods on the remaining outcome indicators.

| Outcome indicator | Therapeutic method | Prob>chi [2] | chi<sup>2</sup> | p |
| --- | --- | --- | --- | --- |
| VAS | EXE EST KT ESWT MFT<br>ACU MAN SST | 0.7979 | 1.66 | 0.798 |
| ER | EXE EST KT ESWT MWD<br>ACU MAN SST | 0.0013* | 19.92 | 0.001 |
| IR | EXE EST KT ESWT MWD<br>ACU MAN SST | 0.2999 | 6.06 | 0.3 |
| ANT | EXE ESWT ACU MAN<br>SST | 0.3648 | 0.84 | 0.365 |

ACU: acupuncture; EST: electrical stimulation therapy; EXT: extended; EXE: exercise therapy; ER: external rotation; IR: internal rotation; KT: Kinesio taping; MAN: manual therapy; MFT: medium-frequency therapy apparatus; MWD: microwave diathermy; ESWT: extracorporeal shock wave therapy; SPADI: Shoulder Pain and Disability Index; SST: special sports training therapy; VAS: visual analog scale.

### Main network meta-analysis results

#### Pain intensity assessed by VAS

Eight treatment approaches were analyzed for the VAS outcome, including exercise therapy, electrical stimulation therapy, Kinesio taping, extracorporeal shock wave therapy, medium-frequency therapy, acupuncture, manual therapy, and special sports training. As shown in Table 10, manual therapy versus exercise therapy showed a statistically significant reduction in VAS score (MD = −3.044, 95% CI −5.440 to −0.647, P = 0.013). In contrast, electrical stimulation therapy, Kinesio taping, extracorporeal shock wave therapy, medium-frequency therapy, acupuncture, and special sports training did not differ significantly from exercise therapy (all P > 0.05). Similarly, the indirect comparisons of ESWT versus EXE/ESWT/ACU and ACU versus EXE/ACU or ESWT/ACU were non-significant. As shown in Fig 10, most pooled estimates were shifted toward benefit relative to exercise therapy, although only the comparison between manual therapy versus exercise therapy showed a clearly significant direct effect.

**Fig 10.**
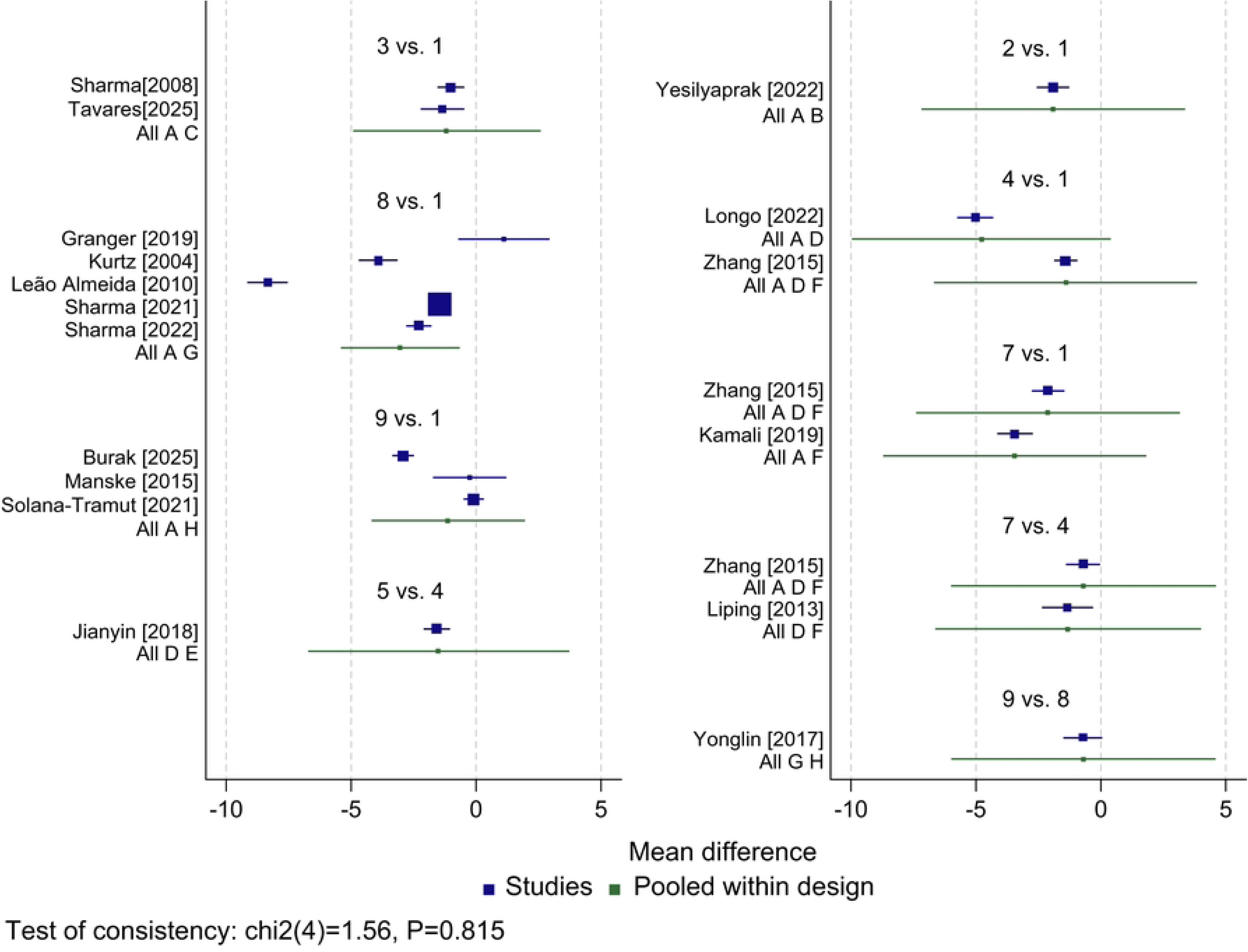
Forest plot of different treatment methods on patients’ visual analog scale scores.

**Table 10.** Pooled effect size of different treatment methods on patients’ visual analog scale scores.

| Type | Control group | Mean | SD | z | p>\|z\| | 95%CI |  |
| --- | --- | --- | --- | --- | --- | --- | --- |
| EST | EXE | -1.900 | 2.708 | -0.70 | 0.483 | -7.209 | 3.409 |
| KT | EXE | -1.189 | 1.914 | -0.62 | 0.534 | -4.941 | 2.562 |
| ESWT | EXE\ESWT\ACU | 3.593 | 3.825 | 0.94 | 0.358 | -3.903 | 11.089 |
|  | EXE | -5.003 | 2.711 | -1.85 | 0.065 | -10.315 | 0.310 |
| MFT | EXE | -6.601 | 3.825 | -1.73 | 0.084 | -14.098 | 0.896 |
| ACU | EXE\ACU | -1.330 | 3.833 | -0.35 | 0.729 | -8.843 | 6.183 |
|  | ESWT\ACU | -4.221 | 4.710 | -0.90 | 0.370 | -13.453 | 5.011 |
|  | EXE | -2.110 | 2.709 | -0.78 | 0.436 | -7.420 | 3.200 |
| MAN | EXE | -3.044 | 1.223 | -2.49 | 0.013* | -5.440 | -0.647 |
| SST | MAN\SST | -2.634 | 3.370 | -0.78 | 0.434 | -9.239 | 3.971 |
|  | EXE | -1.118 | 1.574 | -0.71 | 0.478 | -4.204 | 1.967 |

ACU: acupuncture; EXE: exercise therapy; IR: internal rotation; KT: Kinesio taping; MAN: manual therapy; MFT: medium-frequency therapy; MWD: microwave diathermy; ESWT: extracorporeal shock wave therapy; SST: special sports training therapy.

Table 11 further demonstrates the pairwise comparisons of treatment effects on pain intensity assessed by VAS in 637 swimmers with shoulder impingement syndrome. Compared with exercise therapy alone, medium-frequency therapy plus exercise therapy showed the most favorable point estimate (MD −4.58, 95% CI −10.12 to 0.95), followed by acupuncture plus exercise therapy (MD −3.48, 95% CI −6.45 to −0.50). Fig 11 complements these findings by demonstrating the pairwise relative treatment effects for pain-related outcomes, together with both 95% CIs and 95% prediction intervals. In Fig 11a, medium-frequency therapy plus exercise therapy and acupuncture plus exercise therapy remained the most favorable pairwise comparisons for VAS; however, the corresponding prediction intervals crossed the null (−13.30 to 4.13 and −10.13 to 3.18, respectively), indicating uncertainty in the expected effect across future studies. Taken together, these pairwise findings support a potential benefit of acupuncture- and medium-frequency-based adjunctive treatment for pain-related outcomes; however, they should be interpreted cautiously in light of the wide prediction intervals.

**Fig 11.**
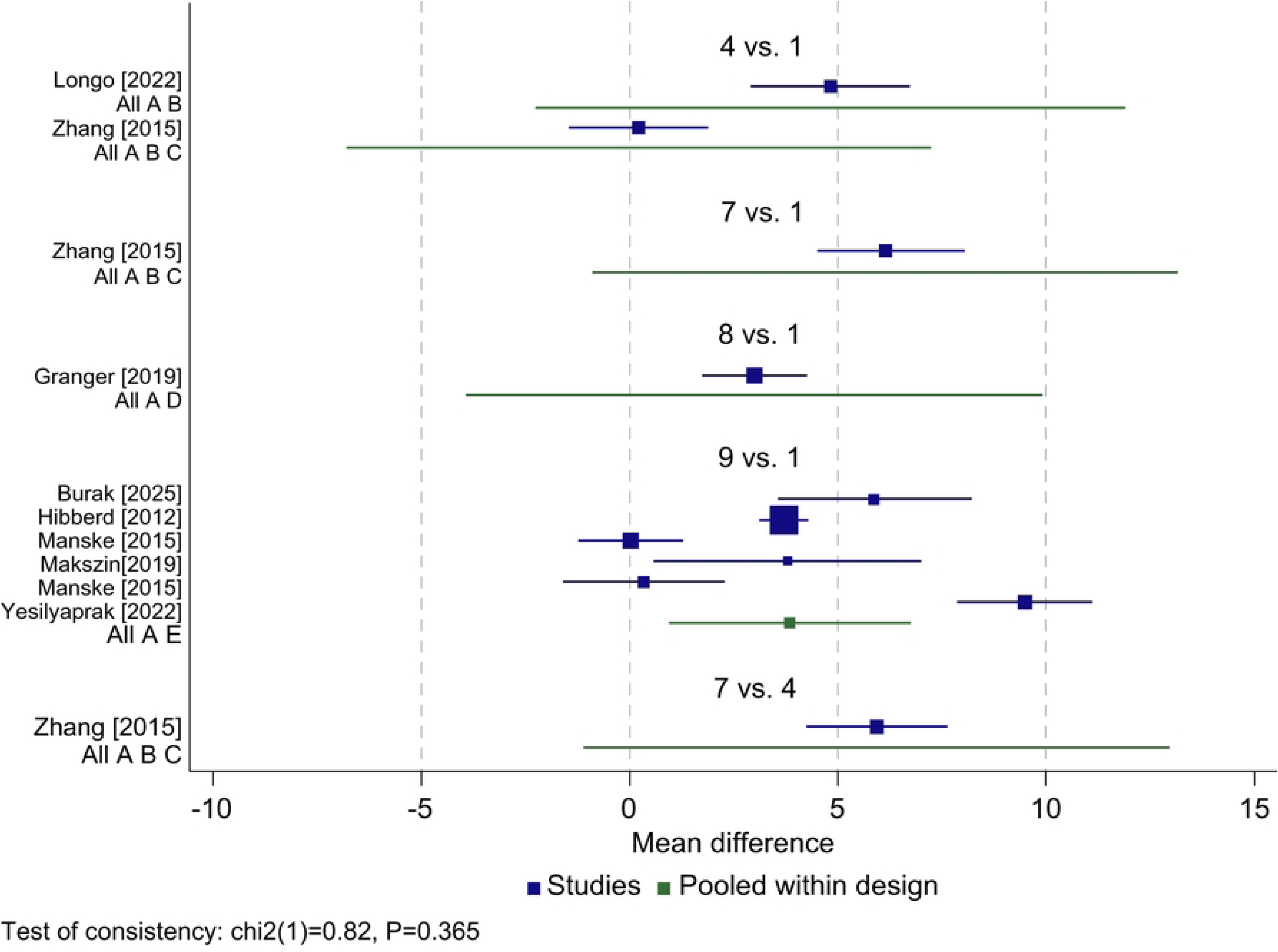
Pairwise comparisons of treatment effects on pain-related outcomes in swimmers with shoulder impingement syndrome. (a) Pairwise comparison of different treatment methods on patients’ visual analog scale evaluation. (b) A pair-to-pair comparison of the effects of different treatment methods on the index of shoulder joint pain and dysfunction. ACU: acupuncture; EST: electrical stimulation therapy; EXE: exercise therapy; KT: Kinesio taping; MFT: medium-frequency therapy; MAN: manual therapy; MWD: microwave diathermy; ESWT: extracorporeal shock wave therapy; SST: special sports training therapy.

**Table 11.**
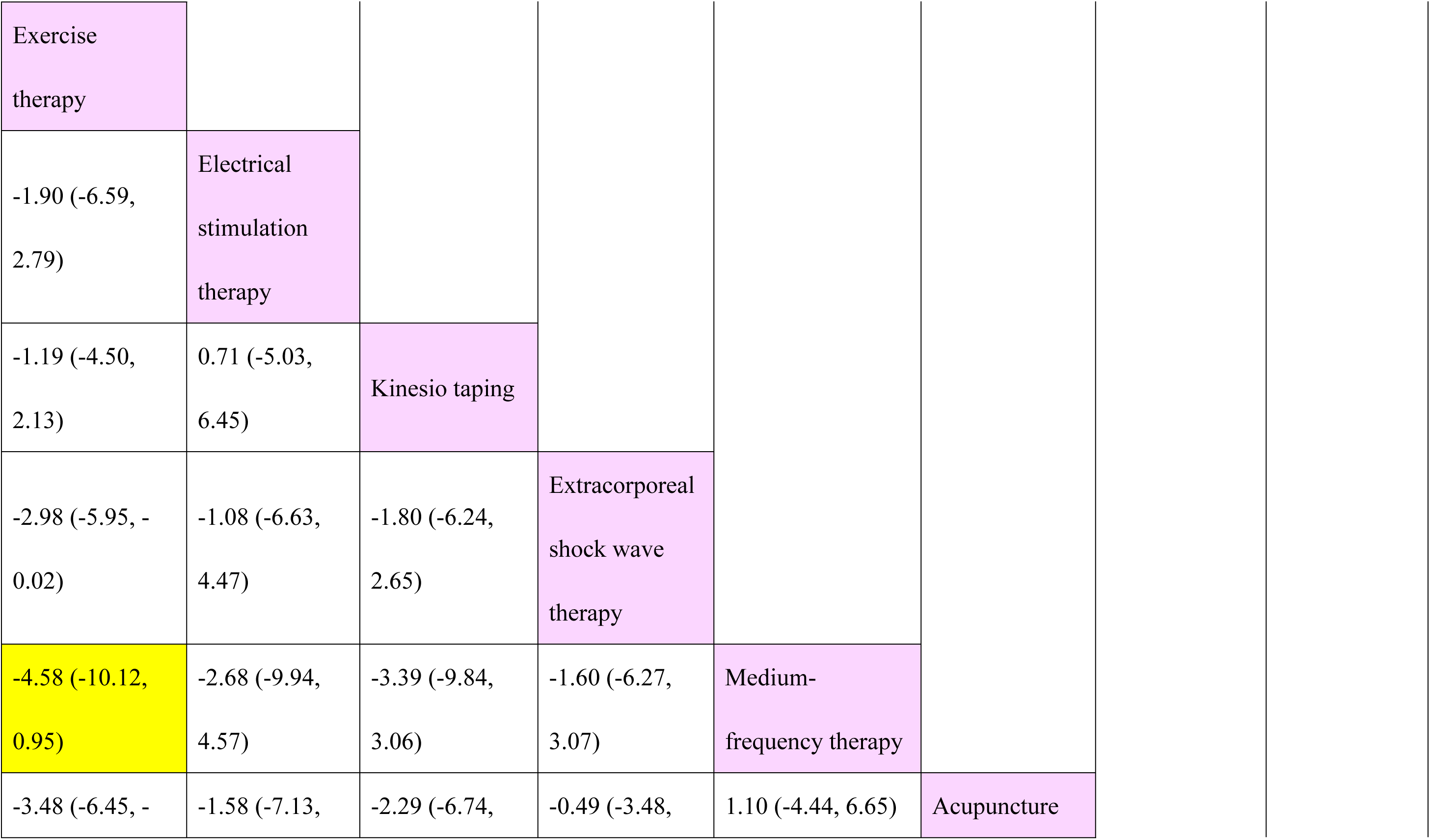
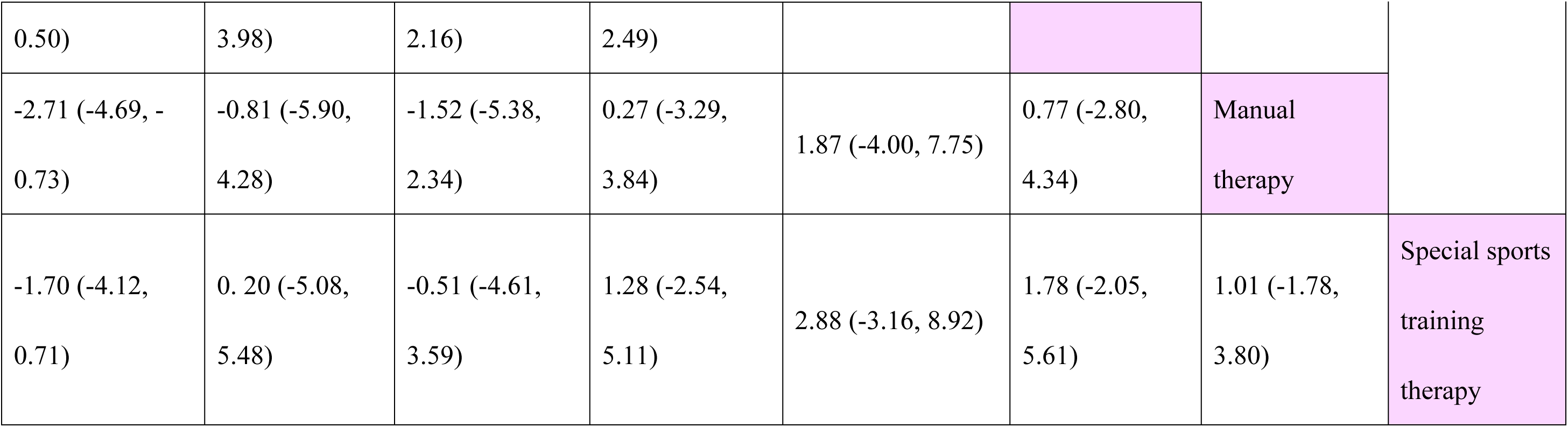
Pairwise comparison of the effects of different treatment methods on the evaluation of patients’ visual analog scale.

|  |  |  |  |  |  |
| --- | --- | --- | --- | --- | --- |
| Exercise therapy |  |  |  |  |  |
| -1.90 (-6.59, 2.79) | Electrical stimulation therapy |  |  |  |  |
| -1.19 (-4.50, 2.13) | 0.71 (-5.03, 6.45) | Kinesio taping |  |  |  |
| -2.98 (-5.95, -0.02) | -1.08 (-6.63, 4.47) | -1.80 (-6.24, 2.65) | Extracorporeal shock wave therapy |  |  |
| -4.58 (-10.12, 0.95) | -2.68 (-9.94, 4.57) | -3.39 (-9.84, 3.06) | -1.60 (-6.27, 3.07) | Medium-frequency therapy |  |
| -3.48 (-6.45, - | -1.58 (-7.13, | -2.29 (-6.74, | -0.49 (-3.48, | 1.10 (-4.44, 6.65) | Acupuncture |

|  |  |  |  |  |  |  |  |
| --- | --- | --- | --- | --- | --- | --- | --- |
| 0.50) | 3.98) | 2.16) | 2.49) |  |  |  |  |
| -2.71 (-4.69, -<br>0.73) | -0.81 (-5.90,<br>4.28) | -1.52 (-5.38,<br>2.34) | 0.27 (-3.29,<br>3.84) | 1.87 (-4.00, 7.75) | 0.77 (-2.80,<br>4.34) | Manual<br>therapy |  |
| -1.70 (-4.12,<br>0.71) | 0.20 (-5.08,<br>5.48) | -0.51 (-4.61,<br>3.59) | 1.28 (-2.54,<br>5.11) | 2.88 (-3.16, 8.92) | 1.78 (-2.05,<br>5.61) | 1.01 (-1.78,<br>3.80) | Special sports<br>training<br>therapy |

### Shoulder flexion range of motion

Five treatment approaches were included in the shoulder flexion network: exercise therapy, extracorporeal shock wave therapy, acupuncture, manual therapy, and special sports training. As shown in Table 12, special sports training significantly improved shoulder flexion compared to exercise therapy (MD = 3.855, 95% CI 0.956 to 6.755, P = 0.009). By contrast, acupuncture (MD = 6.150, 95% CI −0.875 to 13.175, P = 0.086) and manual therapy (MD = 3.000, 95% CI −3.946 to 9.946, P = 0.397) did not differ significantly from exercise therapy, and the indirect ESWT comparison was also non-significant. Fig 11b demonstrates that, for the SPADI, acupuncture plus exercise therapy yielded a favorable estimate compared with electrical stimulation therapy plus exercise therapy, although the corresponding prediction interval remained wide (−51.08 to 77.60), suggesting substantial uncertainty across studies. Fig 12 visually illustrates that most pooled estimates favored active treatment over exercise therapy; however, the precision of these estimates varied substantially across comparisons. In the pairwise comparison table, acupuncture plus exercise therapy showed a favorable estimate compared with exercise therapy (MD = 7.29, 95% CI 0.81 to 13.77), although the corresponding prediction interval remained wide.

**Fig 12.**
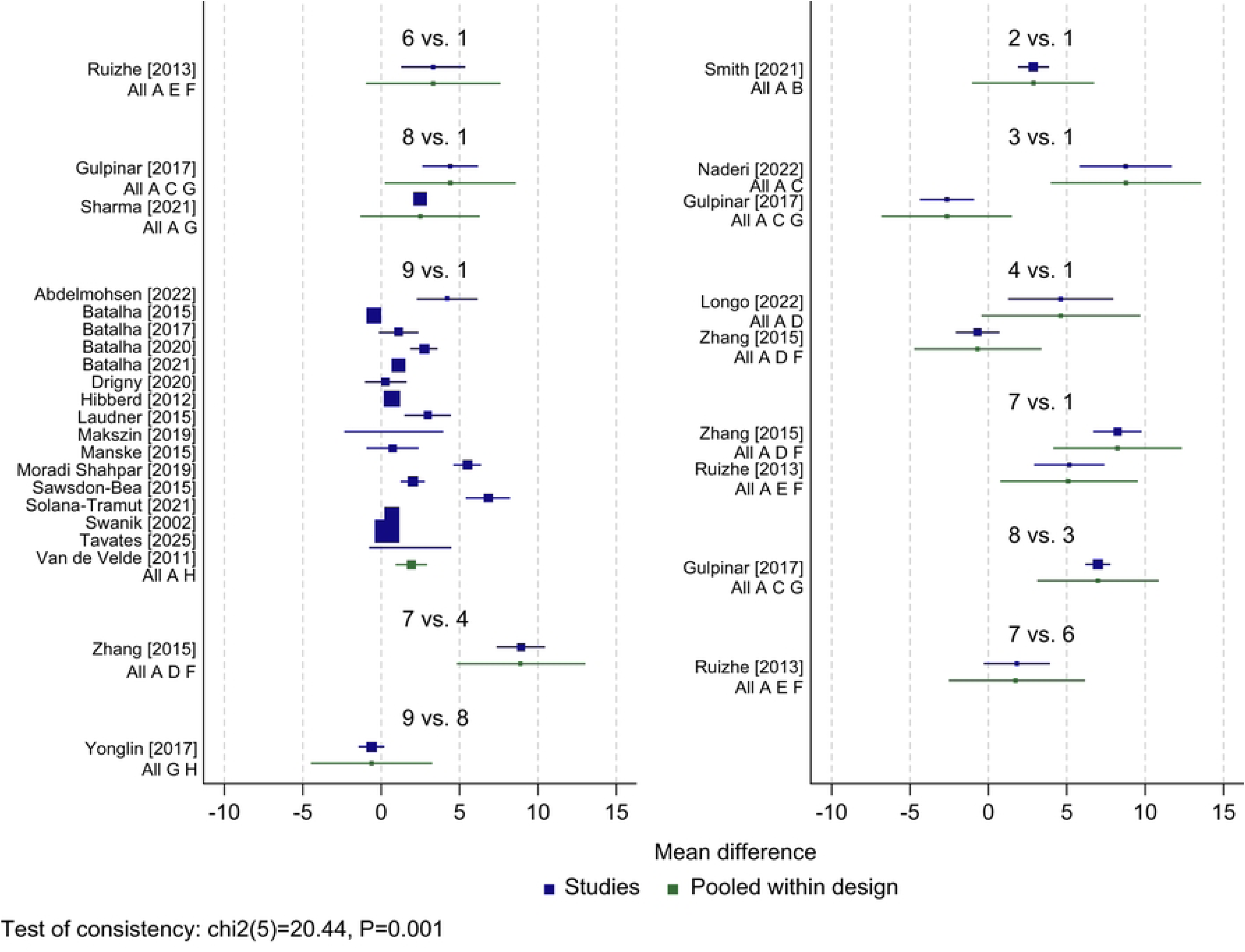
Forest plot of different treatment methods on patients’ antexion shoulder flexion range of motion.

**Table 12.** Combined effect size of different treatment methods on patients’ antexion shoulder flexion range of motion.

| Type | Control group | Mean | SD | z | P>\|z\| | 95%CI |  |
| --- | --- | --- | --- | --- | --- | --- | --- |
| ESWT | EXE\ESWT\ACU | -4.620 | 5.098 | 0.365 | 0.365 | -14.611 | 5.371 |
|  | EXE | 4.830 | 3.620 | 0.182 | 0.182 | -2.265 | 11.925 |
| ACU | EXE | 6.150 | 3.584 | 1.72 | 0.086 | -0.875 | 13.175 |
| MAN | EXE | 3.000 | 3.544 | 0.85 | 0.397 | -3.946 | 9.946 |
| SST | EXE | 3.855 | 1.479 | 2.61 | 0.009* | 0.956 | 6.755 |

ACU: acupuncture; EXE: exercise therapy; MAN: manual therapy; MWD: microwave diathermy; ESWT: extracorporeal shock wave therapy; SD: standard deviation; CI: confidence interval

### External rotation

Seven treatment approaches were included in the ER analysis: exercise therapy, Kinesio taping, extracorporeal shock wave therapy, microwave diathermy, acupuncture, manual therapy, and special sports training. As shown in Table 13, significant effects were observed for Kinesio taping versus exercise therapy/Kinesio taping/manual therapy (MD = −11.401, 95% CI −17.836 to −4.966, P = 0.001), Kinesio taping versus exercise therapy (MD = 8.800, 95% CI 3.952 to 13.648, P = 0.001), acupuncture versus exercise therapy (MD = 8.260, 95% CI 4.102 to 12.418, P = 0.001), manual therapy versus exercise therapy (MD = 4.418, 95% CI 0.171 to 8.665, P = 0.041), and special sports training versus exercise therapy (MD = 2.057, 95% CI 0.998 to 3.115, P = 0.001). By contrast, electrical stimulation therapy, extracorporeal shock wave therapy, and microwave diathermy did not differ significantly from exercise therapy. As shown in Fig 13, the pooled estimates for Kinesio taping, acupuncture, manual therapy, and special sports training were consistently shifted toward benefit relative to exercise therapy, with acupuncture and Kinesio taping showing the most favorable overall pattern. Table 14 further demonstrates the pairwise comparison profile for shoulder external rotation. Among the comparisons versus exercise therapy, acupuncture plus exercise therapy showed the largest favorable estimate (MD 7.49, 95% CI 3.65 to 11.34), followed by manual therapy (MD 4.22, 95% CI 1.16 to 7.29), special sports training (MD 2.17, 95% CI 0.79 to 3.54), and electrical stimulation therapy (MD 2.90, 95% CI −2.46 to 8.26). The comparisons also showed that acupuncture outperformed Kinesio taping (MD 5.88, 95% CI 0.44 to 11.32) and that special sports training was inferior to acupuncture (MD −5.33, 95% CI −9.41 to −1.25). These pairwise findings are consistent with the broader network pattern, in which Kinesio taping and acupuncture showed the most favorable effects on external rotation. Fig 14 visually supports these pairwise findings and further shows that the prediction interval for acupuncture plus exercise therapy remained above the null value, suggesting a relatively more robust favorable effect than that observed for most other interventions.

**Fig 13.**
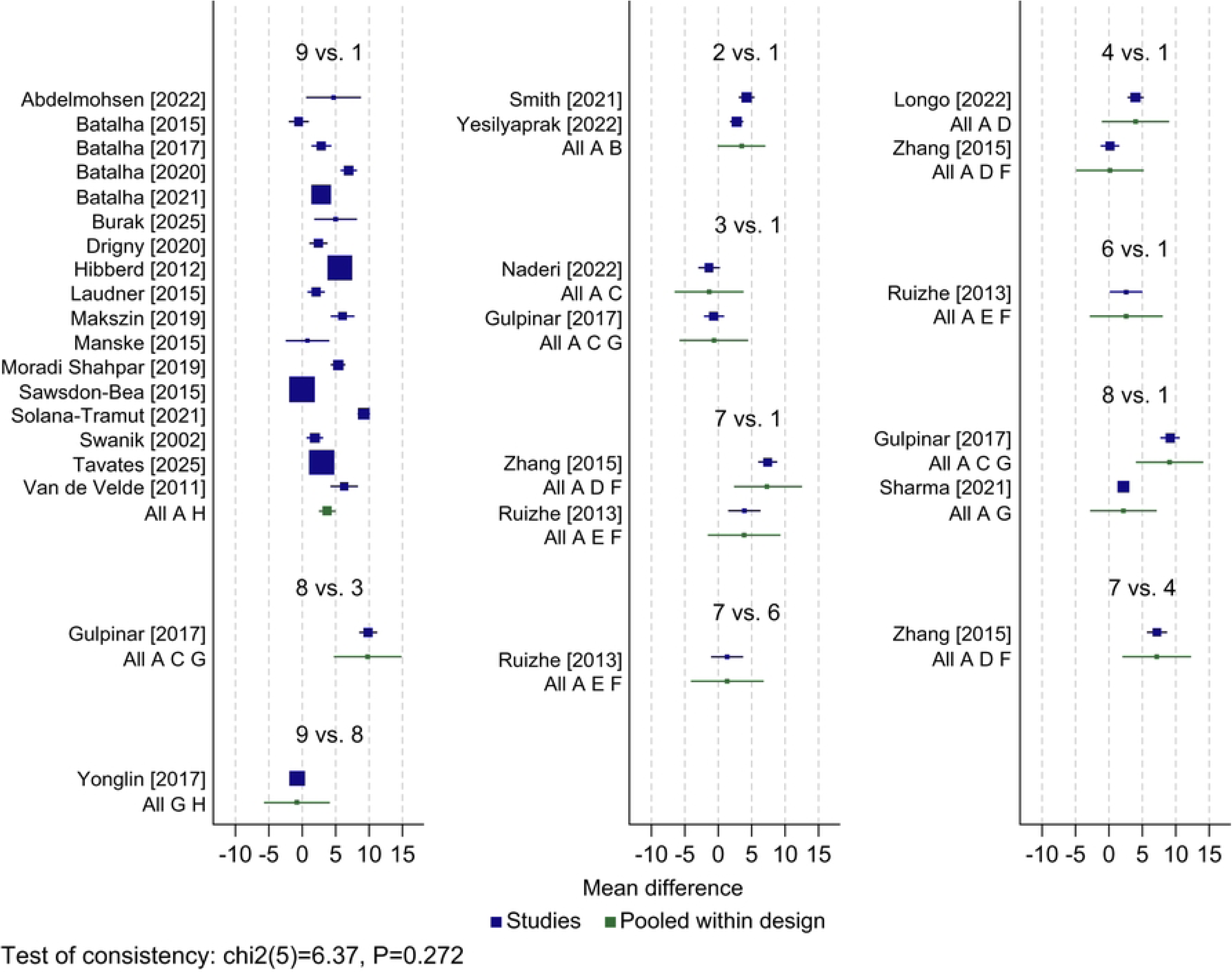
Forest plot of different treatment methods on patients’ external rotation range of shoulder joint.

**Fig 14.**
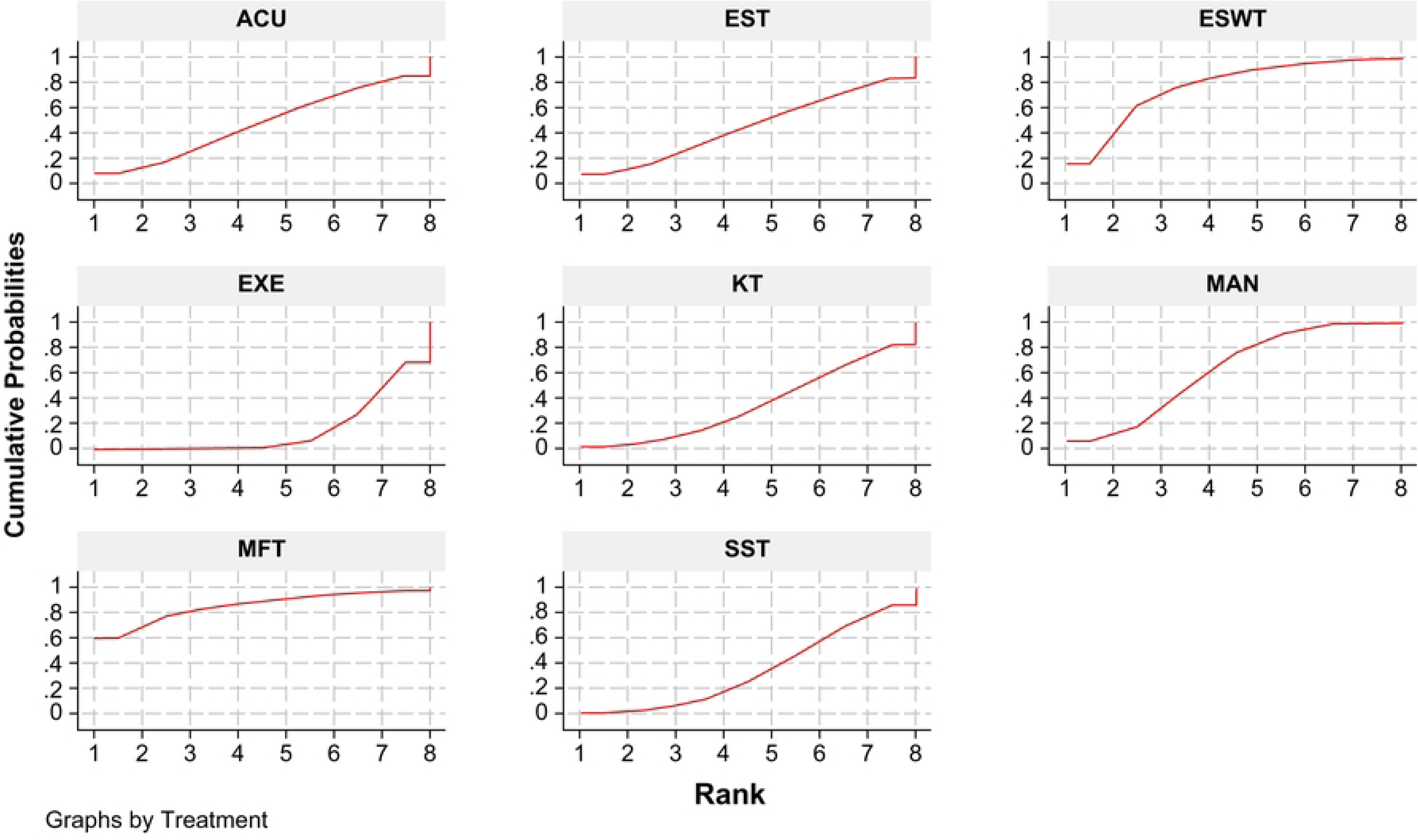
Pairwise comparison of the influence of different treatment methods on external rotation of shoulder joint. ACU: acupuncture; EXE: exercise therapy; MAN: manual therapy; MWD: microwave diathermy; KT: Kinesio taping; ESWT: extracorporeal shock wave therapy; SST: special sports training therapy; CI: confidence interval.

**Table 13.** Combined effect size of different treatment methods on patients’ external rotation motion of shoulder joint.

| Type | Control group | Mean | SD | z | P>\|z\| | 95%CI |  |
| --- | --- | --- | --- | --- | --- | --- | --- |
| EST | EXE | 2.900 | 2.032 | 1.43 | 0.154 | -1.083 | 6.883 |
| KT | EXE\KT\MAN | -11.401 | 3.283 | -3.47 | 0.001* | -17.836 | -4.966 |
|  | EXE | 8.800 | 2.473 | 3.56 | 0.001* | 3.952 | 13.648 |
| ESWT | EXE\ESWT\MWD | -5.270 | 3.348 | -1.57 | 0.115 | -11.832 | 1.292 |
|  | EXE | 4.620 | 2.610 | 1.77 | 0.077 | -0.496 | 9.736 |
| MWD | EXE | 3.330 | 2.226 | 1.50 | 0.135 | -1.033 | 7.693 |
| ACU | EXE\MWD\ACU | -3.090 | 3.113 | -0.99 | 0.321 | -9.191 | 3.011 |
|  | EXE | 8.260 | 2.122 | 3.89 | 0.001* | 4.102 | 12.418 |
| MAN | EXE\MAN | -1.918 | 2.936 | -0.65 | 0.514 | -7.672 | 3.836 |
|  | EXE | 4.418 | 2.167 | 2.04 | 0.041* | 0.171 | 8.665 |
| SST | MAN\SST | 1.750 | 3.003 | 0.58 | 0.560 | -4.136 | 7.637 |
|  | EXE | 2.057 | 0.540 | 3.81 | 0.001* | 0.998 | 3.115 |

ACU: acupuncture; EXE: exercise therapy; KT: Kinesio taping; MAN: manual therapy; MWD: microwave diathermy; ESWT: extracorporeal shock wave therapy; SST: special sports training therapy; SD: standard deviation; CI: confidence interval.

**Table 14.**
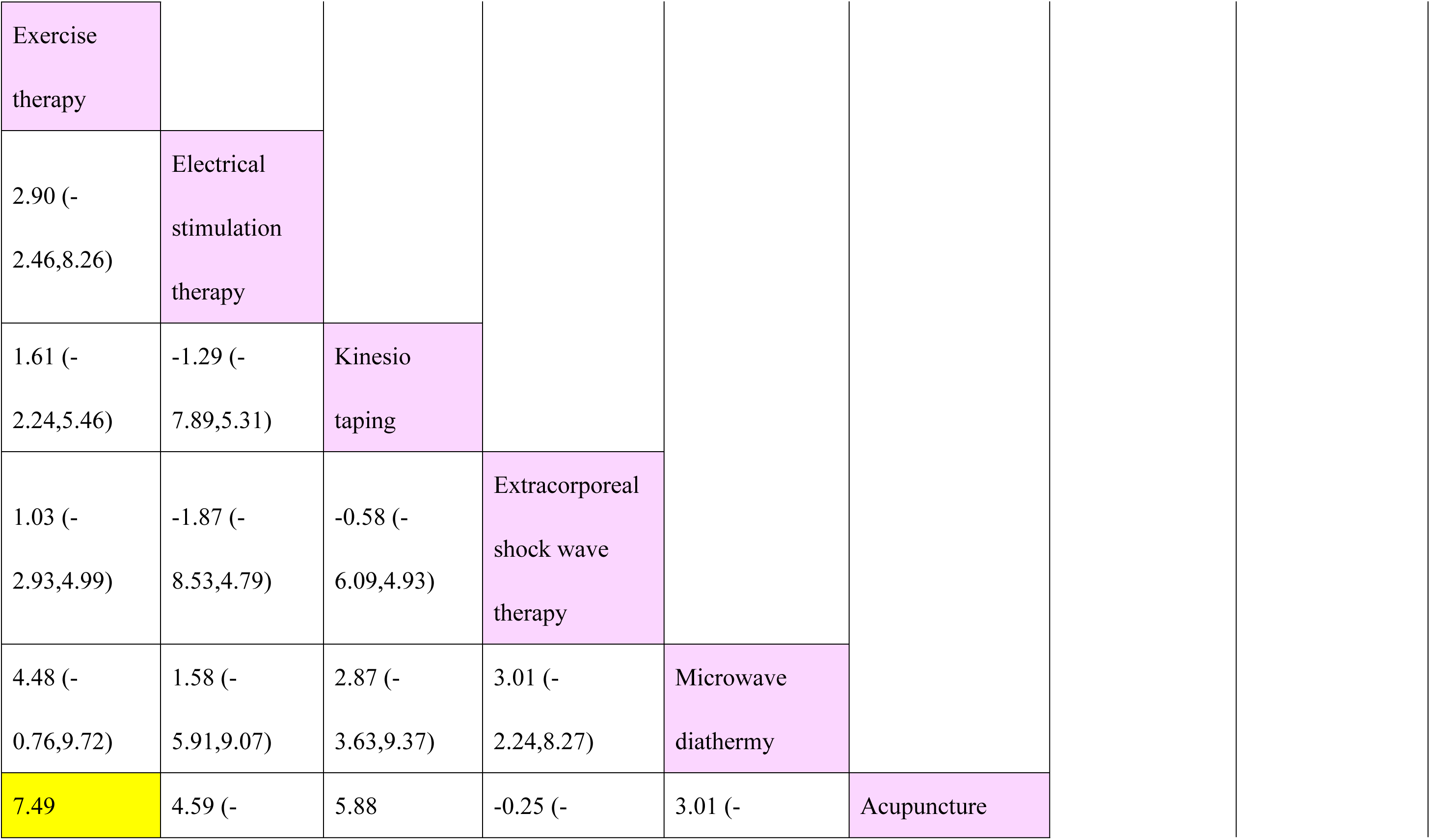
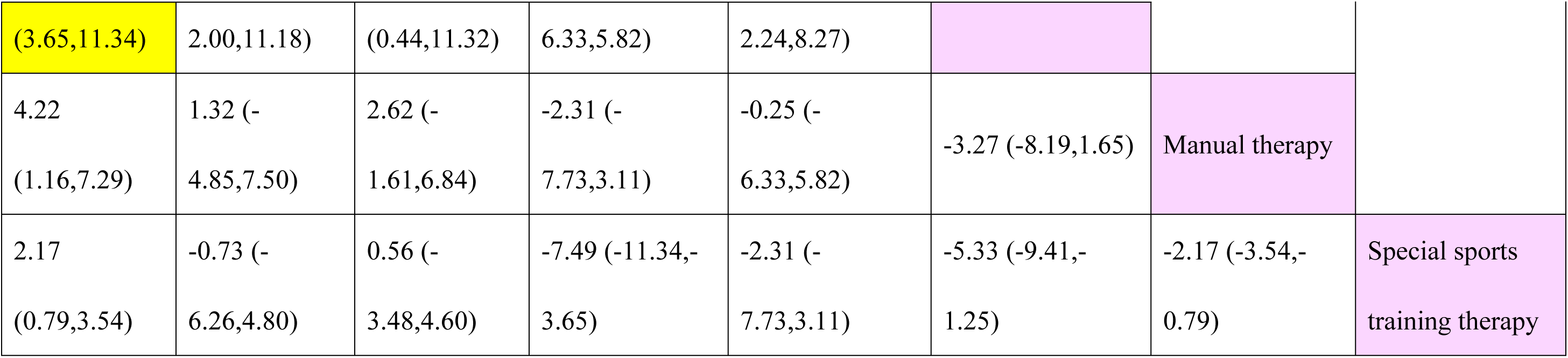
Influence of different treatment methods on external rotation motion of shoulder joint in patients.

|  |  |  |  |  |  |
| --- | --- | --- | --- | --- | --- |
| Exercise therapy |  |  |  |  |  |
| 2.90 (-2.46,8.26) | Electrical stimulation therapy |  |  |  |  |
| 1.61 (-2.24,5.46) | -1.29 (-7.89,5.31) | Kinesio taping |  |  |  |
| 1.03 (-2.93,4.99) | -1.87 (-8.53,4.79) | -0.58 (-6.09,4.93) | Extracorporeal shock wave therapy |  |  |
| 4.48 (-0.76,9.72) | 1.58 (-5.91,9.07) | 2.87 (-3.63,9.37) | 3.01 (-2.24,8.27) | Microwave diathermy |  |
| 7.49 | 4.59 (- | 5.88 | -0.25 (- | 3.01 (- | Acupuncture |

|  |  |  |  |  |  |  |  |
| --- | --- | --- | --- | --- | --- | --- | --- |
| (3.65,11.34) | 2.00,11.18) | (0.44,11.32) | 6.33,5.82) | 2.24,8.27) |  |  |  |
| 4.22<br>(1.16,7.29) | 1.32 (-<br>4.85,7.50) | 2.62 (-<br>1.61,6.84) | -2.31 (-<br>7.73,3.11) | -0.25 (-<br>6.33,5.82) | -3.27 (-8.19,1.65) | Manual therapy |  |
| 2.17<br>(0.79,3.54) | -0.73 (-<br>6.26,4.80) | 0.56 (-<br>3.48,4.60) | -7.49 (-11.34,-<br>3.65) | -2.31 (-<br>7.73,3.11) | -5.33 (-9.41,-<br>1.25) | -2.17 (-3.54,-<br>0.79) | Special sports<br>training therapy |

### Internal rotation

Seven treatment approaches were included in the IR analysis: exercise therapy, electrical stimulation therapy, Kinesio taping, extracorporeal shock wave therapy, microwave diathermy, acupuncture, manual therapy, and special sports training. As shown in Table 15, statistically significant improvements were observed for acupuncture versus exercise therapy (MD = 7.380, 95% CI 2.117 to 12.643, P = 0.006), manual therapy versus exercise therapy (MD = 9.173, 95% CI 3.909 to 14.438, P = 0.001), and special sports training versus exercise therapy (MD = 3.875, 95% CI 2.534 to 5.216, P = 0.001). By contrast, electrical stimulation therapy, Kinesio taping, extracorporeal shock wave therapy, and microwave diathermy did not differ significantly from exercise therapy, and the indirect comparisons involving EXE/KT/MAN, EXE/ESWT/MWD, and EXE/MWD/ACU were also non-significant. Fig 15 demonstrates that acupuncture, manual therapy, and special sports training were consistently positioned on the beneficial side of the pooled estimates, whereas electrical stimulation therapy, Kinesio taping, extracorporeal shock wave therapy, and microwave diathermy showed wider CIs crossing the null. Table 16 demonstrates the pairwise comparison profile for shoulder internal rotation. Compared with exercise therapy, acupuncture (MD 6.38, 95% CI 2.59 to 10.17), manual therapy (MD 5.39, 95% CI 2.37 to 8.41), and special sports training (MD 3.93, 95% CI 2.58 to 5.27) all demonstrated favorable estimates. In addition, acupuncture outperformed Kinesio taping (MD 8.48, 95% CI 3.21 to 13.74), and manual therapy also showed greater benefit than Kinesio taping (MD 7.48, 95% CI 3.34 to 11.62). These pairwise findings support the overall ranking profile, in which manual therapy and acupuncture showed the most favorable effects on internal rotation. Fig 16 visually confirms these pairwise findings, with manual therapy, acupuncture, and special sports training showing favorable estimates relative to exercise therapy.

**Fig 15.**
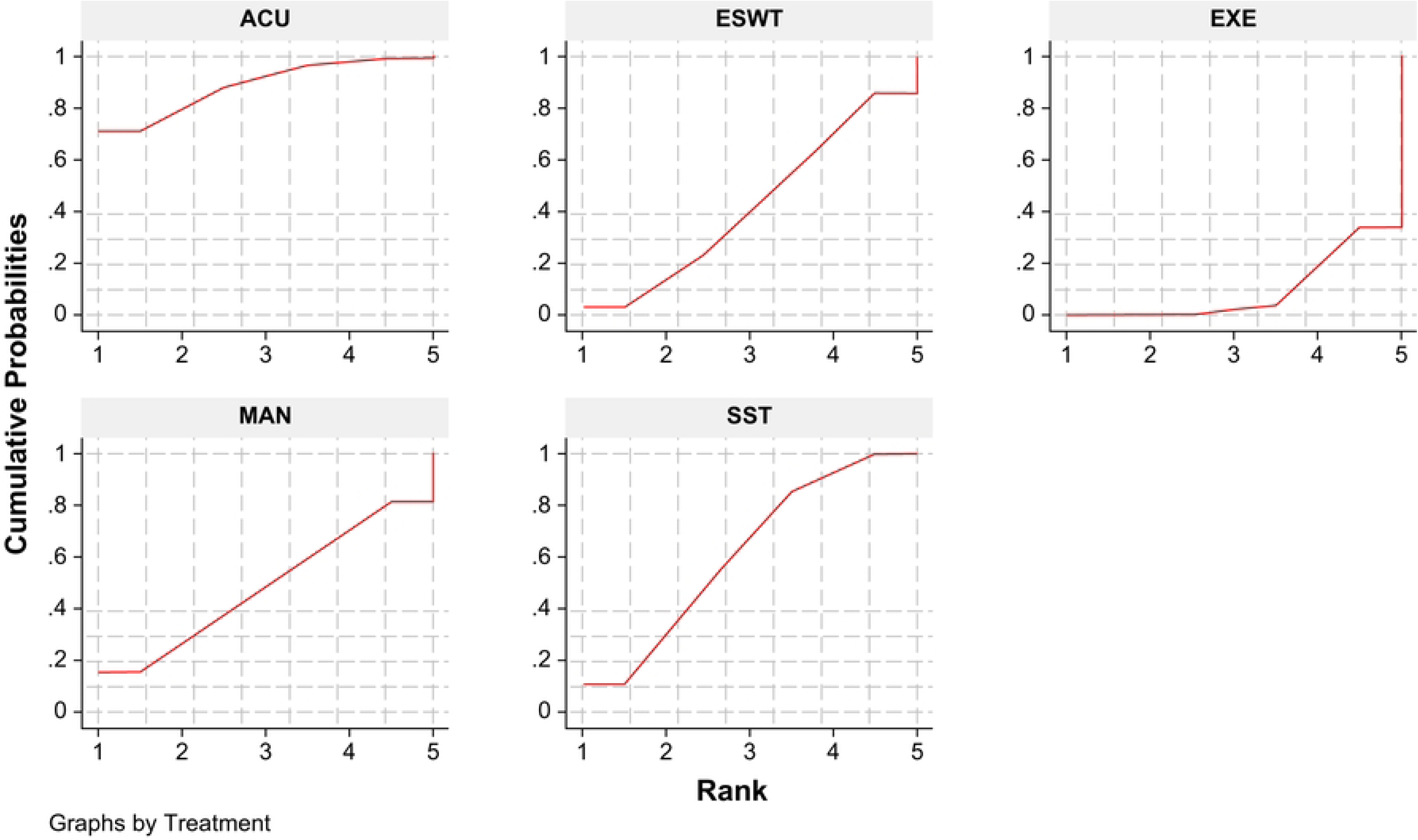
Forest plot of different treatment methods on patients’ internal rotation range of shoulder joint.

**Fig 16.**
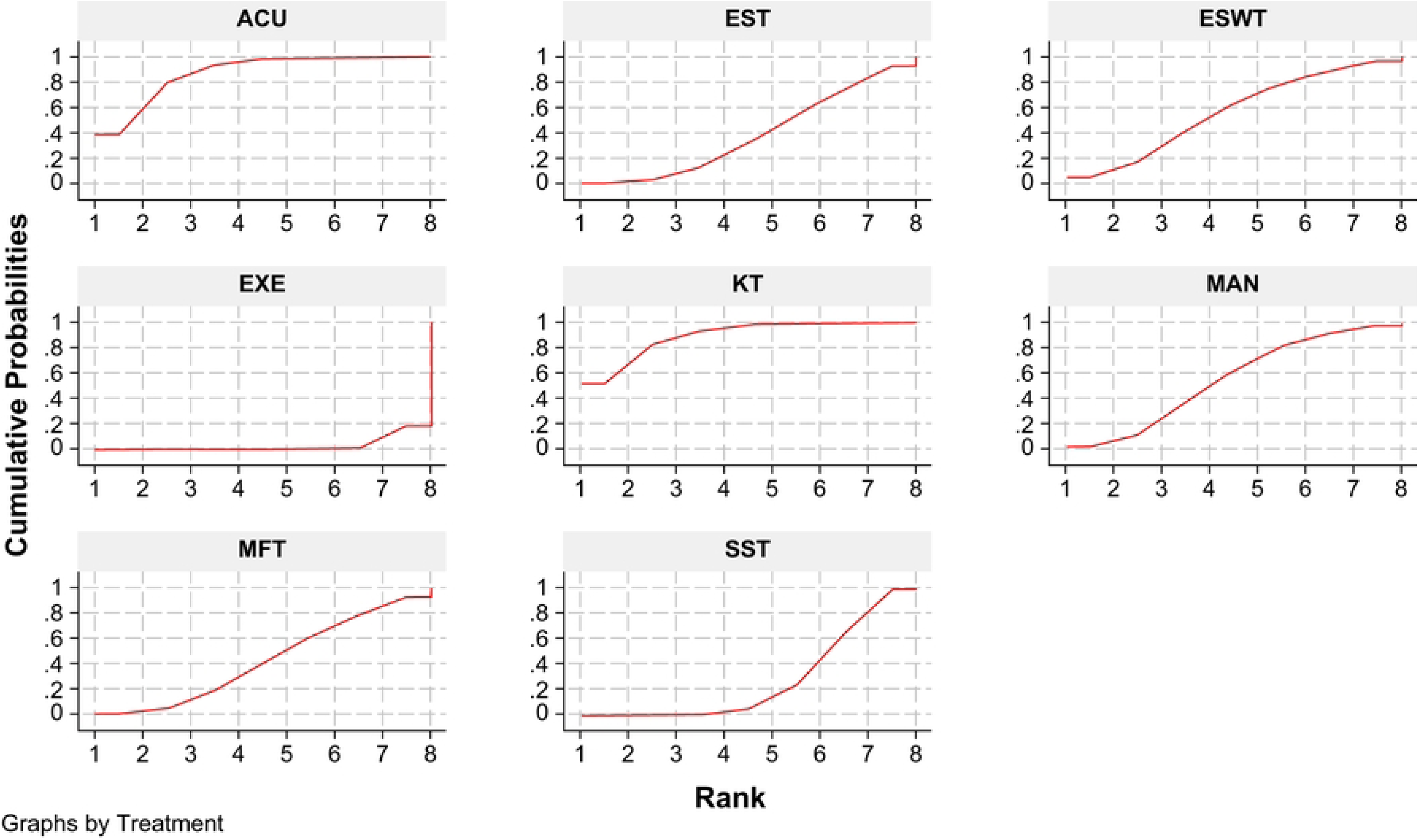
Pairwise comparison of the effects of different treatment methods on internal rotation of shoulder joint. ACU: acupuncture; EXE: exercise therapy; MAN: manual therapy; ESWT: extracorporeal shock wave therapy; KT: Kinesio taping; MWD: microwave diathermy; SST: special sports training therapy; CI: confidence interval.

**Table 15.** Combined effect size results of internal rotation score indices in patients with different treatments.

| Type | Control group | Mean | SD | z | P>\|z\| | 95%CI |  |
| --- | --- | --- | --- | --- | --- | --- | --- |
| EST | EXE | 3.445 | 1.866 | 1.85 | 0.065 | -0.211 | 7.102 |
| KT | EXE\KT\MAN | 0.717 | 3.823 | 0.19 | 0.851 | -6.776 | 8.209 |
|  | EXE | -1.400 | 2.709 | -0.52 | 0.605 | -6.709 | 3.909 |
| ESWT | EXE\ESWT\MWD | -3.810 | 3.773 | -1.01 | 0.313 | -11.204 | 3.584 |
|  | EXE | 4.010 | 2.657 | 1.51 | 0.131 | -1.197 | 9.217 |
| MWD | EXE | 2.580 | 2.859 | 0.90 | 0.367 | -3.024 | 8.184 |
| ACU | EXE\MWD\ACU | -3.510 | 3.922 | -0.89 | 0.371 | -11.197 | 4.177 |
|  | EXE | 7.380 | 2.685 | 2.75 | 0.006* | 2.117 | 12.643 |
| MAN | EXE\MAN | -6.973 | 3.751 | -1.86 | 0.063 | -14.325 | 0.379 |
|  | EXE | 9.173 | 2.686 | 3.42 | 0.001* | 3.909 | 14.438 |
| SST | MAN\SST | 4.576 | 3.798 | 1.20 | 0.228 | -2.868 | 12.019 |
|  | EXE | 3.875 | 0.684 | 5.66 | 0.001* | 2.534 | 5.216 |

ACU: acupuncture; EST: electrical stimulation therapy; EXE: exercise therapy; KT: Kinesio taping; MAN: manual therapy; MWD: microwave diathermy; ESWT: extracorporeal shock wave therapy; SST: special sports training therapy; SD: standard deviation; CI: confidence interval.

**Table 16.**
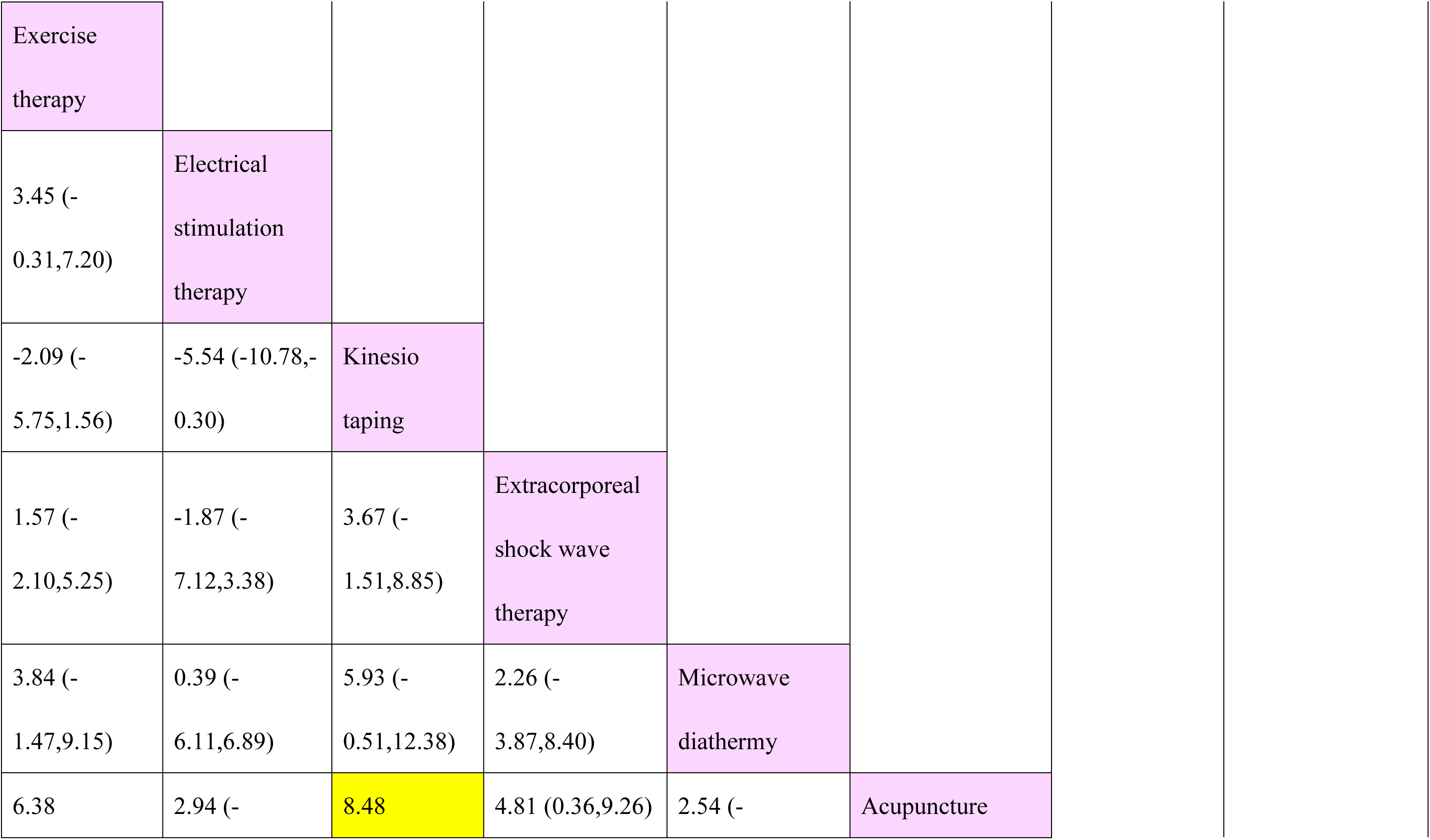
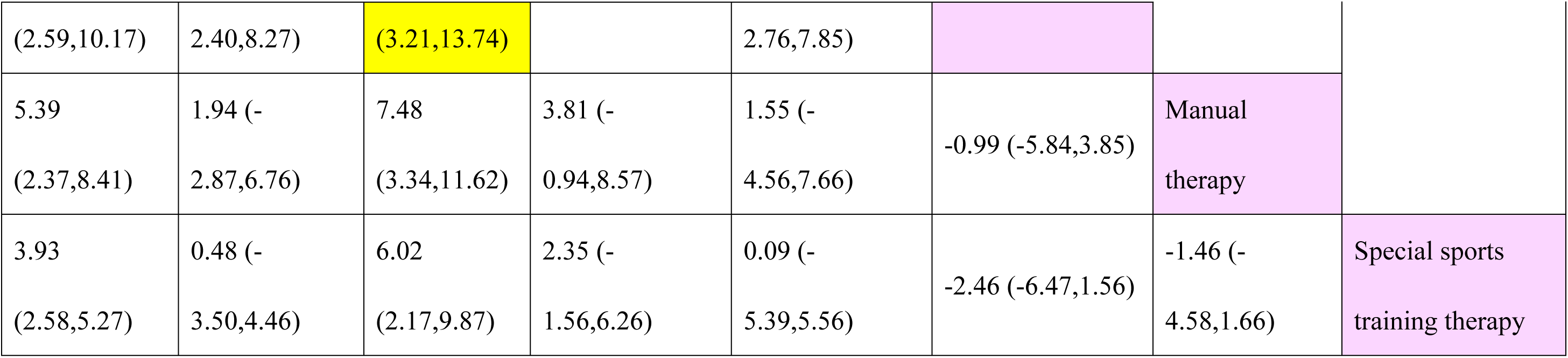
Comparison of the effects of different treatment methods on the internal rotation motion of shoulder joint.

|  |  |  |  |  |  |
| --- | --- | --- | --- | --- | --- |
| Exercise therapy |  |  |  |  |  |
| 3.45 (-0.31,7.20) | Electrical stimulation therapy |  |  |  |  |
| -2.09 (-5.75,1.56) | -5.54 (-10.78,-0.30) | Kinesio taping |  |  |  |
| 1.57 (-2.10,5.25) | -1.87 (-7.12,3.38) | 3.67 (-1.51,8.85) | Extracorporeal shock wave therapy |  |  |
| 3.84 (-1.47,9.15) | 0.39 (-6.11,6.89) | 5.93 (-0.51,12.38) | 2.26 (-3.87,8.40) | Microwave diathermy |  |
| 6.38 | 2.94 (- | 8.48 | 4.81 (0.36,9.26) | 2.54 (- | Acupuncture |

|  |  |  |  |  |  |  |  |
| --- | --- | --- | --- | --- | --- | --- | --- |
| (2.59,10.17) | 2.40,8.27) | (3.21,13.74) |  | 2.76,7.85) |  |  |  |
| 5.39<br>(2.37,8.41) | 1.94 (-<br>2.87,6.76) | 7.48<br>(3.34,11.62) | 3.81 (-<br>0.94,8.57) | 1.55 (-<br>4.56,7.66) | -0.99 (-5.84,3.85) | Manual<br>therapy |  |
| 3.93<br>(2.58,5.27) | 0.48 (-<br>3.50,4.46) | 6.02<br>(2.17,9.87) | 2.35 (-<br>1.56,6.26) | 0.09 (-<br>5.39,5.56) | -2.46 (-6.47,1.56) | -1.46 (-<br>4.58,1.66) | Special sports<br>training therapy |

### Ranking probability

Table 17 and Figs 17–20 summarize the ranking probabilities of different treatment approaches for the four main outcomes. According to SUCRA, medium-frequency therapy ranked first for VAS, acupuncture ranked first for shoulder flexion, Kinesio taping ranked first for ER, and manual therapy ranked first for IR. These ranking results should be interpreted together with the corresponding effect estimates and 95% CIs reported above, rather than being interpreted in isolation.

**Fig 17.**
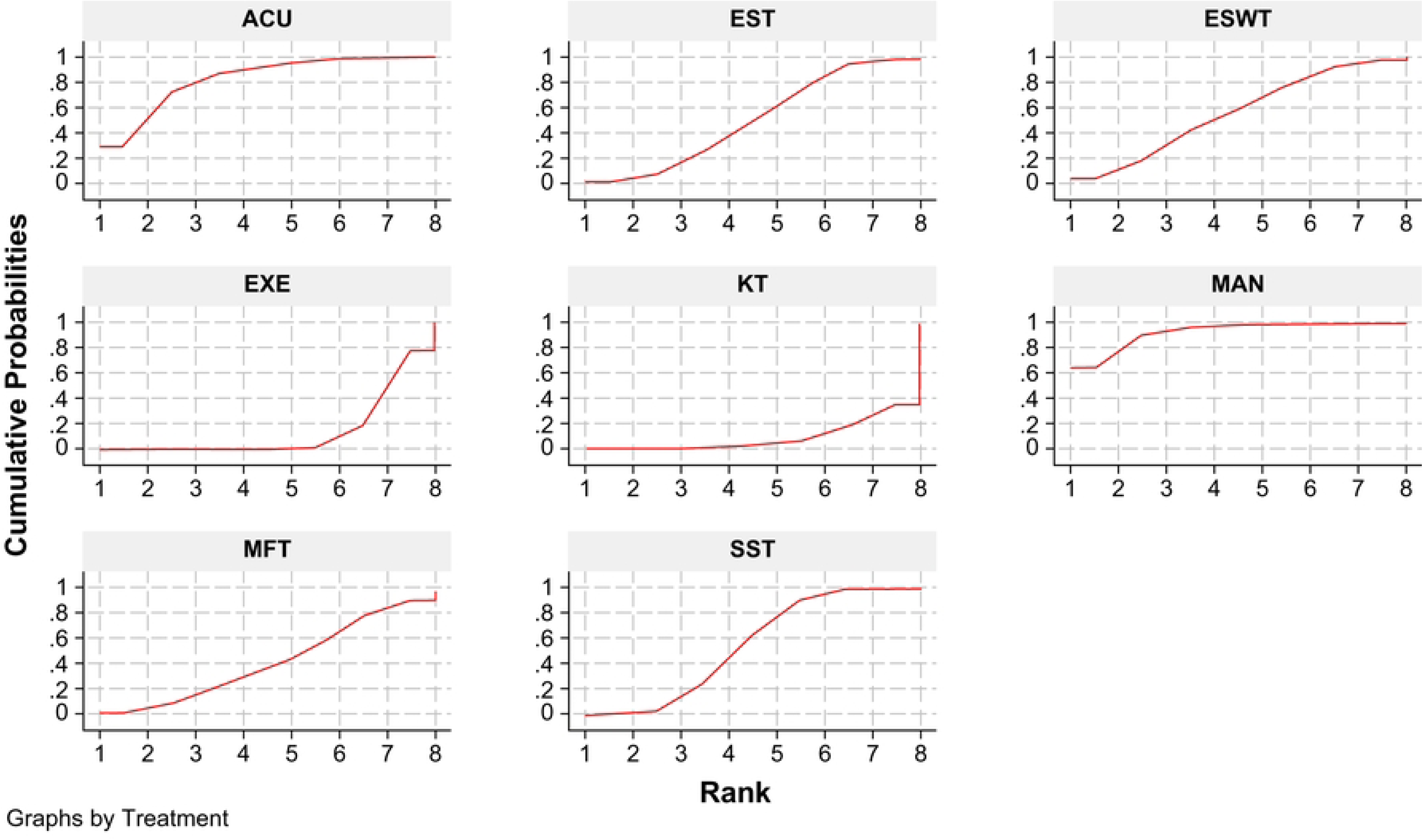
Visual analog scale treatment sequence diagram of patients with different treatment methods. ACU: acupuncture; EST: electrical stimulation therapy; EXE: exercise therapy; KT: Kinesio taping; MFT: medium-frequency therapy; MAN: manual therapy; MWD: microwave diathermy; ESWT: extracorporeal shock wave therapy; SST: special sports training therapy.

**Fig 18.**
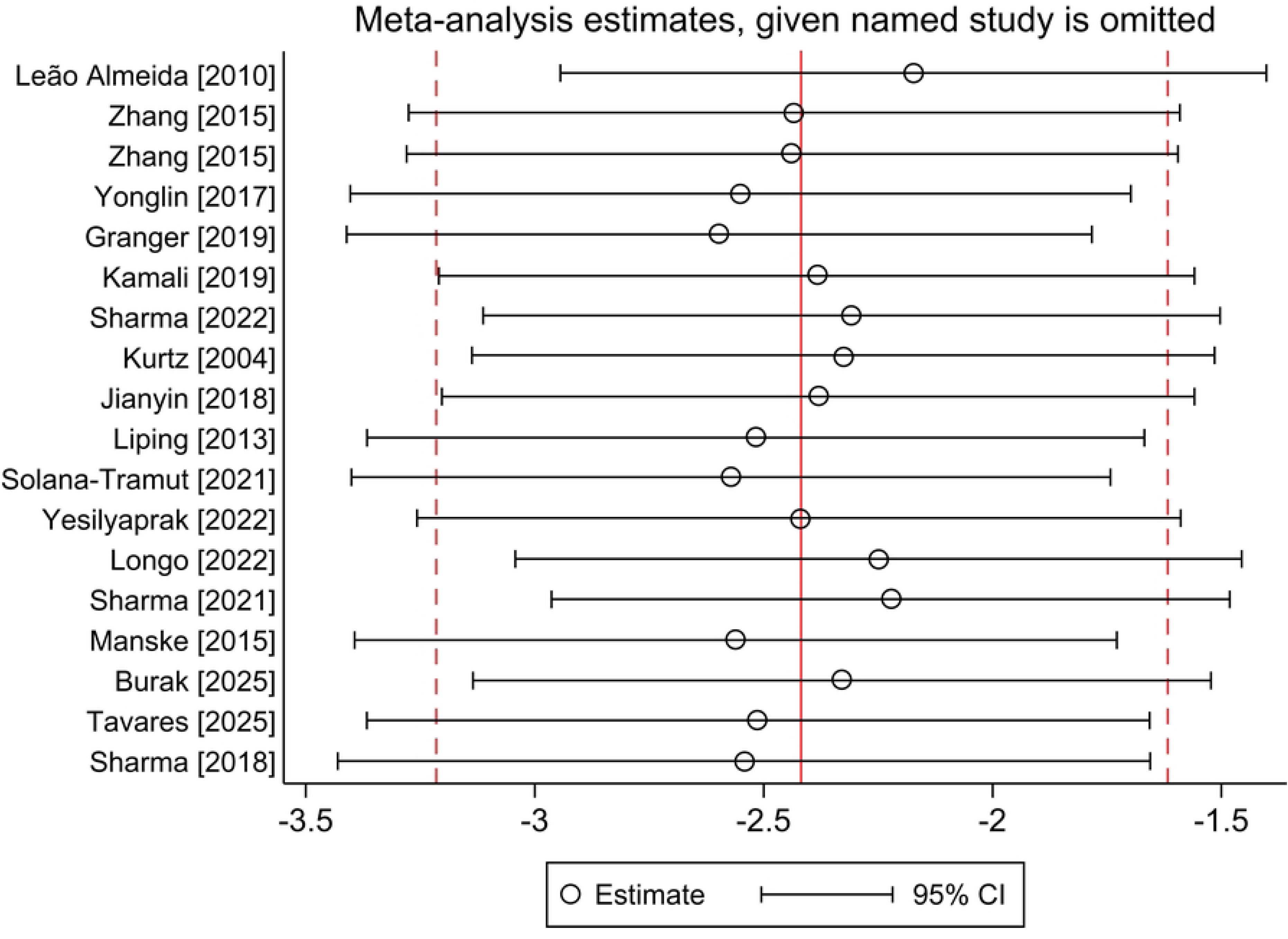
Sequence diagram of antexion treatment for patients with different treatment methods. ACU: acupuncture; EXE: exercise therapy; MAN: manual therapy; ESWT: extracorporeal shock wave therapy; SST: special sports training therapy.

**Fig 19.**
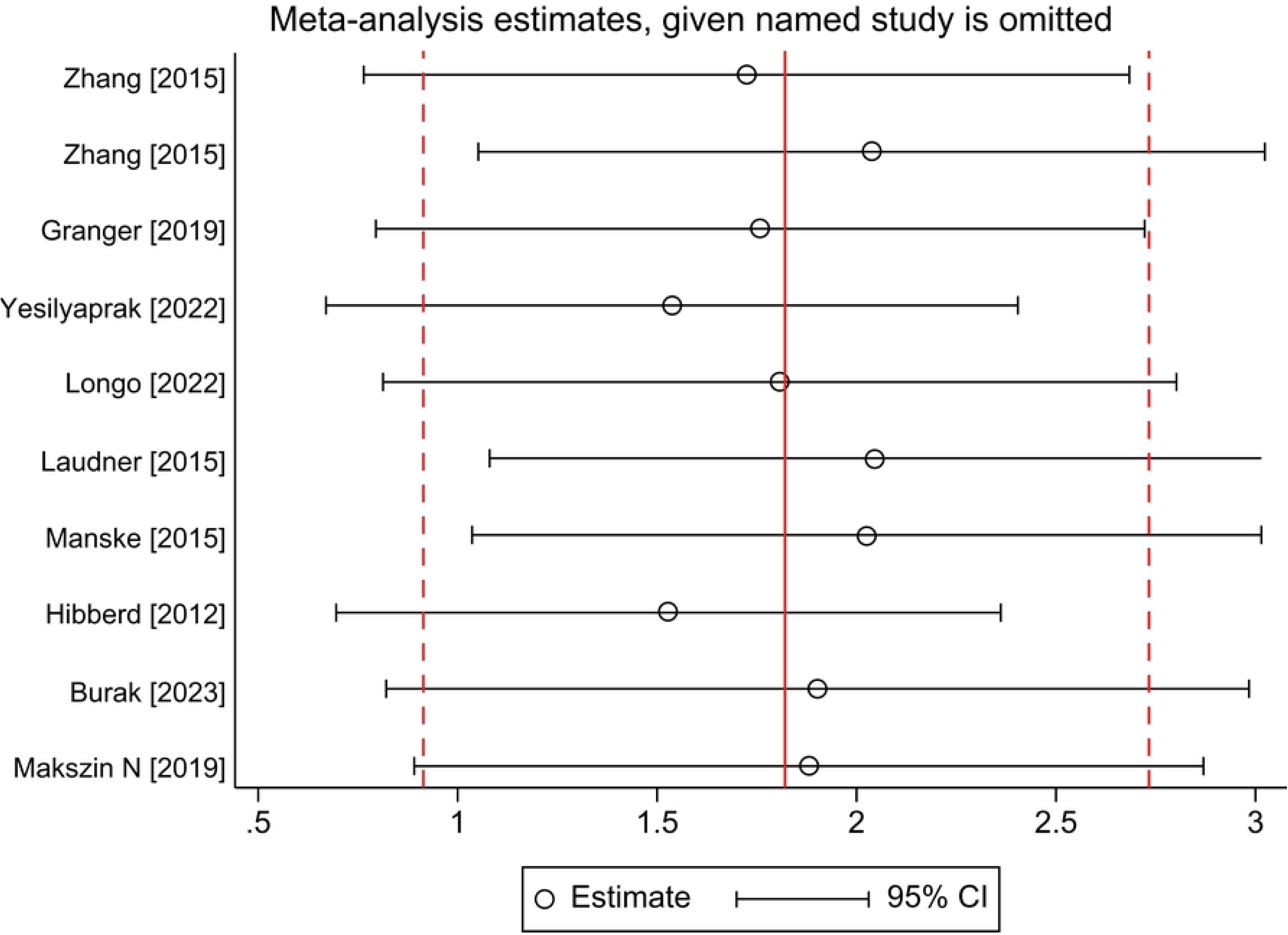
External rotation treatment sequencing diagram of patients with different treatment methods. ACU: acupuncture; EST: electrical stimulation therapy; EXE: exercise therapy; KT: Kinesio taping; MFT: medium-frequency therapy; MAN: manual therapy; MWD: microwave diathermy; ESWT: extracorporeal shock wave therapy; SST: special sports training therapy.

**Fig 20.**
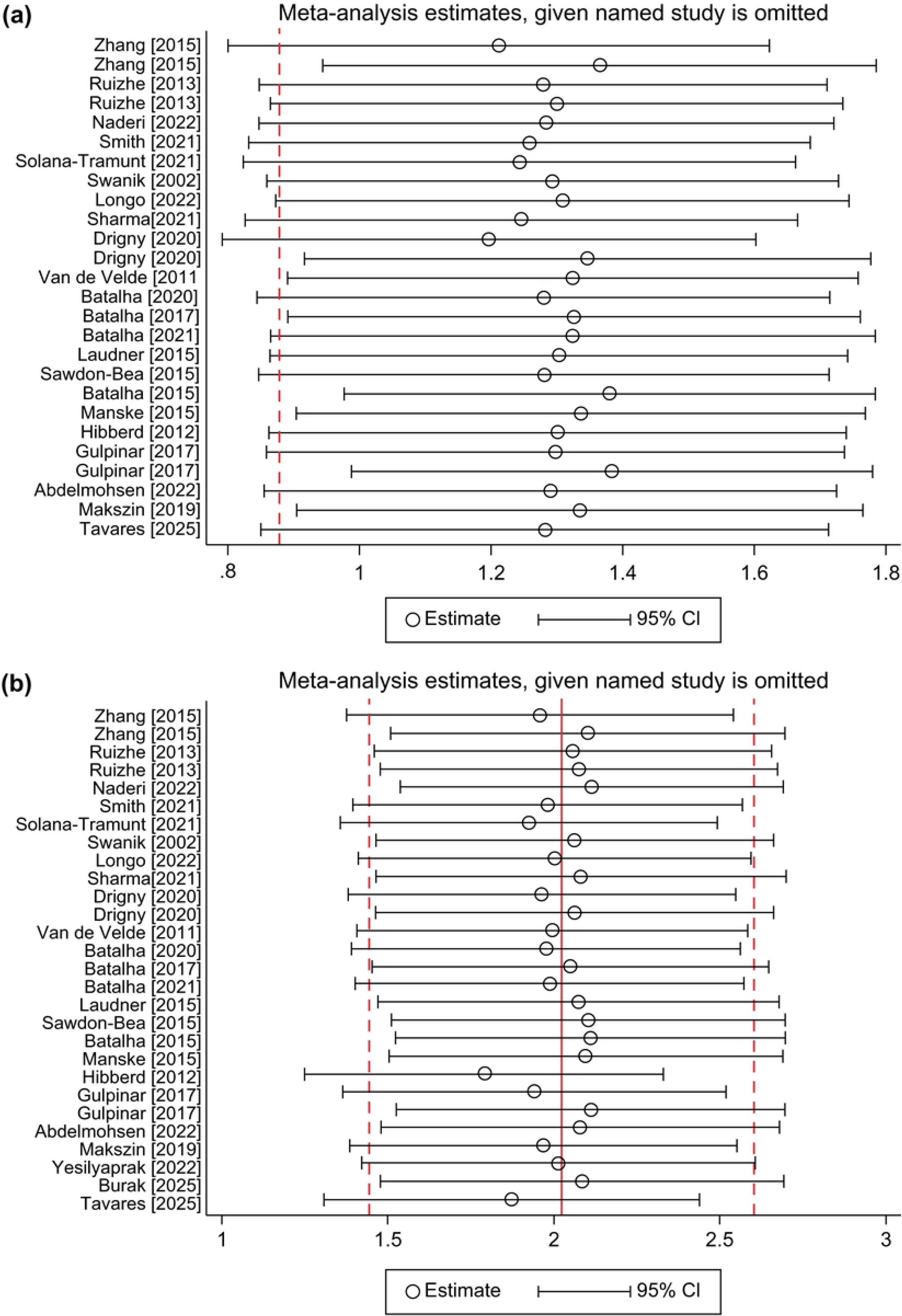
Internal rotation treatment sequence diagram of patients with different treatment methods. ACU: acupuncture; EST: electrical stimulation therapy; EXE: exercise therapy; KT: Kinesio taping; MFT: medium-frequency therapy; MAN: manual therapy; MWD: microwave diathermy; ESWT: extracorporeal shock wave therapy; SST: special sports training therapy.

**Table 17.**
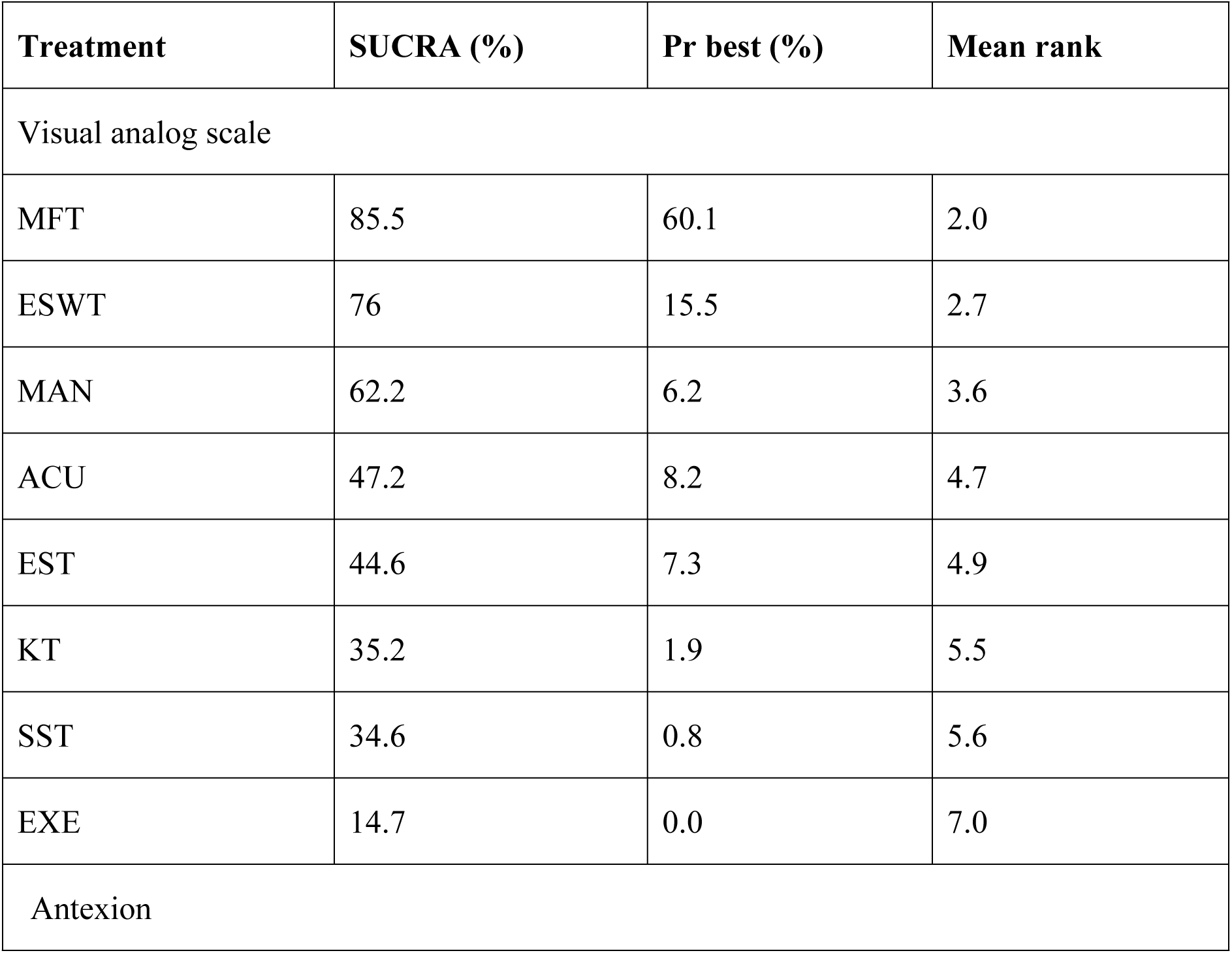
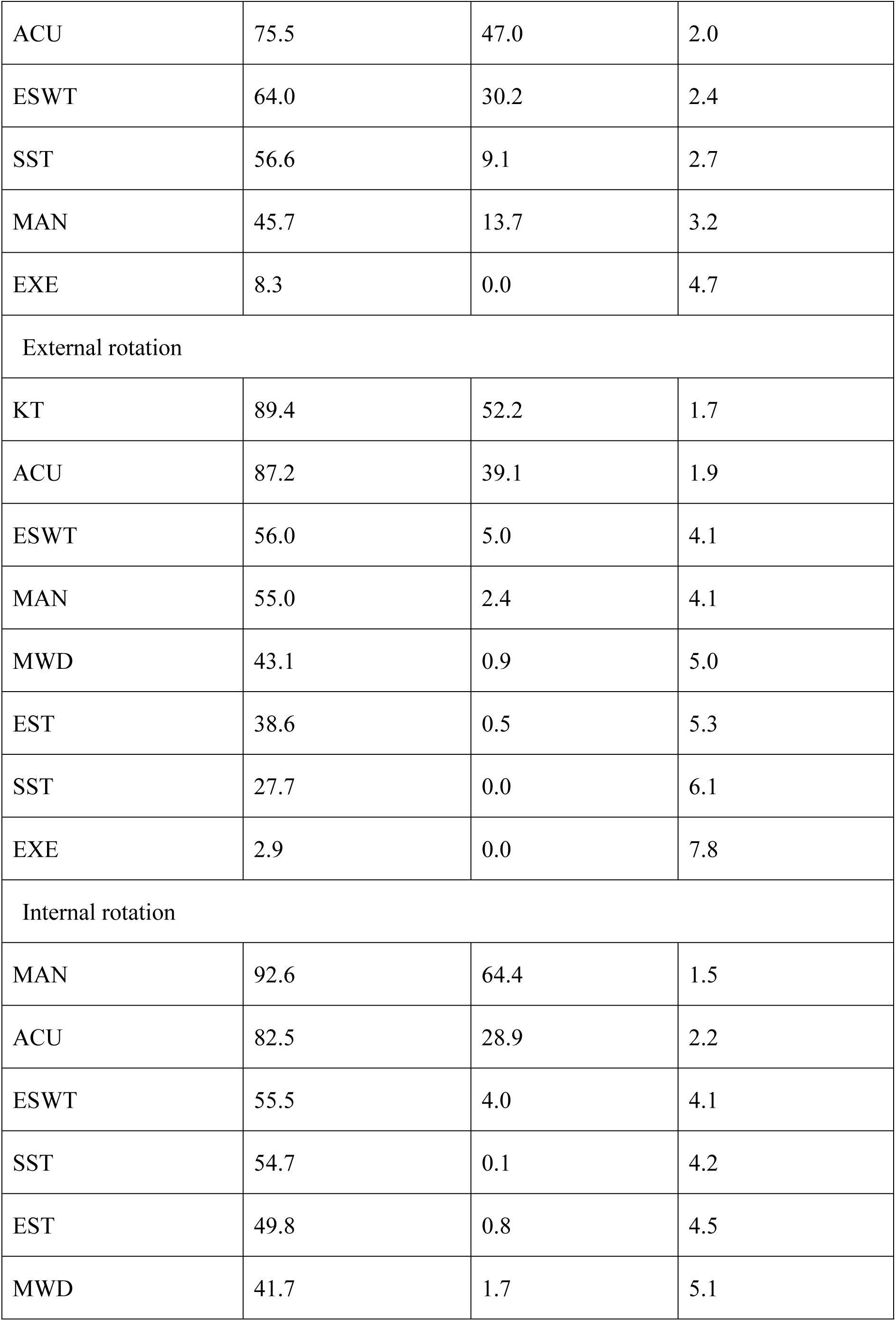
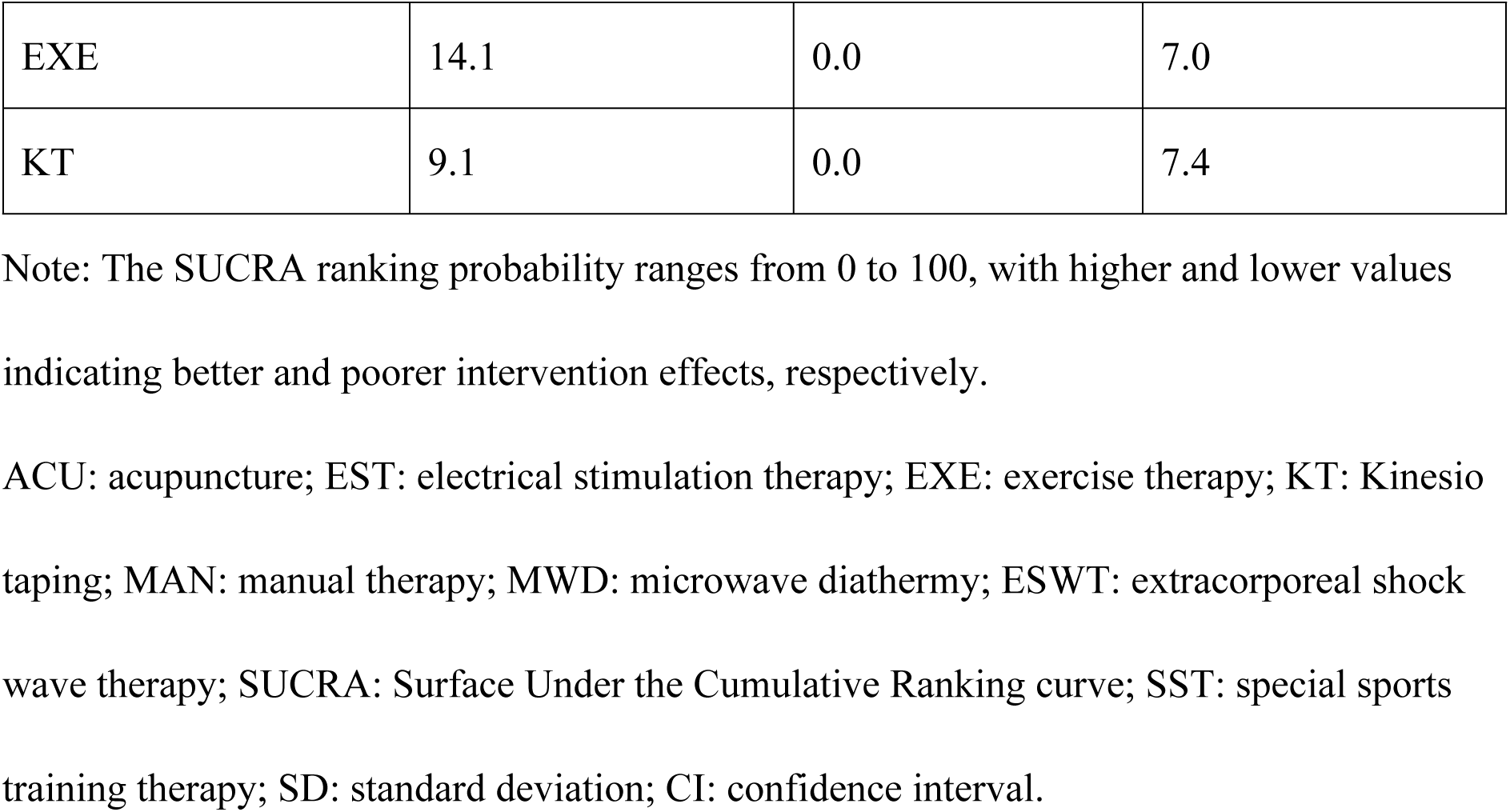
Ranking table of visual analog scale, antexion, external rotation, and internal rotation effects of different treatment methods.

### Sensitivity analysis and small-study effects

Sensitivity analyses suggested that the pooled estimates for the four primary outcomes were generally stable. For VAS, the pooled estimate remained centered at −2.44 (95% CI −3.23 to −1.64) after sequential omission of individual studies (Fig 21). A similar pattern was observed for shoulder flexion, with a pooled estimate of 1.83 (95% CI 0.85 to 2.81) (Fig 22). For shoulder rotational outcomes, the pooled estimate for ER remained approximately 1.29 (95% CI 0.86 to 1.72), whereas that for IR remained around 2.04 (95% CI 1.47 to 2.61) across leave-one-out analyses, indicating that these findings were not driven by any single study (Fig 23). Comparison-adjusted funnel plots revealed some degree of asymmetry across the four primary outcomes. Eight studies fell outside the funnel for VAS (Fig 24), five for shoulder flexion (Fig 25), ten for ER (Fig 26), and fifteen for IR (Fig 27), suggesting possible small-study effects. However, because funnel-plot asymmetry in network meta-analysis may also reflect heterogeneity and model complexity, these findings should be interpreted cautiously rather than as definitive evidence of publication bias.

**Fig 21.**
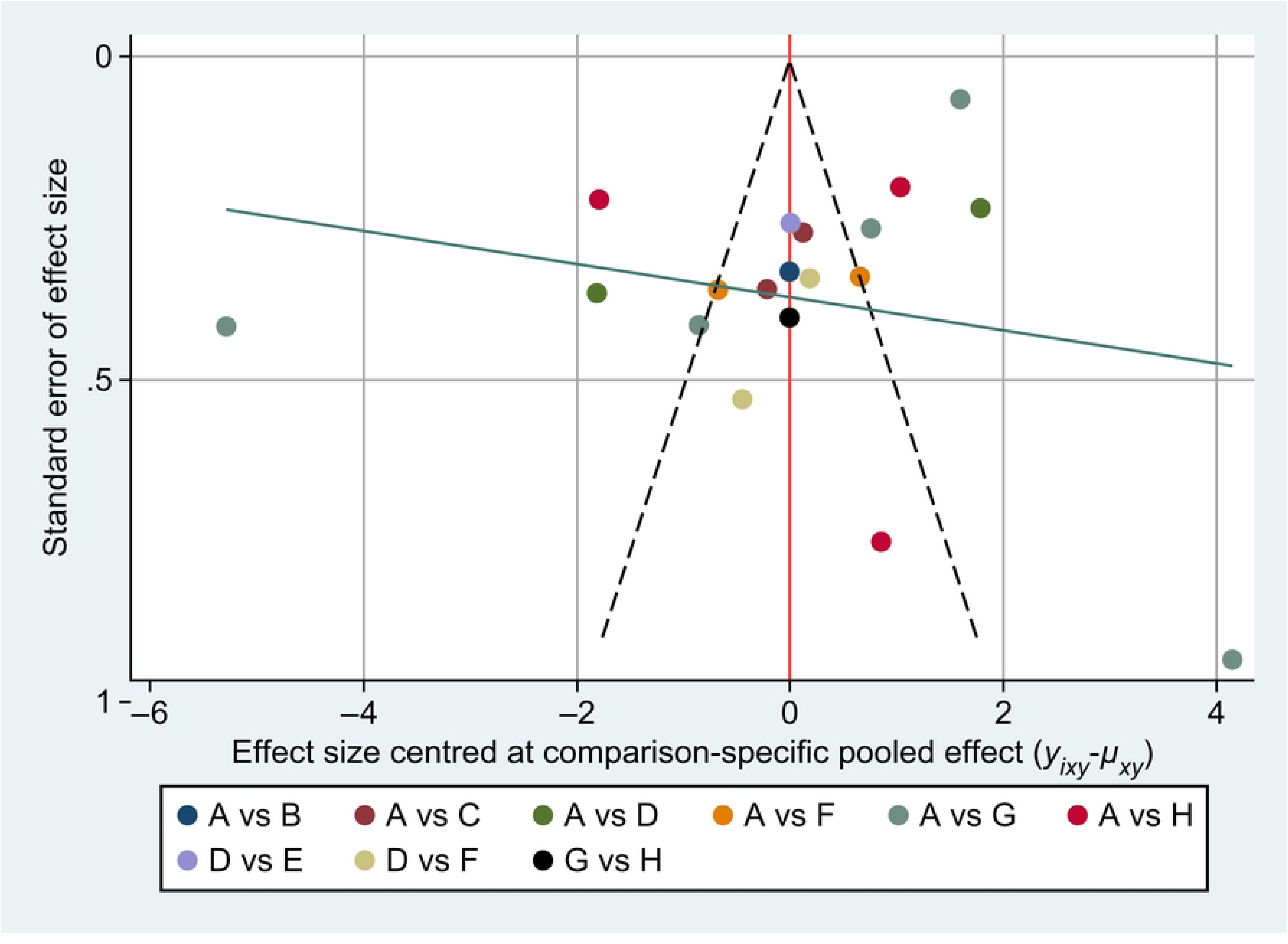
Sensitivity analysis of the visual analog scale. CI, confidence interval

**Fig 22.**
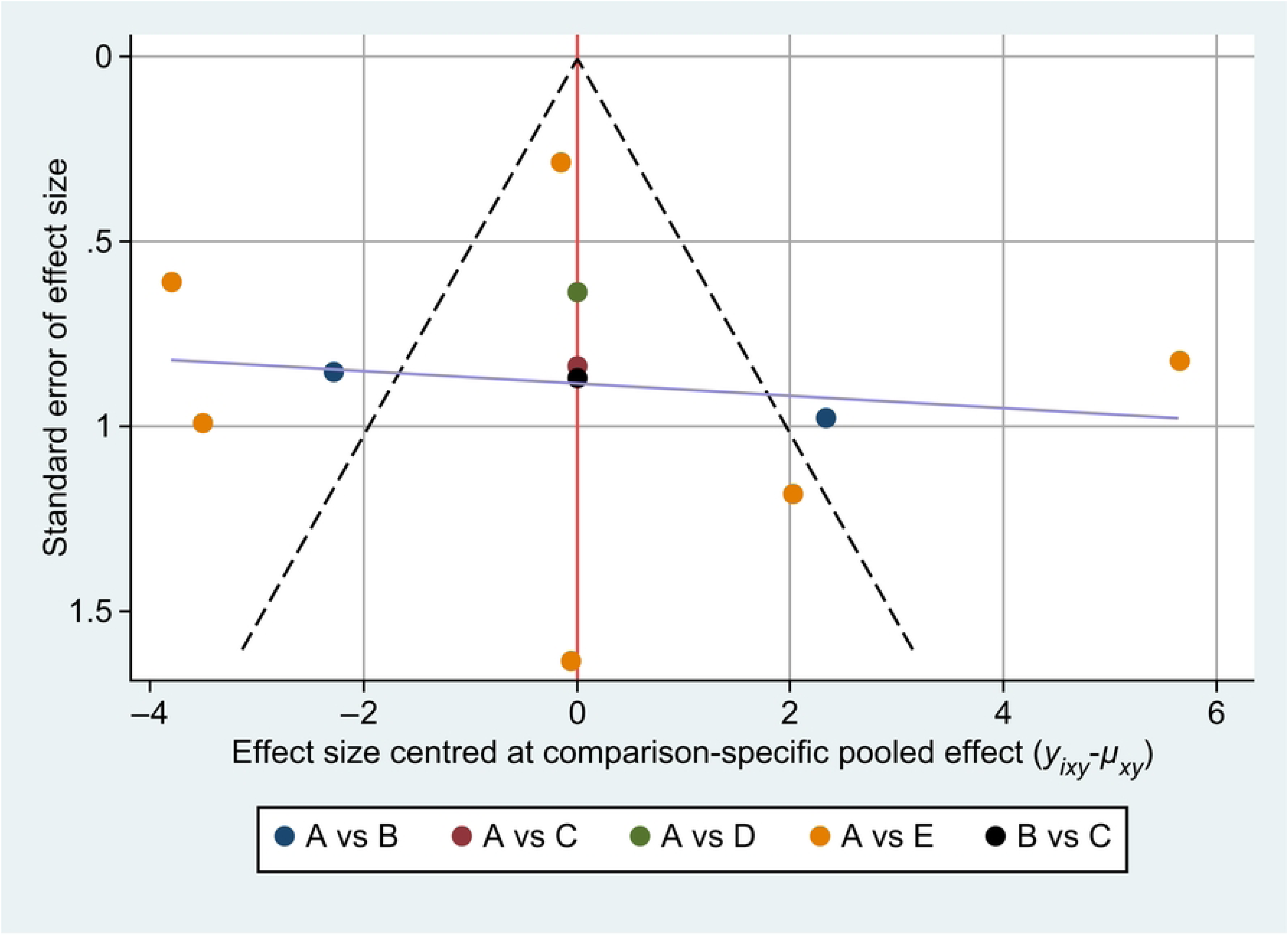
Sensitivity analysis of shoulder joint flexion motion. CI: confidence interval.

**Fig 23.**
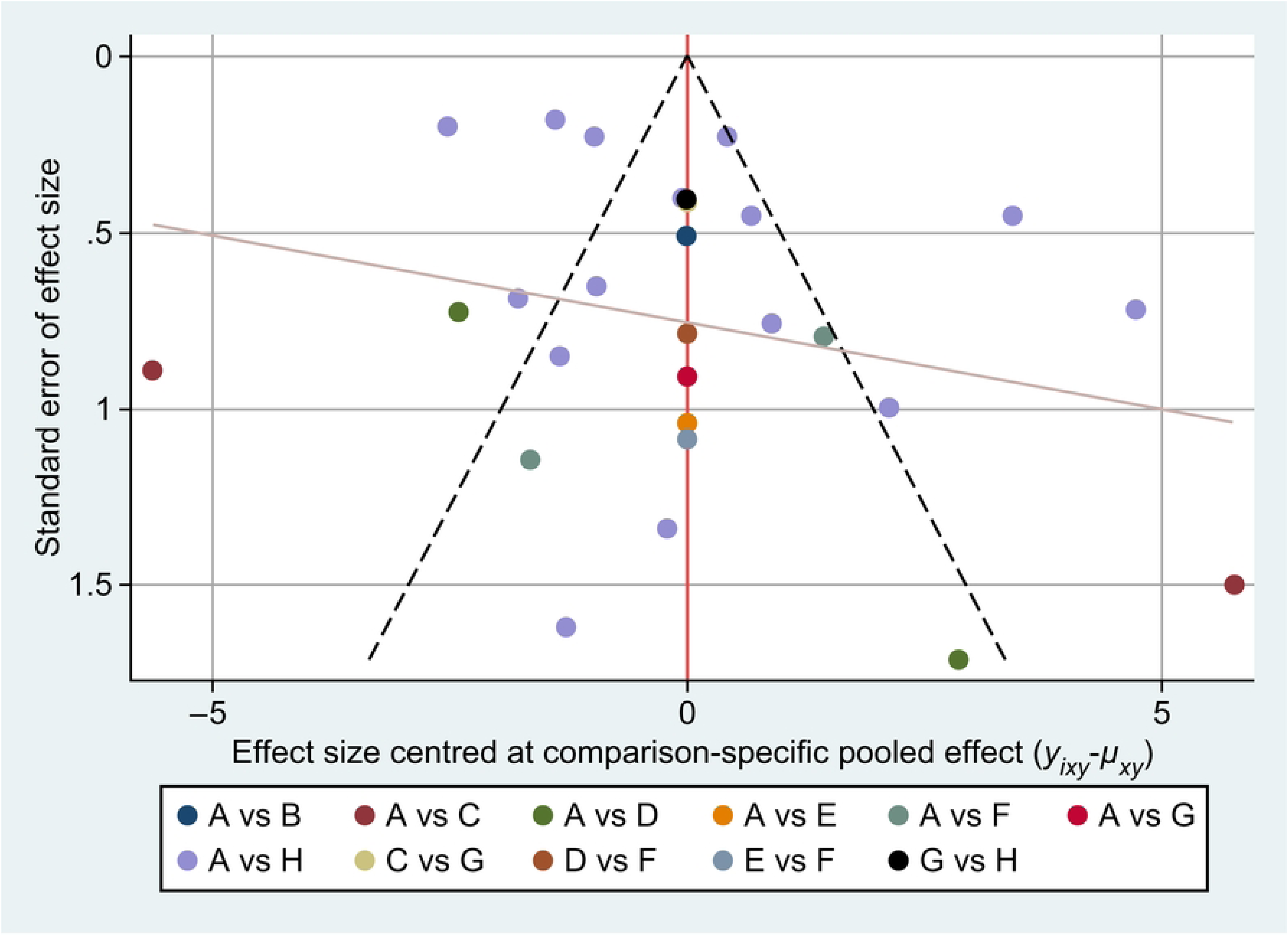
Sensitivity analyses of shoulder internal and external rotation range of motion. (a) Sensitivity analysis of external rotation motion of shoulder joint. (b) Sensitivity analysis of internal rotation motion of shoulder joint. CI: confidence interval.

**Fig 24.**
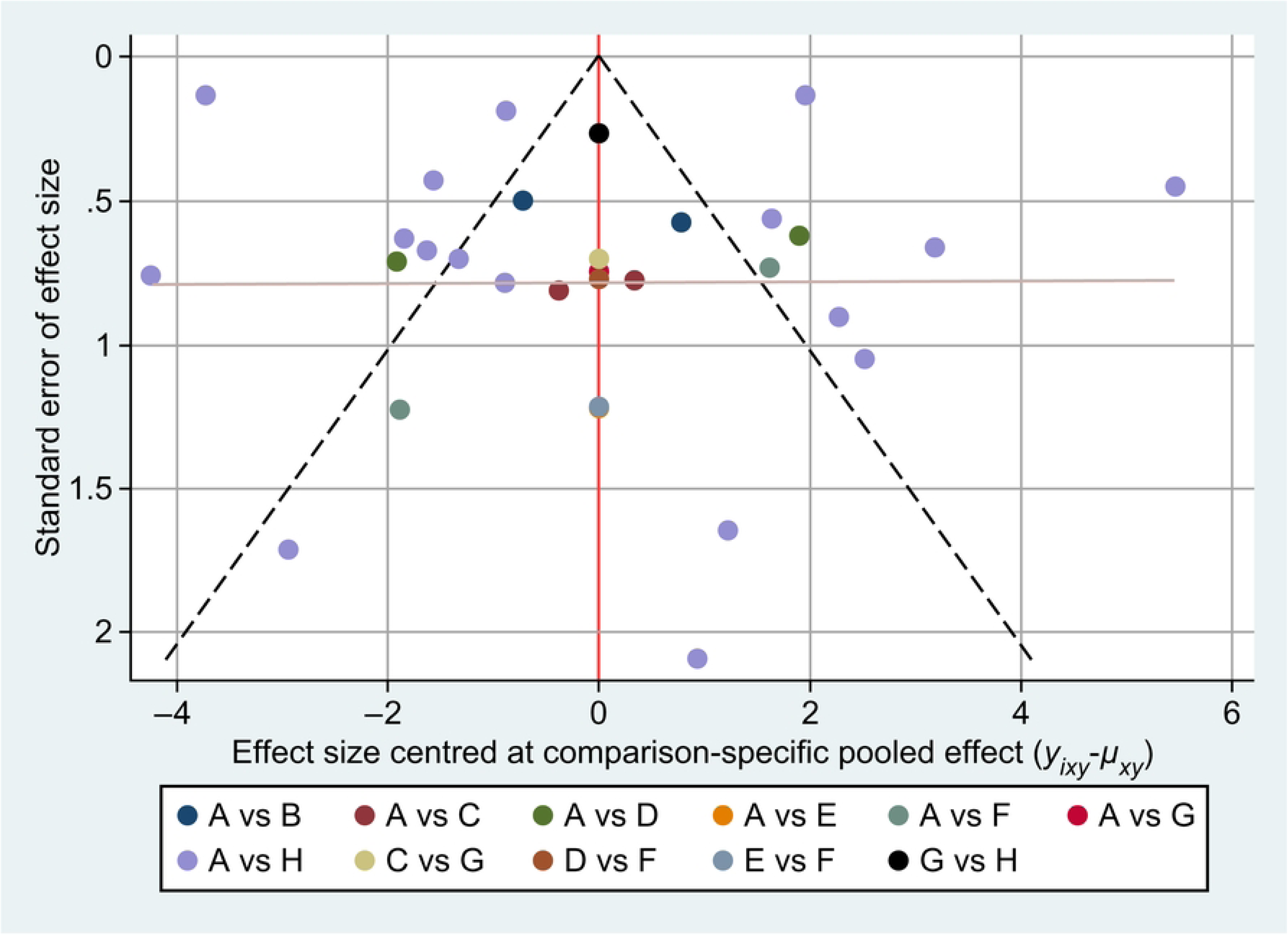
Visual analog scale funnel diagram.

**Fig 25.**
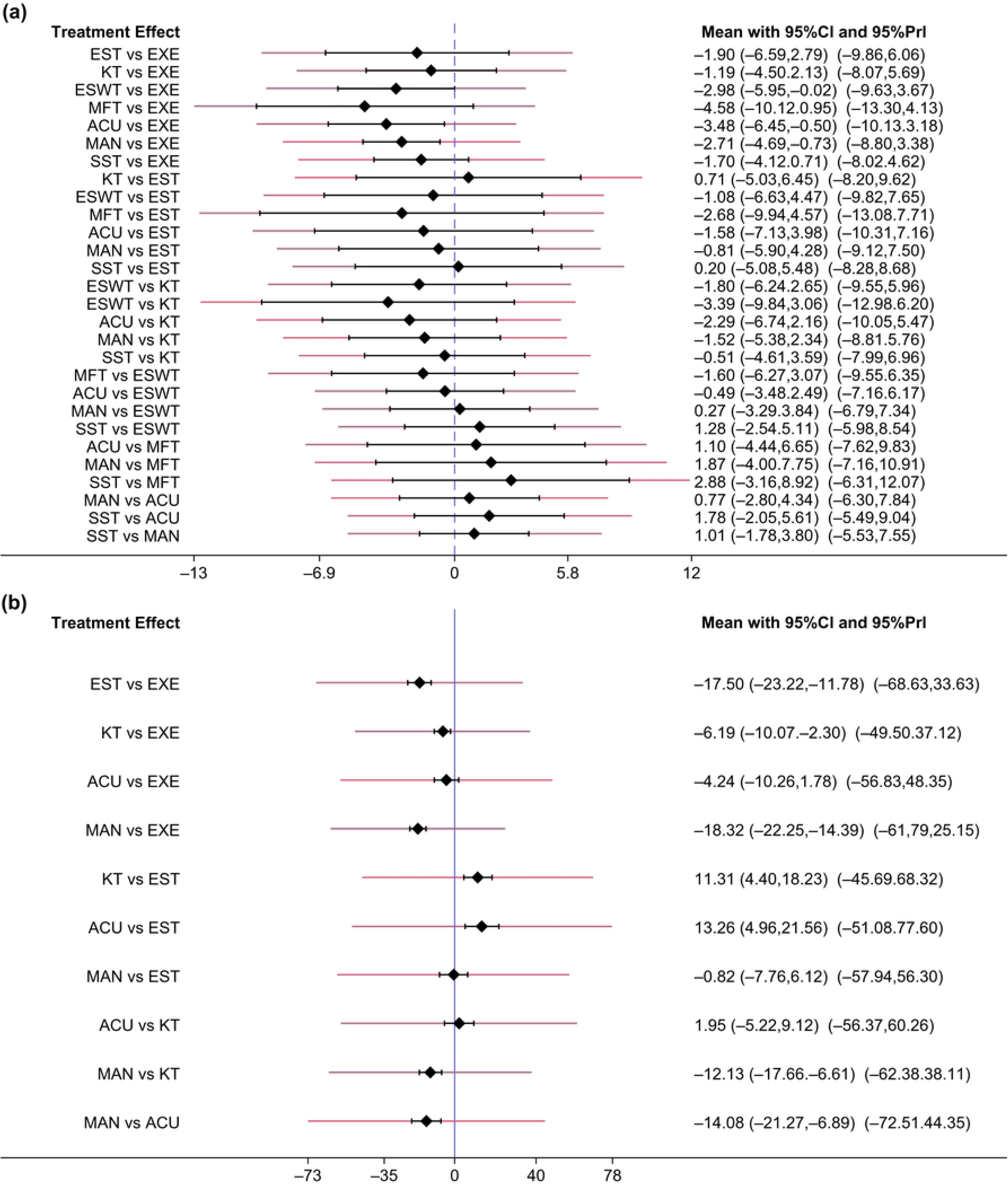
Funnel chart of shoulder joint forward flexion range of motion.

**Fig 26.**
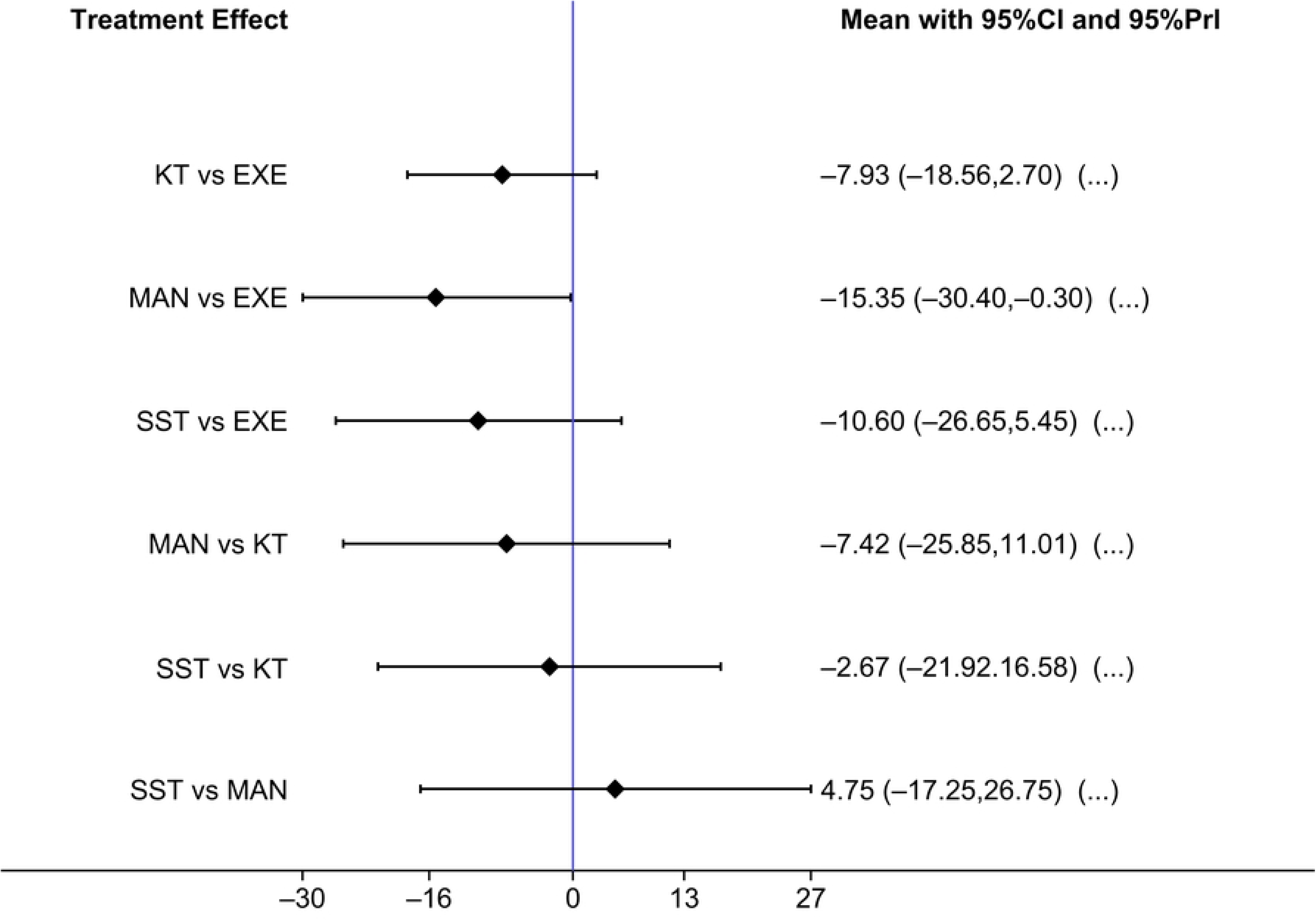
Funnel chart of external rotation range of motion of shoulder joint.

**Fig 27.**
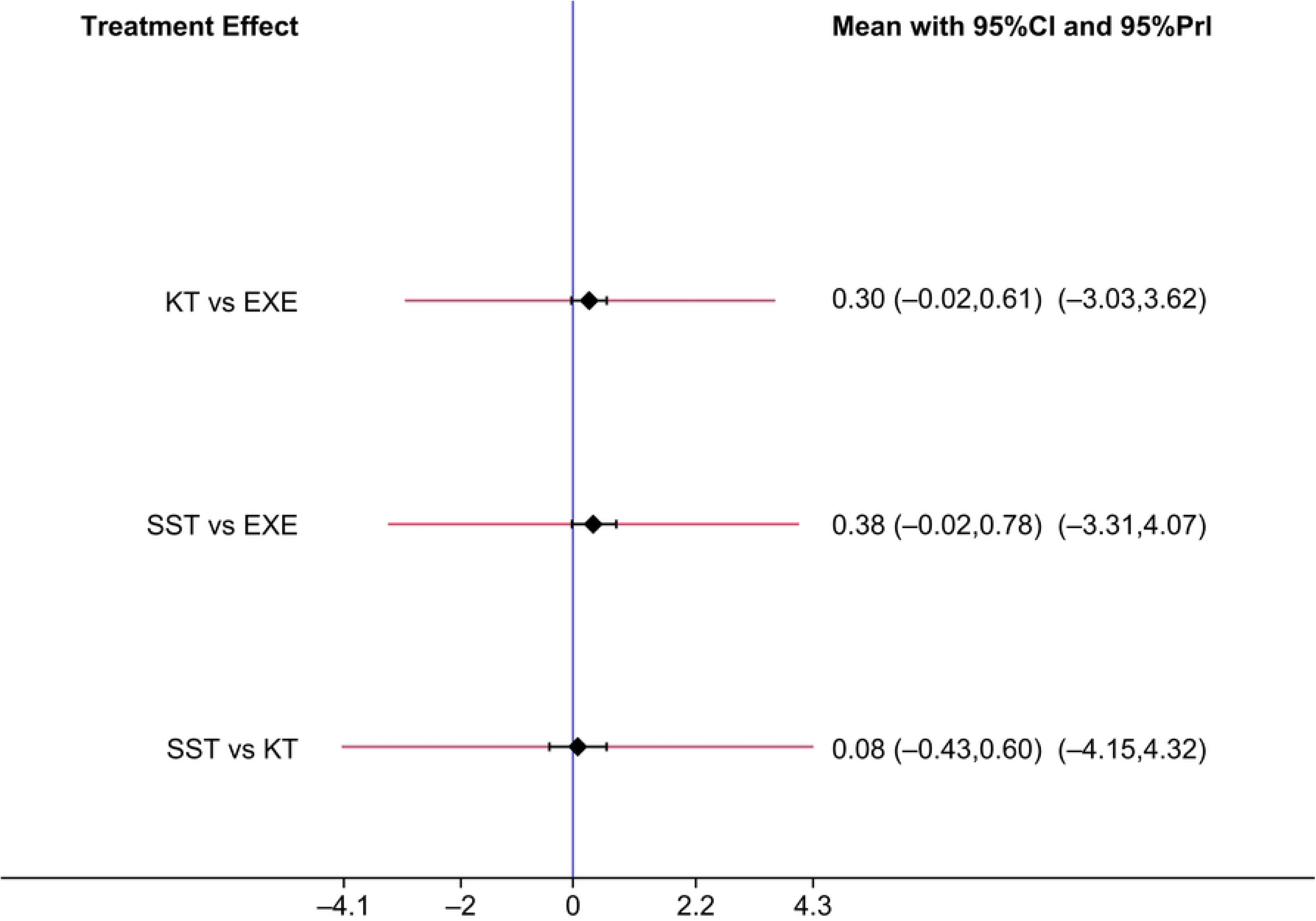
Funnel chart of external rotation range of motion of shoulder joint.

### Pairwise comparisons for secondary outcomes

Pairwise league-table analyses were further performed for secondary outcomes, including shoulder pain and disability, upper-limb function, structural indicators, muscle-performance indices, and additional range-of-motion outcomes.

### Shoulder pain and disability (SPADI)

Table 18 demonstrates the pairwise comparisons of treatment effects on SPADI in 296 swimmers with shoulder impingement syndrome. Acupuncture plus exercise therapy showed a favorable estimate compared with electrical stimulation therapy plus exercise therapy (MD 13.26, 95% CI 4.96 to 21.56), although the corresponding prediction interval was wide. This result suggests a potential benefit of acupuncture-based treatment for pain and disability, but the magnitude of the effect should be interpreted with caution.

**Table 18.** Pair-to-pair comparison of the effects of different treatment methods on the index of shoulder joint pain and dysfunction.

| Exercise therapy |  |  |  |  |
| --- | --- | --- | --- | --- |
| -17.50 (-23.22, -11.78) | Electrical stimulation therapy |  |  |  |
| -6.19 (-10.07, -2.30) | 11.31 (4.40, 18.23) | Kinesio taping |  |  |
| -4.24 (-10.26, 1.78) | 13.26 (4.96, 21.56) | 1.95 (-5.22, 9.12) | Acupuncture |  |
| -18.32 (-22.25, -14.39) | -0.82 (-7.76, 6.12) | -12.13 (-17.66, -6.61) | -14.08 (-21.27, -6.89) | Manual therapy |

### Upper-limb function (DASH)

For upper-limb function assessed by DASH, the available evidence was limited and the pairwise estimates were imprecise. As demonstrated in Table 19 and Fig 28, the comparison between special sports training and manual therapy yielded an MD of 4.75 (95% CI −17.25 to 26.75), indicating substantial uncertainty. Similarly, the comparisons of Kinesio taping versus exercise therapy (MD −7.93, 95% CI −18.56 to 2.70), manual therapy versus exercise therapy (MD −15.35, 95% CI −30.40 to −0.30), and special sports training versus exercise therapy (MD −10.60, 95% CI −26.65 to 5.45) were associated with wide CIs. Overall, the available evidence for DASH remains limited and should be interpreted cautiously.

**Fig 28.**
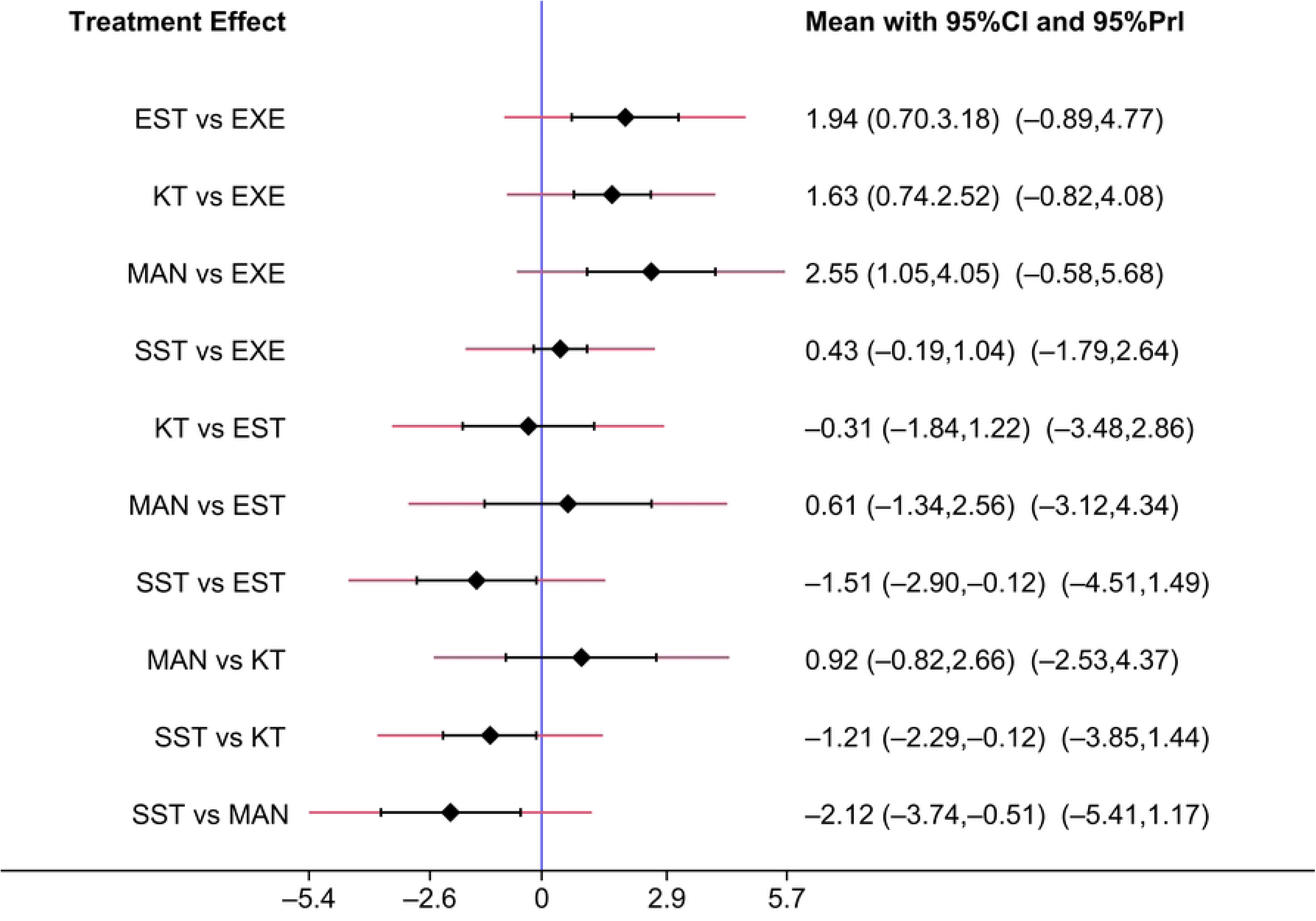
Pairwise comparison of different treatment methods on upper limb function evaluation of patients. EXE: exercise therapy; KT: Kinesio taping; MAN: manual therapy; ESWT: extracorporeal shock wave therapy; SST: special sports training therapy; CI: confidence interval.

**Table 19.** A pairwise comparison of the effects of different treatment methods on the evaluation of upper-limb function.

|  |  |  |  |
| --- | --- | --- | --- |
| Exercise therapy |  |  |  |
| -7.93 (-18.56, 2.70) | Kinesio taping |  |  |
| -15.35 (-30.40, -0.30) | -7.42 (-25.85, 11.01) | Manual therapy |  |
| -10.60 (-26.65, 5.45) | -2.67 (-21.92, 16.58) | 4.75 (-17.25, 26.75) | Special sports training therapy |

### Structural indicators

For the PMI, Table 20 and Fig 29 demonstrate that special sports training yielded the most favorable estimate relative to exercise therapy (MD 0.38, 95% CI −0.02 to 0.78), whereas Kinesio taping versus exercise therapy showed a smaller estimate (MD 0.30, 95% CI −0.02 to 0.61). The comparison between special sports training and Kinesio taping was minimal (MD 0.08, 95% CI −0.43 to 0.60). Taken together, these findings suggest a possible advantage of special sports training for PMI, although the CIs indicate limited precision.

**Fig 29.**
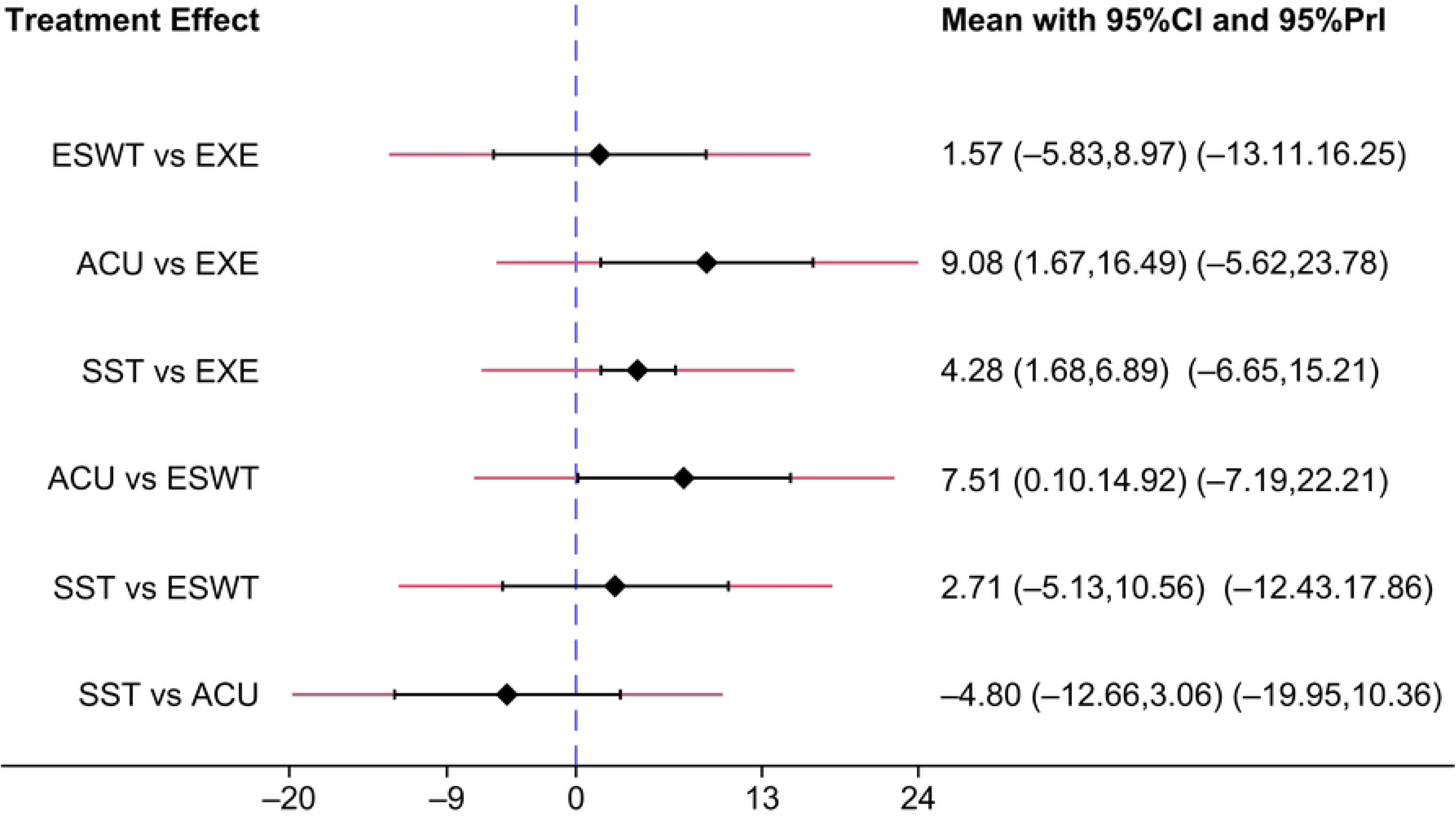
Pairwise comparison of different treatment methods on patients with pectoralis minor muscle index. EXE: exercise therapy; KT: Kinesio taping; SST: special sports training therapy; CI: confidence interval.

**Table 20.**
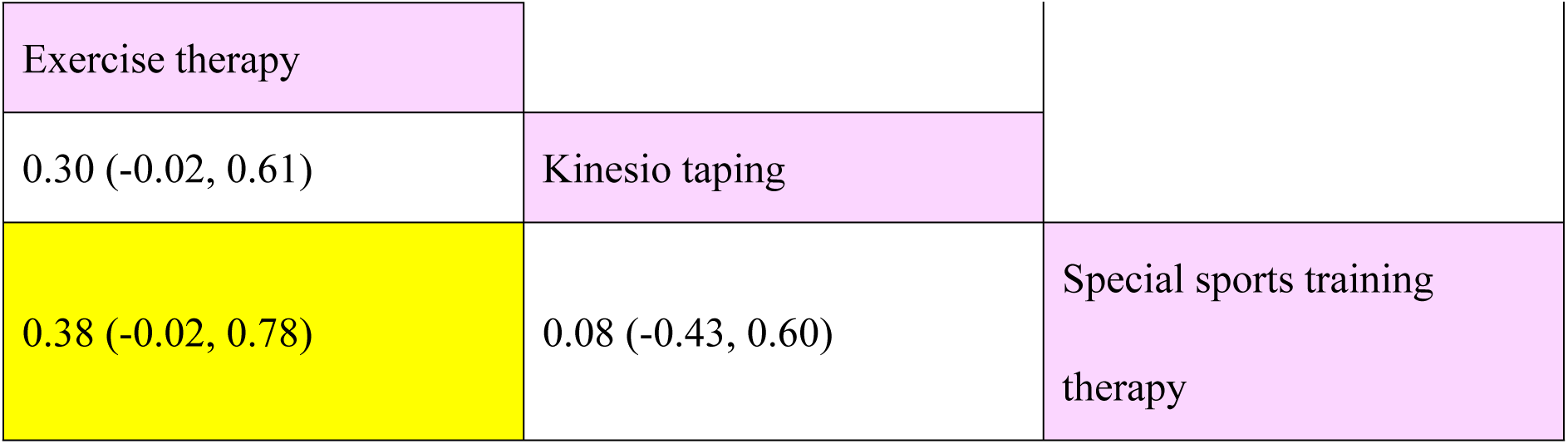
Pairwise comparison of the effect of different treatments on the pectoralis minor muscle index of patients.

For acromiohumeral distance, Table 21 and Fig 30 demonstrate that manual therapy plus exercise therapy yielded the largest estimated improvement compared with exercise therapy (MD 2.55, 95% CI 1.05 to 4.05). Electrical stimulation therapy (MD 1.94, 95% CI 0.70 to 3.18) and Kinesio taping (MD 1.63, 95% CI 0.74 to 2.52) also showed favorable estimates relative to exercise therapy, whereas special sports training showed a smaller and non-significant estimate (MD 0.43, 95% CI −0.19 to 1.04). These findings suggest that manual therapy may be more beneficial than the other evaluated interventions for improving acromiohumeral distance.

**Fig 30.**
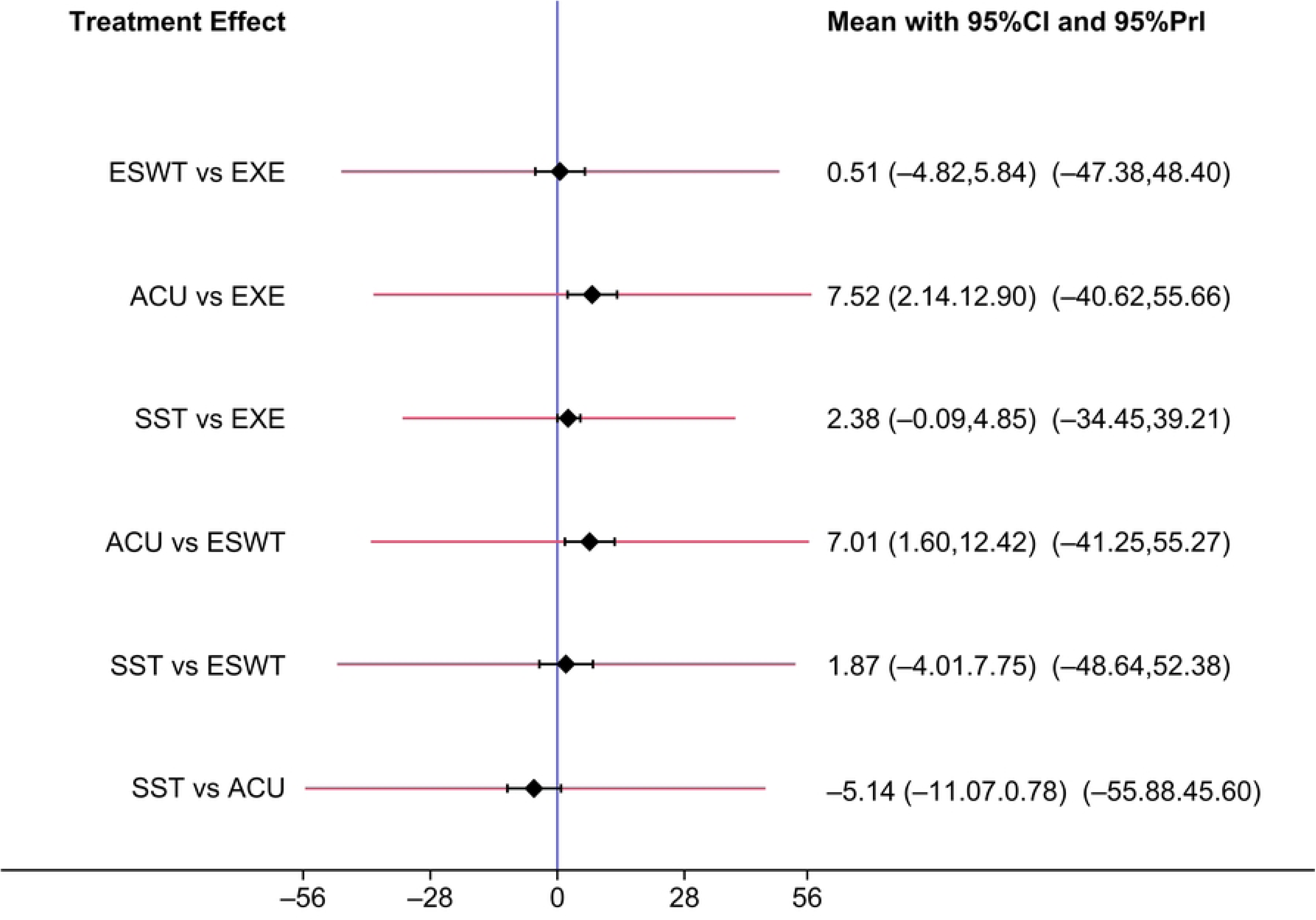
Pairwise comparison of different treatment methods on the treatment of acromial humeral distance in patients. EST: electrical stimulation therapy; EXE: exercise therapy; KT: Kinesio taping; MFT: medium-frequency therapy; MAN: manual therapy; SST: special sports training therapy; CI: confidence interval.

**Table 21.**
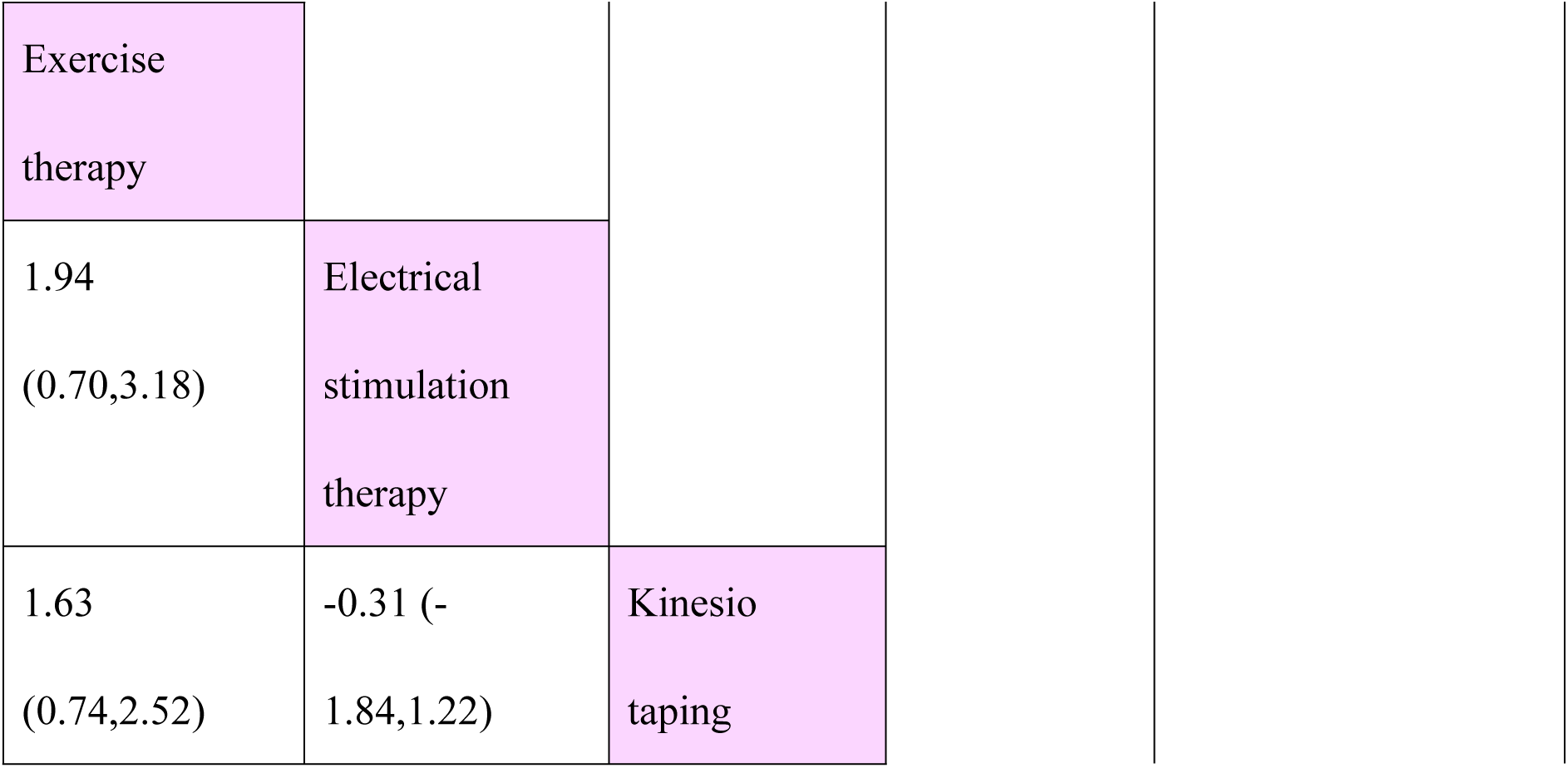
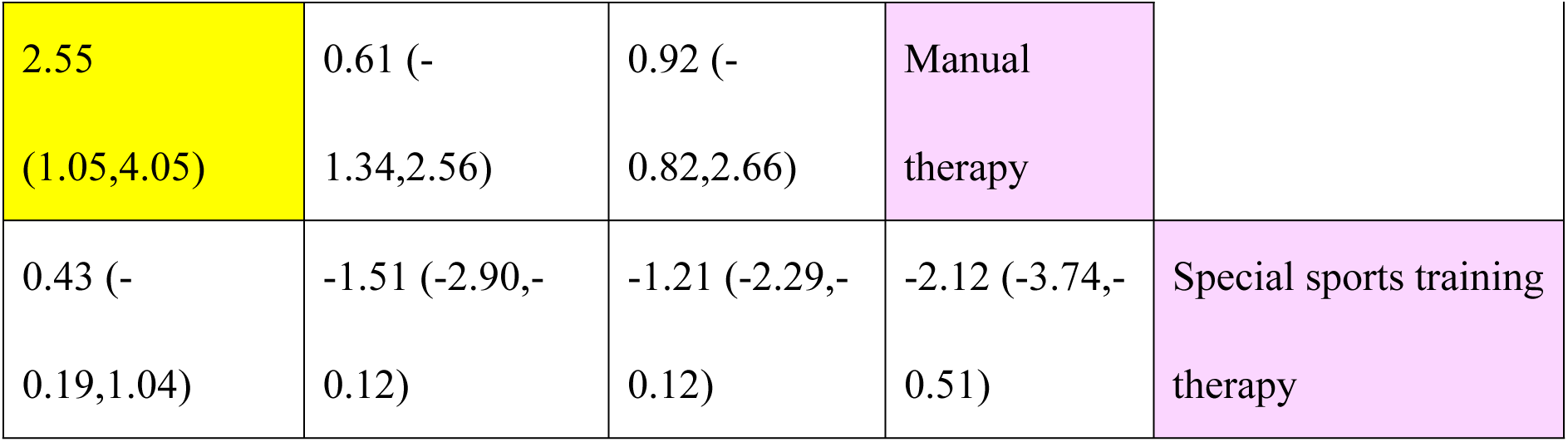
Pair-to-pair comparison of the effects of different treatments on acromial humeral distance in patients.

|  |  |  |  |  |
| --- | --- | --- | --- | --- |
| Exercise therapy |  |  |  |  |
| 1.94<br>(0.70,3.18) | Electrical stimulation therapy |  |  |  |
| 1.63<br>(0.74,2.52) | -0.31 (-1.84,1.22) | Kinesio taping |  |  |
| 2.55<br>(1.05,4.05) | 0.61 (-<br>1.34,2.56) | 0.92 (-<br>0.82,2.66) | Manual<br>therapy |  |
| 0.43 (-<br>0.19,1.04) | -1.51 (-2.90,-<br>0.12) | -1.21 (-2.29,-<br>0.12) | -2.12 (-3.74,-<br>0.51) | Special sports training<br>therapy |

### Muscle-performance indicator

For the IR/ER strength ratio, Table 22 and Fig 31 demonstrate that acupuncture plus exercise therapy yielded the largest estimated benefit relative to exercise therapy (MD 9.08, 95% CI 1.67 to 16.49), followed by special sports training versus exercise therapy (MD 4.28, 95% CI 1.68 to 6.89). The comparison between acupuncture and extracorporeal shock wave therapy also favored acupuncture (MD 7.51, 95% CI 0.10 to 14.92). Although these point estimates suggest a favorable profile for acupuncture, the corresponding prediction intervals remained wide, indicating uncertainty in the expected effect across future studies.

**Fig 31.**
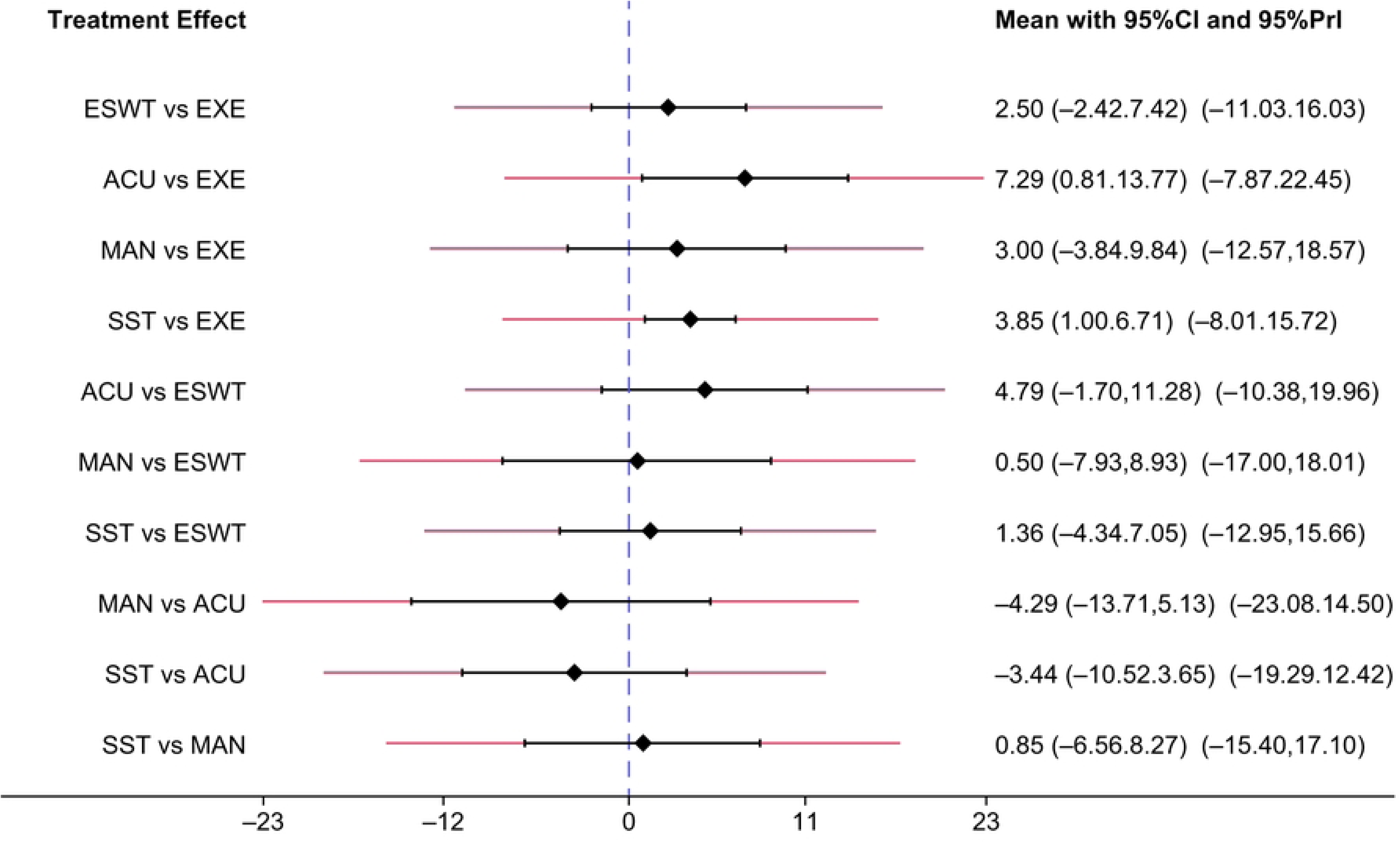
Pairwise comparison of treatment methods on internal and external rotation muscle strength of shoulder joint. ACU: acupuncture; ESWT: extracorporeal shock wave therapy; SST: special sports training therapy; CI: confidence interval.

**Table 22.**
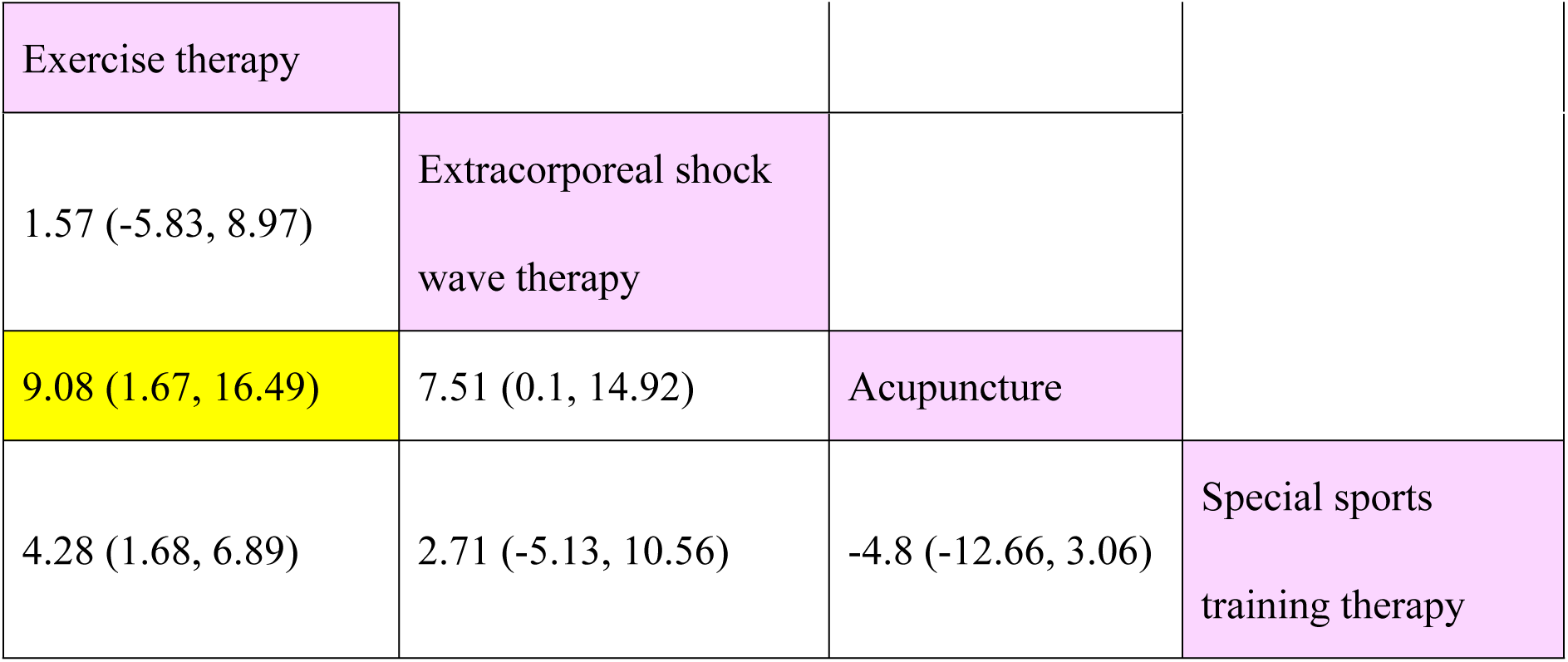
Effect of different treatment methods on the ratio of internal and external rotation muscle strength of shoulder joint.

### Additional range-of-motion outcomes

For shoulder extension, Table 23 and Fig 32 demonstrate that acupuncture plus exercise therapy yielded the most favorable estimate relative to exercise therapy (MD 7.52, 95% CI 2.14 to 12.90), whereas special sports training versus exercise therapy showed a smaller and non-significant estimate (MD 2.38, 95% CI −0.09 to 4.85). The comparison between acupuncture and extracorporeal shock wave therapy also favored acupuncture (MD 7.01, 95% CI 1.60 to 12.42). However, the corresponding prediction intervals were wide, indicating substantial uncertainty despite the favorable point estimates.

**Fig 32.**
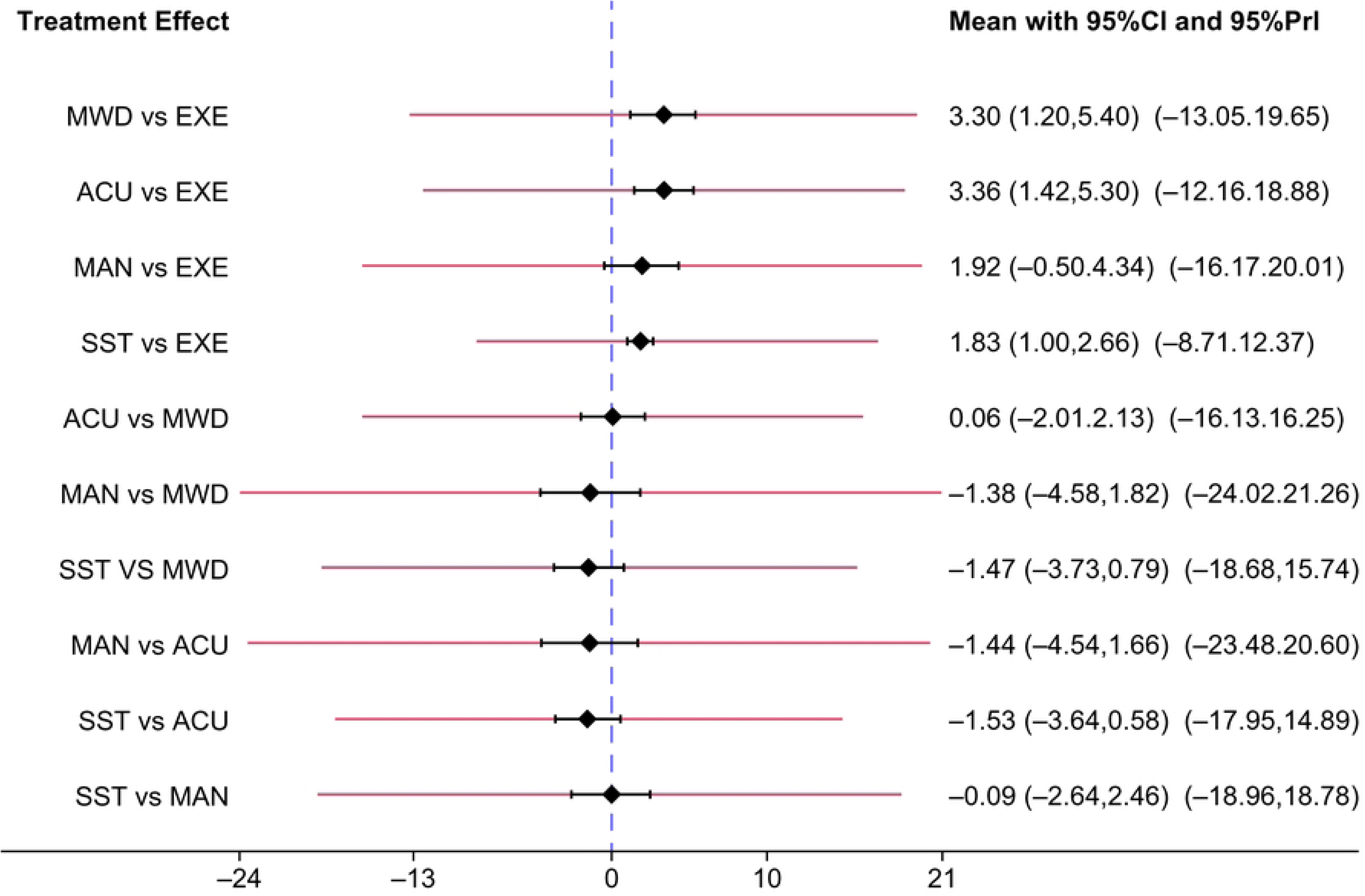
Pairwise comparison of the effects of different treatments on the posterior extension range of shoulder. ACU: acupuncture; EXE: exercise therapy; ESWT: extracorporeal shock wave therapy; SST: special sports training therapy; CI: confidence interval.

**Table 23.** Pairwise comparison of the influence of different treatment methods on the posterior extension range of shoulder joint.

|  |  |  |  |
| --- | --- | --- | --- |
| Exercise therapy |  |  |  |
| 0.51 (-4.82,5.84) | Extracorporeal shock wave therapy |  |  |
| 7.52 (2.14, 12.90) | 7.01 (1.60, 12.42) | Acupuncture |  |
| 2.38 (-0.09, 4.85) | 1.87 (-4.01, 7.75) | -5.14 (-11.07, 0.78) | Special sports training therapy |

For shoulder flexion, Table 24 and Fig 33 demonstrate that acupuncture plus exercise therapy yielded a favorable estimate compared with exercise therapy (MD 7.29, 95% CI 0.81 to 13.77), whereas special sports training versus exercise therapy showed a smaller but still favorable estimate (MD 3.85, 95% CI 1.00 to 6.71). In contrast, manual therapy versus exercise therapy was not statistically significant (MD 3.00, 95% CI −3.84 to 9.84). Overall, the pairwise comparisons support a potential advantage of acupuncture-based treatment for improving shoulder flexion, although uncertainty persisted for several comparisons.

**Fig 33.**
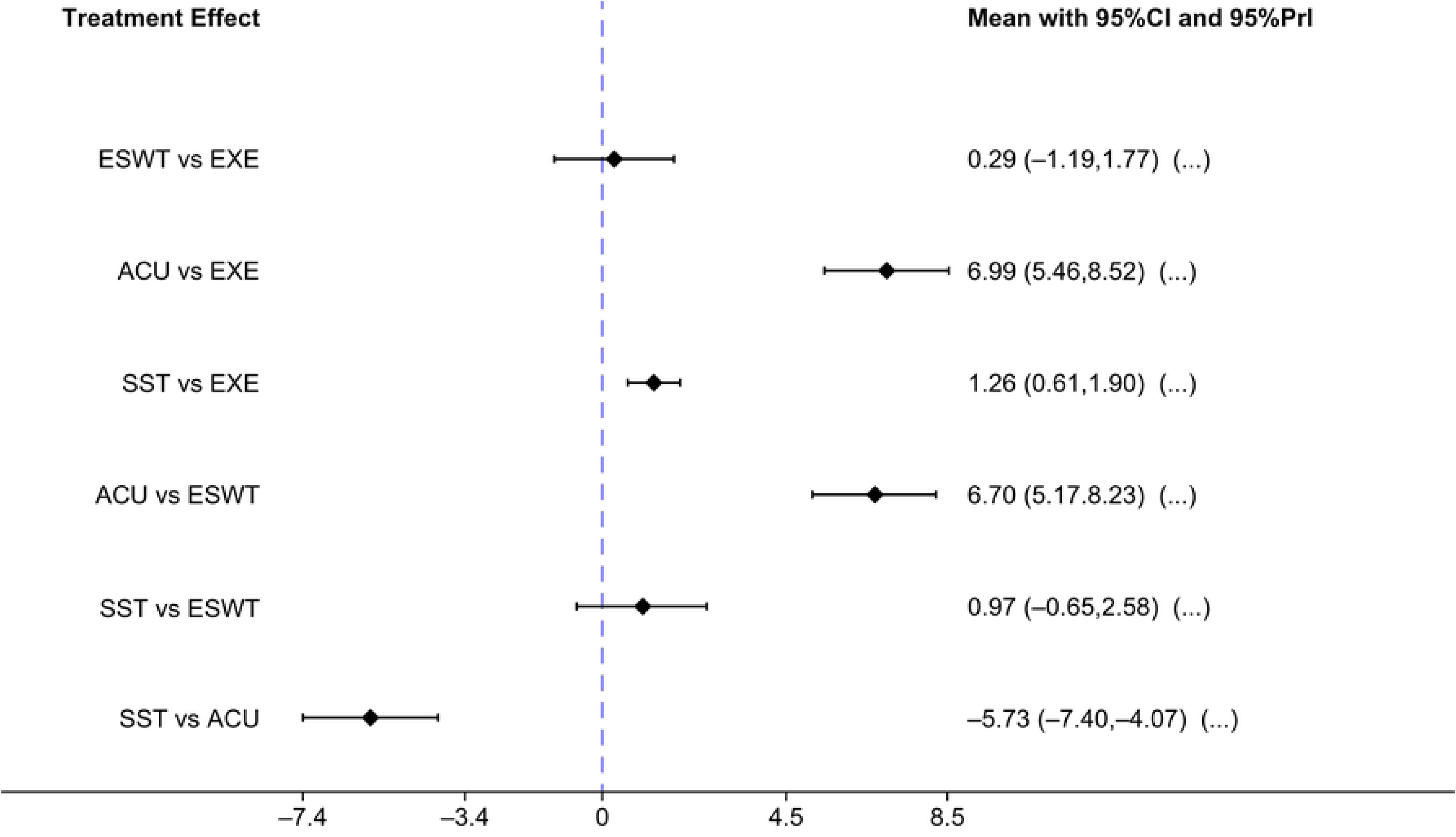
Pairwise comparison of treatment methods on the influence of patients with shoulder joint flexion range. ACU: acupuncture; EXE: exercise therapy; MAN: manual therapy; ESWT: extracorporeal shock wave therapy; SST: special sports training therapy; CI: confidence interval.

**Table 24.** Comparison of the influence of different treatment methods on the anterior flexion motion of shoulder joint.

|  |  |  |  |  |
| --- | --- | --- | --- | --- |
| Exercise therapy |  |  |  |  |
| 2.50 (-2.42, 7.42) | Extracorporeal shock wave therapy |  |  |  |
| 7.29 (0.81, 13.77) | 4.79 (-1.70, 11.28) | Acupuncture |  |  |
| 3.00 (-3.84, 9.84) | 0.50 (-7.93, 8.93) | -4.29 (-13.71, 5.13) | Manual therapy |  |
| 3.85 (1.00, 6.71) | 1.36 (-4.34, 7.05) | -3.44 (-10.52, 3.65) | 0.85 (-6.56, 8.27) | Special sports training therapy |

For shoulder abduction, Table 25 and Fig 34 demonstrate that acupuncture plus exercise therapy yielded the largest estimated improvement relative to exercise therapy (MD 3.36, 95% CI 1.42 to 5.30). Microwave diathermy (MD 3.30, 95% CI 1.20 to 5.40) and special sports training (MD 1.83, 95% CI 1.00 to 2.66) also showed favorable estimates, whereas manual therapy versus exercise therapy remained imprecise (MD 1.92, 95% CI −0.50 to 4.34). Although acupuncture showed the most favorable point estimate, the prediction intervals for several comparisons remained wide, indicating uncertainty in the consistency of these effects.

**Fig 34.**
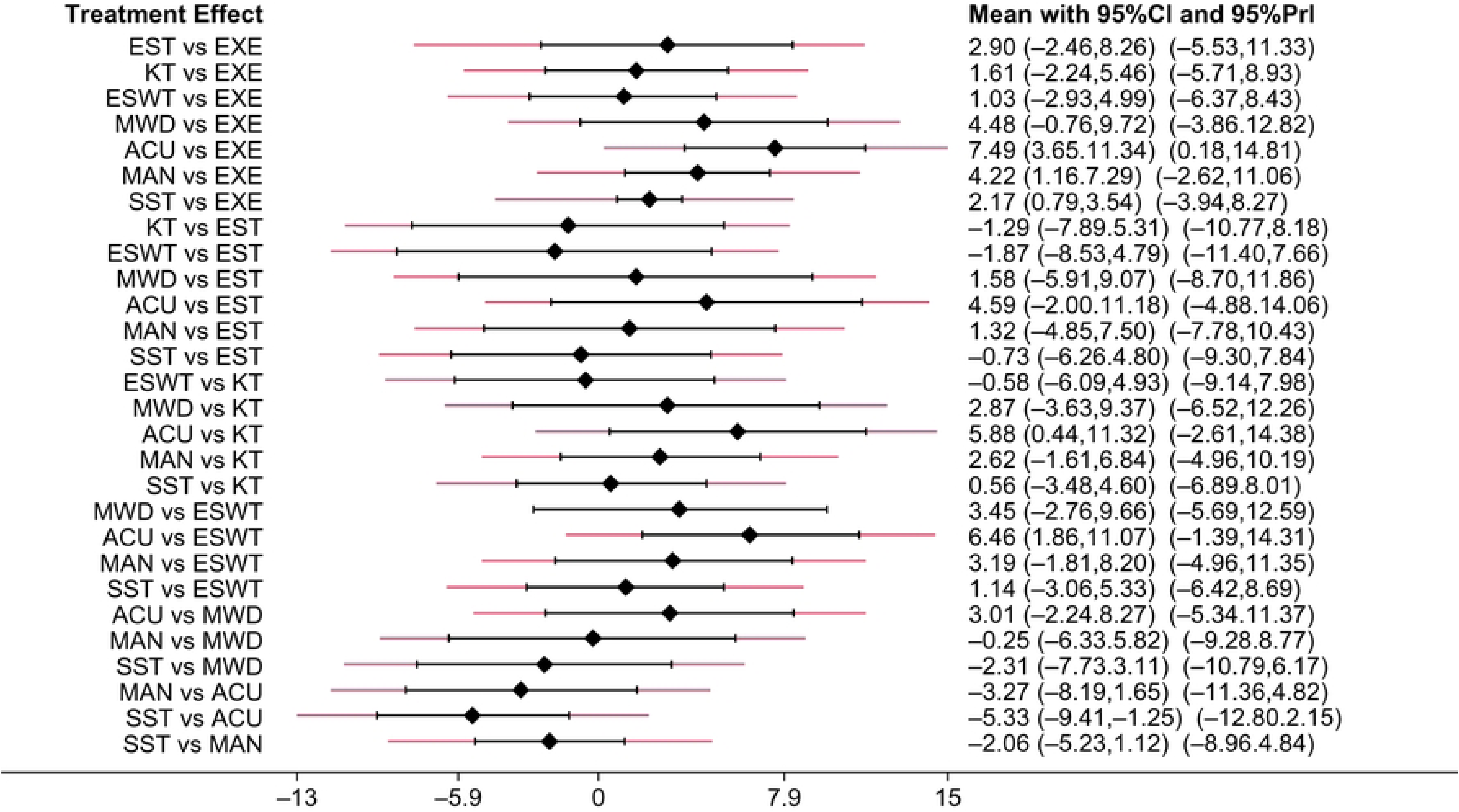
Pairwise comparison of the effects of different treatments on the abductive mobility of shoulder joint. ACU: acupuncture; EXE: exercise therapy; MAN: manual therapy; MWD: microwave diathermy; SST: special sports training therapy; CI: confidence interval.

**Table 25.** Pairwise comparison of the effects of different treatment methods on the abductive mobility of shoulder joint in patients.

|  |  |  |  |  |
| --- | --- | --- | --- | --- |
| Exercise therapy |  |  |  |  |
| 3.30 (1.20, 5.40) | Microwave diathermy |  |  |  |
| 3.36 (1.42, 5.30) | 0.06 (-2.01, 2.13) | Acupuncture |  |  |
| 1.92 (-0.50, 4.34) | -1.38 (-4.58, 1.82) | -1.44 (-4.54, 1.66) | Manual therapy |  |
| 1.83 (1.00, 2.66) | -1.47 (-3.73, 0.79) | -1.53 (-3.64, 0.58) | -0.09 (-2.64, 2.46) | Special sports training therapy |

For shoulder adduction, Table 26 and Fig 35 demonstrate that acupuncture plus exercise therapy yielded the most favorable estimate relative to exercise therapy (MD 6.99, 95% CI 5.46 to 8.52), followed by special sports training versus exercise therapy (MD 1.26, 95% CI 0.61 to 1.90). The comparison between acupuncture and extracorporeal shock wave therapy also favored acupuncture (MD 6.70, 95% CI 5.17 to 8.23). These findings suggest a relatively consistent advantage of acupuncture-based treatment for improving adduction, although the number of contributing studies was limited.

**Fig 35.**
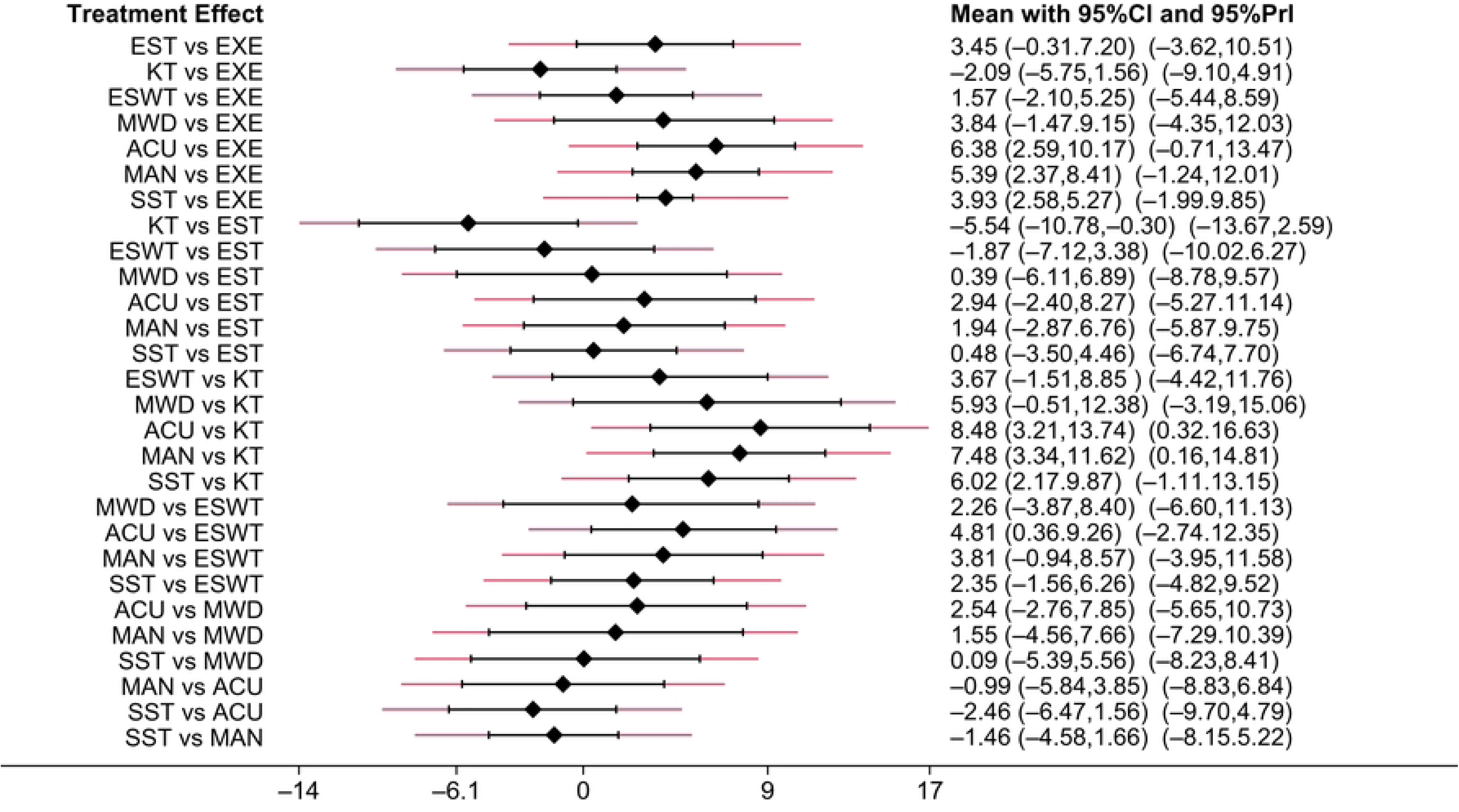
Pairwise comparison of different treatment methods on patients with shoulder adduction range of motion. ACU: acupuncture; ESWT: extracorporeal shock wave therapy; EXE: exercise therapy; SST: special sports training therapy; CI: confidence interval.

**Table 26.** Pairwise comparison of the effects of different treatment methods on the adduction motion of shoulder joint.

|  |  |  |  |
| --- | --- | --- | --- |
| Exercise therapy |  |  |  |
| 0.29 (-1.19, 1.77) | Extracorporeal shock wave therapy |  |  |
| 6.99 (5.46, 8.52) | 6.70 (5.17, 8.23) | Acupuncture |  |
| 1.26 (0.61, 1.90) | 0.97 (-0.65, 2.58) | -5.73 (-7.40, -40.7) | Special sports training therapy |

### Mesh league table: pain-related outcomes

Table 15 demonstrates a comparison of direct and indirect evidence for eight treatment methods based on VAS scores of 637 swimming athletes with shoulder impingement syndrome, together with the effect sizes and 95% CIs for each treatment method. Compared with exercise therapy alone, medium-frequency therapy plus exercise therapy showed the most favorable point estimate (MD −4.58, 95% CI −10.12 to 0.95), followed by acupuncture plus exercise therapy (MD −3.48, 95% CI −6.45 to −0.50).

## Discussion

We used a network meta-analysis of randomized controlled trials to compare the relative effects of available interventions for shoulder impingement syndrome (SIS) in swimmers. Overall, the findings show that treatment effects were outcome-specific rather than uniform across all domains. For pain relief, exercise combined with medium-frequency therapy showed the most favorable ranking profile, with extracorporeal shock wave therapy also performing well, whereas manual therapy demonstrated a significant advantage over exercise therapy in the direct effect-size analysis. For shoulder mobility, Kinesio taping and acupuncture appeared particularly favorable for external rotation, whereas acupuncture, manual therapy, and special sports training showed more consistent benefits for internal rotation. In addition, acupuncture and special sports training showed favorable effects on the IR/ER strength ratio, suggesting added value for restoring functional shoulder balance. Taken together, these findings indicate that treatment selection for swimmers with SIS should be individualized according to the athlete’s primary rehabilitation goal, whether pain reduction, restoration of specific shoulder movements, or improvement in rotational muscle balance.

The findings for pain outcomes should be interpreted with some caution. In the present analysis, conservative interventions did not produce uniformly significant effects across all subjective symptom measures, which may partly reflect the characteristics of the included populations. Most participants were described as having stage I or II SIS, and baseline symptom severity may therefore have been relatively modest. In addition, competitive swimmers often tolerate repeated shoulder loading and may exhibit relatively high pain thresholds, which may reduce the sensitivity of subjective scales to treatment-related change. Nevertheless, the present results still suggest that exercise combined with medium-frequency therapy may provide useful short-term symptom relief, and a similarly favorable signal was observed for manual therapy. From a practical standpoint, these approaches may be especially helpful when the immediate goal is to reduce pain sufficiently to improve participation in active rehabilitation, rather than to serve as stand-alone solutions. This interpretation is also consistent with the broader pattern of the network, in which exercise remained the common therapeutic foundation across many effective comparisons.

Shoulder range-of-motion outcomes showed a clearer and more differentiated treatment response than subjective symptom measures. Improvements were particularly evident for shoulder flexion, external rotation, and internal rotation. For external rotation, Kinesio taping, acupuncture, manual therapy, and special sports training all showed favorable effects relative to exercise therapy alone, whereas the ranking profile placed taping and acupuncture near the top. By contrast, for internal rotation, the more consistent benefits were observed with acupuncture, manual therapy, and special sports training, whereas Kinesio taping appeared less favorable. This difference between ER and IR is clinically meaningful, because it suggests that the same intervention may not influence all planes of shoulder movement to the same extent. In swimmers, where shoulder mechanics are highly repetitive and performance depends on efficient movement sequencing, such distinctions are especially important. A treatment that improves one direction of motion may not necessarily optimize overall shoulder function.

Several mechanisms may plausibly explain these movement-specific findings. Kinesio taping may offer relatively rapid assistance for external rotation by improving proprioceptive input, reducing excessive muscle tension, and enhancing movement support during dynamic activity. Acupuncture and manual therapy, in contrast, may exert broader effects through pain modulation, improvement in local circulation, reduction in periarticular soft-tissue restriction, and modulation of neuromuscular control. These pathways may be particularly relevant for internal rotation, which is often more closely linked to posterior shoulder tightness, rotator cuff imbalance, and altered scapulohumeral mechanics in symptomatic swimmers. Special sports training also showed favorable effects across several movement-related outcomes, which is not surprising given the sport-specific nature of swimmer’s shoulder dysfunction. Interventions that target scapular stability, rotator cuff coordination, and movement control under swimming-related loading conditions are more likely to transfer to actual swimming function than passive treatment alone. These explanations remain inferential, but they fit the overall direction of the network findings.

The findings for the IR/ER strength ratio add another clinically relevant dimension to the present study. In swimmers, this ratio is more informative than isolated strength values because it reflects the balance between the internal and external rotators, which is critical for shoulder stability and efficient stroke mechanics. In the present analysis, acupuncture and special sports training showed the most favorable profile for this outcome. This suggests that interventions aimed at restoring muscular coordination and movement control may be particularly valuable when treatment goals extend beyond short-term symptom relief and toward functional rehabilitation. Special sports training may improve the IR/ER ratio by enhancing rotator cuff strength, scapular control, and sport-specific movement efficiency, whereas acupuncture may contribute more indirectly by reducing pain, regulating muscle tone, and improving the quality of active movement during rehabilitation. Together, these findings reinforce the idea that symptom relief and functional restoration should not be treated as interchangeable endpoints in swimmer rehabilitation.

A clear innovation of the present study is that we move beyond conventional pairwise comparisons and provide a swimmer-specific comparative synthesis of multiple treatment strategies for SIS. Rather than asking whether a single intervention is effective in isolation, this network meta-analysis allowed direct and indirect comparison across a broad treatment network and across multiple clinically meaningful outcomes, including pain, shoulder range of motion, and rotational muscle balance. This is particularly valuable in swimmers, whose rehabilitation priorities often differ from those of the general population because pain control, preservation of training continuity, restoration of stroke-relevant mobility, and optimization of shoulder stability must all be considered simultaneously. Another important innovative feature is that the present study does not force a single overall conclusion across all outcomes. Instead, it shows that different interventions may be preferentially useful for different rehabilitation targets. This outcome-specific interpretation is more clinically realistic and provides a more practical basis for individualized treatment planning in swimmers with SIS.

The present study has several notable strengths. First, it focused specifically on swimmers, a population with distinctive biomechanical demands and rehabilitation priorities, thereby improving the clinical relevance of the synthesis. Second, the use of network meta-analysis allowed both direct and indirect comparisons across multiple conservative interventions, offering a broader comparative framework than conventional pairwise meta-analysis alone. Third, we evaluated multiple clinically meaningful outcomes rather than relying on pain alone, including shoulder mobility, muscle balance, and structural indicators. This makes the findings more useful for sports medicine practice, where return to training depends on symptom reduction and on restoration of shoulder function. Together, these strengths enhance the practical and translational value of the present work.

Despite these strengths, some limitations should be acknowledged. First, most included trials had relatively short follow-up periods, often no longer than one year, which limits conclusions regarding the long-term durability of treatment effects. Second, heterogeneity and local inconsistency were present in some outcome networks, and several comparisons were supported by only a small number of studies, which increases uncertainty around some estimates. Third, many included studies reported using only a single control group, and the overall risk of performance bias was not negligible. Fourth, although SUCRA rankings were informative, they should not be interpreted as proof of superiority independent of the corresponding effect estimates and CIs. Finally, several secondary outcomes were supported by limited data, and some prediction intervals remained wide, indicating that the apparent hierarchy of interventions should be interpreted cautiously. These limitations are especially important in network meta-analysis, where rankings can appear more definitive than the underlying evidence actually supports.

## Conclusion

This network meta-analysis demonstrated that no single intervention can be considered universally optimal for swimmers with SIS. Instead, treatment effectiveness appears to depend on the clinical target. Exercise remained the core therapeutic foundation across most comparisons, while several adjunctive conservative interventions showed additional value for specific rehabilitation goals. Exercise combined with medium-frequency therapy appeared more useful when pain relief was the priority, whereas Kinesio taping and acupuncture were more favorable for improving external rotation. For internal rotation and restoration of rotational muscle balance, acupuncture, manual therapy, and special sports training showed more consistent advantages.

The importance of the present study lies in its swimmer-specific, outcome-oriented comparison of multiple conservative treatment strategies. By showing that different interventions may be preferentially useful for different rehabilitation targets, we provide a clearer evidence base for individualized management of swimmer’s SIS. These findings are clinically relevant because they support more targeted treatment selection in a population where pain control, movement quality, and maintenance of training continuity must often be balanced at the same time. Further high-quality swimmer-specific trials with larger samples and longer follow-up are needed to confirm the durability of these effects and to refine treatment decision-making.

## Data Availability

All data generated or analyzed during this study are included in this published article.

## Acknowledgments

We would like to thank Editage (www.editage.cn) for English language editing.

